# Associations of employment status, working time and job satisfaction with sleep duration and sleep quality among the Japanese 50+ population

**DOI:** 10.1101/2024.03.28.24305011

**Authors:** Jacques Wels, Rong Fu

**Affiliations:** Free University of Brussels (ULB), EpiSoc, Belgium; University College London, MRC unit for Lifelong Health and Ageing, United Kingdom; Waseda university, School of commerce, Japan

## Abstract

**Background:** Few studies have captured the relationship between employment status, working time and job satisfaction and sleep duration and quality in Japan where poor sleep quality and low sleep duration are major public health concerns.

**Methods:** We use four waves from the Japan Study of Aging and Retirement (JSTAR) to assess the relationship between employment status and self-reported job satisfaction and sleep duration and self-reported sleep quality. We control for socio-demographic characteristics, working time and self-reported measures of health. The initial sample includes 7,082 respondents. We use mixed effects modified Poisson regression for binary outcomes for sleep quality and linear mixed effects for sleep duration and multiple imputations to correct for sample attrition.

**Results:** No major difference is observed between employment status and poor sleep quality except for housekeepers (0.123 [ 95%CI: 0.041; 0.205]) in comparison with full-time employed workers. All categories of workers tend to report sleeping longer than full-time employees with higher hours among those who retired (0.339 [95%CI: 0.218; 0.460]). Poor job satisfaction is associated with higher risks of self-reported poor sleep quality (0.230 [95%CI: 0.040, 0.421]) and waking up at night (0.362 [95%CI: 0.025, 0.699]) but the associations fade away when controlling for other health measurements (respectively, −0.137 [95%CI: −0.328, 0.054] and 0.092 [95%CI: −0.248; 0.432]).

**Conclusion:** Retirement increases sleep duration without improving sleep quality and housekeepers sleep longer but with poorer sleep quality. Job satisfaction is a major cofounder of sleep quality among the workforce but the effect is mediated by physical and mental health levels.

## Background

Sleep duration is low in Japan in comparison with other top economies. On average, 56 percent of the Japanese population sleeps less than seven hours against, respectively, 45 percent in the US, 35 in the UK, 30 in Germany and 26 in Canada ^1^. This is not a new trend as sleep duration in Japan has been declining since the 1960s ^2^. In 2014, “good sleep” became a policy priority with specific sleep guidelines published for different generations ^3^ although mainly focusing on individuals’ behaviours and not targeting the social mechanisms leading to poor sleep quality and short sleep duration among which work and employment are usually seen as detrimental ^4,5^. As a matter of fact, despite regulations passed over the past decades to prevent long working hours, working time remains high in Japan ^6^ and the labour market is fragmented with many women – particularly among the oldest generations – remaining out of the labour market (as housekeepers) ^7^ and older workers sometimes downgraded to specific employment statuses (contract work) ^8,9^. The relationship between work and employment and sleep is of interest in such a context as low sleep duration and poor sleep quality both have detrimental effects with sleep disturbances associated with depressive symptoms among the older population ^10^ and low (<6 h) and high sleeping time (>9h) associated with higher mortality risks ^11^. However, despite many studies on sleep in Japan, little attention has been paid on the explanatory role of work and employment among the ageing population.

Typically, when questioning such a relationship, two different dimensions are examined. On the one hand, the employment status – that refers to the classification of an individual’s relationship with an employer, indicating whether they are employed, unemployed, self-employed, or inactive in the labour force – is seen to play a crucial role in explaining sleep patterns. A substantial amount of research was produced on the relationship between transition from work to retirement and sleep quality and duration using longitudinal data. It was shown that retirement is not only associated with short-term reductions in sleep difficulties but also increase in sleep duration over 1 to 2 years ^12,13^. Results are similar using panel data from France ^14^. Using longer follow-up longitudinal data, it was demonstrated that these positive effects last over time for non-restorative sleep, premature awakening and restless sleep ^15^ with potential greater effects on female as well as greater effects on those retiring from part-time jobs ^16^. By contrast, comparing white collar Japanese employees with the rest of the population, a cross-sectional study ^17^ has shown a higher prevalence of poor sleep quality (based on the Pittsburgh Sleep Quality Index (PSQI)) – between 30 and 45 percent – among the former. Significant factors associated with poor sleep included stress, job dissatisfaction, being unmarried, lower education, younger age and poor sleep quality was associated with absenteeism, poor health, work and relationship problems and workplace accidents. It was shown that higher grades of employment within the civil servant workforce are associated with better sleep quality ^18^ and this echoes a more general finding where employment insecurity increases the risk of sleep disturbance independently of country context ^19^. However, non-employed people are also at risk. It was evidenced that unemployment (i.e., those looking for a job) and non-employment are associated with high insomnia-related symptoms prevalence ^20^, particularly among the male population and to a lesser extent among the female population ^21^. Both employment and non-employment may affect sleep duration and quality through different mechanisms but the amount of evidence is limited when it comes to addressing short sleep and poor sleep quality across different statuses ^20^.

On the other hand, work exposure has been given considerable of interest with a particular focus on job satisfaction and working time. One issue when looking at the studies on the Japanese workforce is that most of them target specific sub-groups of workers with little information on female workers, contingent workers, or the owners of an independent business ^22^. A cross-sectional study ^23^ conducted with factory workers aged 20-59 has shown that high job work-related stress was associated with lower sleep quality, while various lifestyle habits were linked to different sleep characteristics. In another survey on factory workers ^24^ authors find sleep differences by gender, age, and work pattern. Women reported fewer awakenings and less napping, older males had shorter sleep but less reported sleep insufficiency, and shift workers experienced longer sleep onset times, more awakenings, but less napping and caffeine intake. Another study of 334 female daytime workers at a Japanese electric equipment manufacturer ^25^ explored the link between perceived job stress and sleep habits, focusing on factors like job control, workload, and social support, as well as sleep quality indicators such as sleep duration and insomnia. Findings indicated correlations between job stress aspects and sleep habits: skill underutilization positively affected sleep duration, cognitive demands reduced daytime napping and sleepiness, and overtime was linked to shorter sleep but more frequent poor sleep quality. Social support from supervisors, co-workers, and family or friends showed negative correlations with several indicators of poor sleep, suggesting that social support can alleviate some stress-related sleep issues. Similarly, work-related stress is associated with poorer sleep quality independently of stress experiences at home ^26^.

Another feature of work is working time. Whilst high working hours tend to be associated with shorter sleep duration ^27,28^, studies on working time and health are few in Japan with most dealing with the impact of poor sleep on productivity ^29^ or work injuries ^30^. Mafune and Yokoya ^31^ show that workers with over 100 hours of overtime experiencing less than 6 hours of sleep, late dinners, and increased dining out. Night shift workers also reported more frequent awakenings during sleep. The conclusion highlighted that around 30% of the surveyed temporary workers were at risk of overwork-related health issues, including insufficient sleep, late meals, and mental health symptoms, suggesting a need for regulations to prevent excessive overtime requests. Early start of the working day is associated with lower sleep duration, sleep problems, and fatigue ^32^. Working time is also associated with sleep duration, with those working less that 8 hours a day sleeping more than those working more than 8 hours a day ^33^.

Whilst most previous studies have focused on specific segments of the Japanese population, the main objective of this study is to look at the associations between employment status, job satisfaction and working time among a nationally representative sample of the Japanese population aged 50 and over. By doing so, we provide insights into potential non-behavioural factors contributing to low sleep duration and poor sleep quality, with the intention of informing public policy decisions.

## Methods

### Data source and samples

Data come from the Japanese Study of Aging and Retirement (JSTAR), a longitudinal dataset that currently contains four waves collected in 2007, 2009, 2011 and 2013. The original sample strategy is described in ^34^. At the baseline (2007), JSTAR includes respondents aged 50 to 75 living in five municipalities in eastern area of Japan ^34^: Takikawa in Hokkaido, Sendai in the Tohoku area, Adachi Ward within Tokyo, Kanazawa in Hokuriki and Shirakawa in the Chubu area. Two refreshment samples were collected in 2009 and 2011 to increase the number of cities. The 2009 wave includes a refreshment sample from two additional cities (Tosu and Naha) and the 2011 wave includes another refreshment sample from three additional cities (Chofu, Hiroshima and Tondabayashi). In total, the sample contains information on ten cities. Sample design is shown in <u>supplementary file 1</u>. The full sample includes 18,762 observations over four time-points, including a total of 7,082 respondents.

### Sleep duration and sleep quality

Information on sleep quality and sleep duration was not collected the same way in all waves. Information on sleep quality was collected over each wave as part of the General Health Questionnaire (GHQ) with a question on poor sleep frequency per week (not at all, one to two days a week, three to four days a week and five days a week or more). The variable was recoded as binary (0: not at all; 1: 1 day a week or more). Information on sleep duration during weekdays was collected in waves 2009 and 2011 (both follow-up and refreshment) as well as in wave 2013. The variable is numeric and calculated in hours. Finally, waves 2011 (refreshment sample only) and 2013 contain several specific questions on time to fall asleep, times waking up during the night, waking up in early hours and waking up to urinate. Time to fall asleep was re-coded as a binary variable with two modalities: 0: 30 minutes or less; 1: More than 30 minutes. Times waking up during the night and waking up in the early hours were also coded as binary with the following modalities: 0: Never or one or two times a week; 1: More than one or two times a week. Finally, times waking up to urinate was coded with two modalities: 0: Never or once per night; 1: Two times per night or more. Full information on coding and variables availability is show in <u>supplementary file 2</u>.

To include the maximum amount of information contained in JSTAR, we have generated three sub-samples. *Sample 1* focuses on poor sleep (sleep quality) across all the four JSTAR waves. *Sample 2* includes poor sleep as well as sleep time in weekdays across three waves (excluding wave 2007). Finally, *sample 3* includes all the six variables across two waves (the 2011 refreshment sample and the 2013 sample). Analyses were replicated on each sample. Differences across sub-samples are briefly discussed but only results flowing from the maximum size samples are reported in the result section.

### Employment status, working time and job satisfaction

Two models are used in this study. Model 1 looks at the relationship between employment status and sleep quality and duration, whilst Model 2 selected the working population (employment or self-employment) and looks at the relationship between sleep quality and duration and working and job satisfaction. The employment status contains 13 modalities that distinguish different positions within and outside the labour market on which information was collected in the survey: (1) employed full-time, (2) company executive, (3) employed part-time, (4) employed under contract, (5) temporary employed, (6) employed other, (7) owner of independent business, (8) help in independent business, (9) side job at home, (10) retired, (11) not retired – receiving medical care, (12) keeping house (13) inactive for other reasons. The first modality (employed full-time) – that is the most represented among the working population – is selected as the reference category. Job satisfaction was assessed using a self-reported variable (‘Overall, I am satisfied with my current job’). The variable contains four modalities: strongly agree, somewhat agree, do not really agree and strongly disagree. The first category is selected as the reference. Working time was calculated based on self-reported working time per week including overtime and was categorized over six modalities: less than 20 hours/week, 21 to 30 hours/week, 31 to 40 hours/week, 41 to 50 hours/week, 51 to 60 hours/week, more than 60 hours/week.

### Covariates and adjustment levels

The model controls for the following covariates: age (in years of age); gender (female, male – male being the reference category); highest level of education obtained (distinguishing elementary to middle school, high school, junior college, vocational school and university degree – the latter being the reference category); marital status (distinguishing those who are married or have a common law spouse versus those who are not); whether the household borrowed money to friends or family (‘yes’, ‘no’ – ‘no’ being the reference category); whether the household rent their accommodation (‘yes’, ‘no’ – ‘no’ being the reference category); whether the respondent has a private health insurance under their own name (‘yes’, ‘no’ – ‘no’ being the reference category); self-reported health (coded on a 5-item scale ranging from ‘good’ to ‘Not good and used as a numeric variable); multimorbidity (distinguishing respondents who declared having been diagnosed with at least two health conditions including heart disease, high blood pressure, hyperlipemia, cerebral accident, diabetes, chronic lung disease, liver disease, ulcer or stomach disorder, joint disorder, bladder disorder, depression or emotional disorder, cancer); General Health Questionnaire (GHQ-20) caseness that is based on the answers to twenty items (from 1. Not at all to 5. 5 days a week or more) that are summed up and categorized into two categories across the time-point means (‘yes’ or ‘no’, ‘no’ being the reference category); whether the respondent was an outpatient at an hospital over the past year (‘yes’ or ‘no’, ‘no’ being the reference category); whether the respondent spent at least one night at the hospital over the past year (‘yes’ or ‘no’, ‘no’ being the reference category); life satisfaction that distinguishes those reporting being somewhat unsatisfied or unsatisfied and those reporting being satisfied or very satisfied (reference category). All the variables used in this study are time-varying except gender, highest level of education, marital status and whether the respondent has a health insurance that are fixed.

The models includes three layers of adjustment: (1) The *unadjusted model* only controls for gender and age. (2) The *socio-demographic adjusted* model additively controls for the highest level of education obtained, the marital status, whether the household borrowed money to friends or family, whether the household rent or own its accommodation and whether the respondents had a private health insurance.(3) The *fully adjusted model* additively controls health variables including self-reported health, whether the respondent was diagnosed with two or more health conditions (multimorbidity), GHQ-20, whether the respondent was an outpatient at an hospital over the past year, whether the respondent spent the night at the hospital and life satisfaction.

By using these three layers of adjustment we provide crude estimates that control for basic demographics but also socio-demographic covariates adjusting for potential behavioural cofounders (that could be competing exposures) as well as health and mental health variables that may be associated with poor sleep and we be on the way of the relationship between work and employment and sleep quality and duration ^35^. Model specifications by adjustment level are shown in <u>supplementary file S3</u>.

### Statistical analyses

We use a mixed effects Modified Poisson Regression for binary outcomes with robust standard errors (sandwich estimator) ^36,37^ that allows to calculate the Relative Risks instead of the Odds Ratios ^38^ for sleep quality measurements. Models including sleep duration as an outcome are calculated based on a linear mixed effects modelling. In other words, results for sleep duration can be interpreted in hours whilst results for sleep quality variables should be interpreted in terms of relative risks. The model is replicated twice. First, we look at the association between the employment status and sleep duration and quality (model 1). Second, we examine the association between job satisfaction and sleep duration and quality among the working population (i.e., excluding those not in employment) (model 2). We also assess data missingness across waves and apply multiple imputations to correct sample bias due to attrition. We then meta-analyse the estimates flowing from the subsequent imputed datasets. We also compare results flowing from the non-imputed models with results flowing from the imputed model. Multiple imputations were replicated for each sub-sample separately.

## Results

Descriptive statistics for the exposure variables are shown in table 1 whilst full descriptive statistics are shown in <u>supplementary file S4</u>. Poor sleep concerns about 30 percent of the sample and is stable over time with a lowest of 27.6 percent in wave 2013 and a highest of 34.3 percent in the 2011 refreshment sample. Sleep duration during weekdays was not asked in wave 2007. Subsequent waves show that sleep duration is, on average, less than 7 hours with 7 hours in wave 2009 and 6.8 hours in wave 2013. Finally, sleep quality indicators were only asked in sample 3 (wave 2011 and 2013) with between 17.1 and 25.8 percent of the sample reporting taking more than 30 minutes to fall asleep, between 23.9 and 30.9 percent of the sample waking up more than one or two times a week, 20.9 and 26.5 percent waking up in the early hours more than one or two times a week, and 21.9 and 25.9 percent waking up to urinate 2 times or more per night.

**Table 1.**
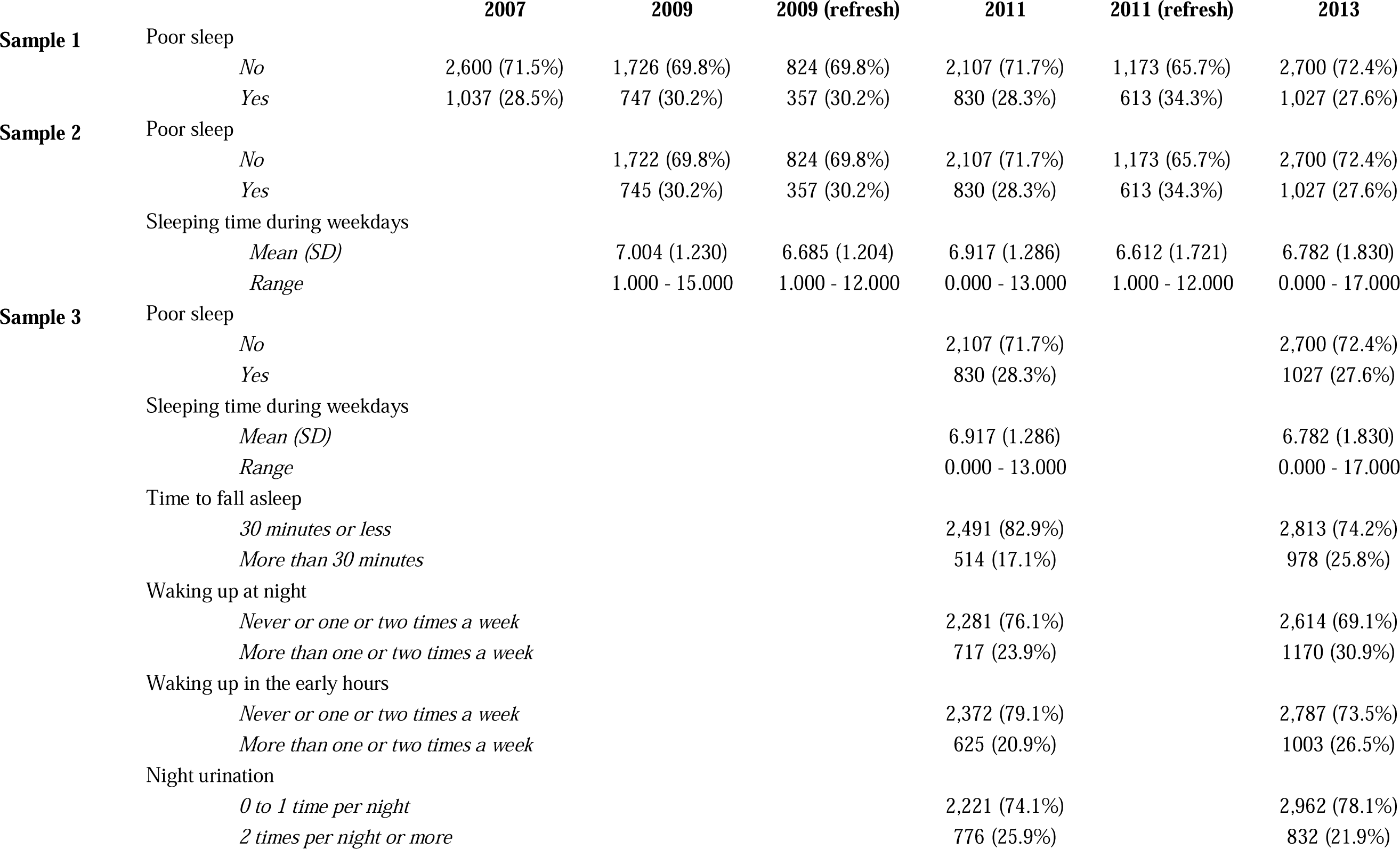
Descriptive statistics on sleep quality and duration by JSTAR wave.

|  |  | **2007** | **2009** | **2009 (refresh)** | **2011** | **2011 (refresh)** | **2013** |
| --- | --- | --- | --- | --- | --- | --- | --- |
| **Sample 1** | Poor sleep |  |  |  |  |  |  |
|  | *No* | 2,600 (71.5%) | 1,726 (69.8%) | 824 (69.8%) | 2,107 (71.7%) | 1,173 (65.7%) | 2,700 (72.4%) |
|  | *Yes* | 1,037 (28.5%) | 747 (30.2%) | 357 (30.2%) | 830 (28.3%) | 613 (34.3%) | 1,027 (27.6%) |
| **Sample 2** | Poor sleep |  |  |  |  |  |  |
|  | *No* |  | 1,722 (69.8%) | 824 (69.8%) | 2,107 (71.7%) | 1,173 (65.7%) | 2,700 (72.4%) |
|  | *Yes* |  | 745 (30.2%) | 357 (30.2%) | 830 (28.3%) | 613 (34.3%) | 1,027 (27.6%) |
|  | Sleeping time during weekdays |  |  |  |  |  |  |
|  | *Mean (SD)* |  | 7.004 (1.230) | 6.685 (1.204) | 6.917 (1.286) | 6.612 (1.721) | 6.782 (1.830) |
|  | *Range* |  | 1.000 - 15.000 | 1.000 - 12.000 | 0.000 - 13.000 | 1.000 - 12.000 | 0.000 - 17.000 |
| **Sample 3** | Poor sleep |  |  |  |  |  |  |
|  | *No* |  |  |  | 2,107 (71.7%) |  | 2,700 (72.4%) |
|  | *Yes* |  |  |  | 830 (28.3%) |  | 1027 (27.6%) |
|  | Sleeping time during weekdays |  |  |  |  |  |  |
|  | *Mean (SD)* |  |  |  | 6.917 (1.286) |  | 6.782 (1.830) |
|  | *Range* |  |  |  | 0.000 - 13.000 |  | 0.000 - 17.000 |
|  | Time to fall asleep |  |  |  |  |  |  |
|  | *30 minutes or less* |  |  |  | 2,491 (82.9%) |  | 2,813 (74.2%) |
|  | *More than 30 minutes* |  |  |  | 514 (17.1%) |  | 978 (25.8%) |
|  | Waking up at night |  |  |  |  |  |  |
|  | *Never or one or two times a week* |  |  |  | 2,281 (76.1%) |  | 2,614 (69.1%) |
|  | *More than one or two times a week* |  |  |  | 717 (23.9%) |  | 1170 (30.9%) |
|  | Waking up in the early hours |  |  |  |  |  |  |
|  | *Never or one or two times a week* |  |  |  | 2,372 (79.1%) |  | 2,787 (73.5%) |
|  | *More than one or two times a week* |  |  |  | 625 (20.9%) |  | 1003 (26.5%) |
|  | Night urination |  |  |  |  |  |  |
|  | *0 to 1 time per night* |  |  |  | 2,221 (74.1%) |  | 2,962 (78.1%) |
|  | *2 times per night or more* |  |  |  | 776 (25.9%) |  | 832 (21.9%) |

Figure 1 exhibits the estimates and 95 percent confidence intervals for the association between employment status and poor sleep quality for the different levels of adjustment after multiple imputations (model 1). Full estimates are sown in <u>supplementary file S5</u>.

**Figure 1.**
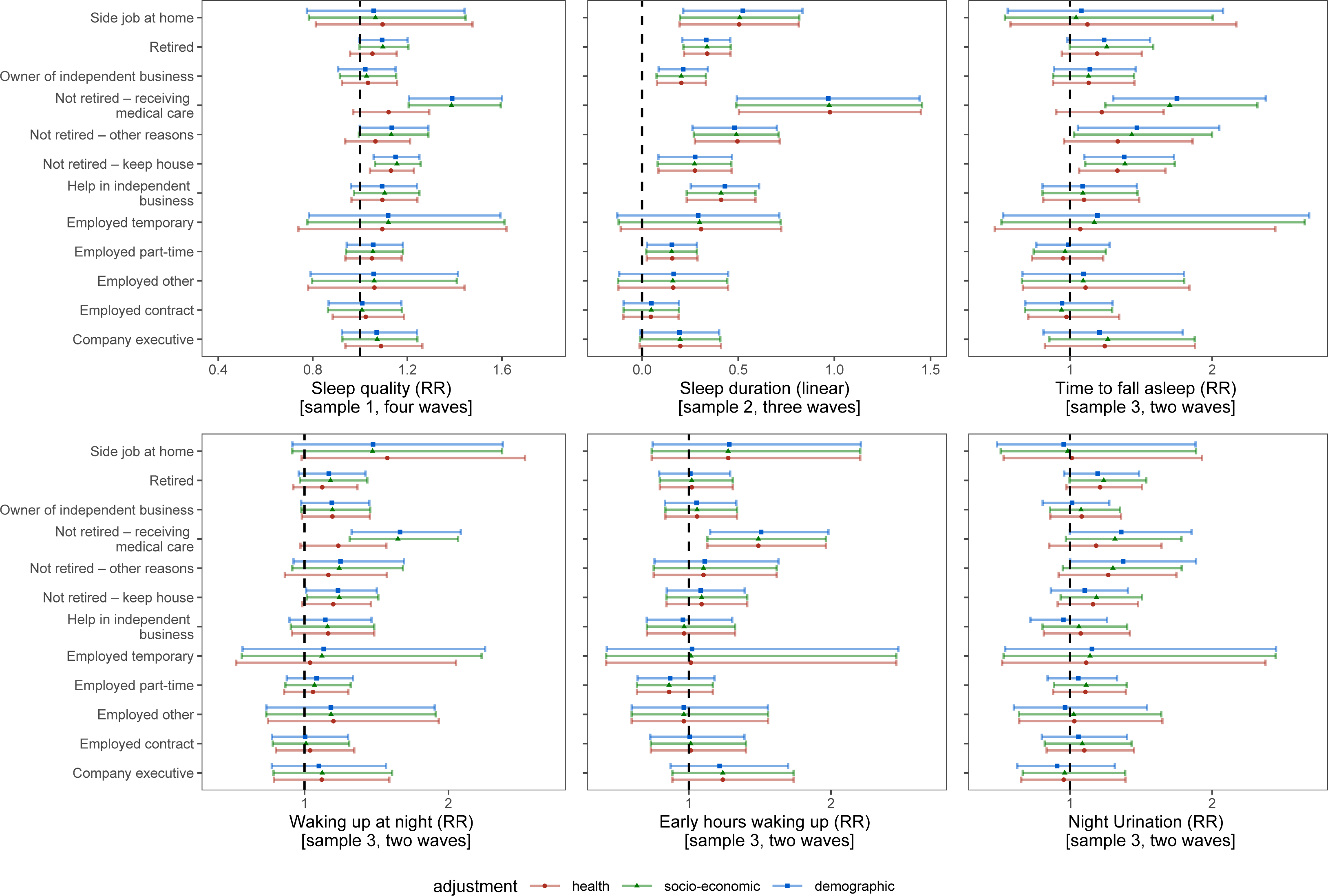
Association between employment category and sleep duration and quality (model 1 with multiple imputations)

Looking at sleep quality for sample 1, we observe a slightly positive relationship between being retired and poor sleep quality in comparison with being full-time employed after controlling for demographic and socio-economic characteristics (RR=0.092 [95%CI: −0.002; 0.186]) but the relationship is not significant when controlling for health (RR=0.051 [95%CI: −0.043; 0.144]). Similarly, receiving medical care is highly associated with poor sleep quality risks for both levels of adjustment (RR=0.327 [95%CI:0.187; 0.467]) but the health adjusted model is not significant (RR=0.092 [95%CI: −0.029; 0.257]). By contrast, housekeeping is associated with poor sleep quality risks in the three levels of adjustment (RR, health adjustment=0.123 [95%CI: 0.041; 0.205]).

A different pattern is observed for sleep duration in sample 2. Those retired, receiving medical care, not retired for other reasons (i.e., inactive), housekeepers, those helping in independent business and those in part-time employment report higher sleep duration in comparison with the full-time employed workers. Particularly high coefficients are observed in the fully adjusted model for those retired (0.339 [95%CI: 0.218; 0.460]), having a side job at home (0.505[ 95%CI: 0.195; 0.815]) or receiving medical care (0.977 [95%CI: 0.504; 1.450]). Adjusting for self-reported health and psychological distress reduces the magnitude of the risk for those receiving medical care and the retired population but there does not seem to have a proper impact of controlling for these variables when looking at sleep duration.

Finally, looking at the other measurements of sleep quality for sample 3, we observe low levels of significance that are due both to the small sample sizes and little change the is observed over time. However, a few associations are informative. First, we observe highest risks in the socio-economic adjusted model of taking more than 30 minutes to fall asleep for those receiving medical care (RR=0.532 [95%CI: 0.222; 0.841]), for those inactive (RR=0.361 [95%CI: 0.029; 0.693]) as well as for the housekeepers (RR=0.327 [95%CI: 0.102; 0.552]). This is true when adjusting both demographic and socio-economic covariates, but the confidence intervals overlap the null hypothesis (RR=1) when adjusting for health, except for housekeepers where the risk remains significant (RR=0.288 [95%CI: 0.062; 0.514]). The same trend is observed for those waking up at night with higher risks for those receiving medical care and keeping the house but not significant when controlling for heath. Estimates are not significant for waking up in the early hours and night urination except for those receiving medical care.

The main results for the imputed model 2 are shown in figure 2. The figure particularly focuses on working time and job satisfaction. Full estimates are sown in <u>supplementary file S6.</u>

**Figure 2.**
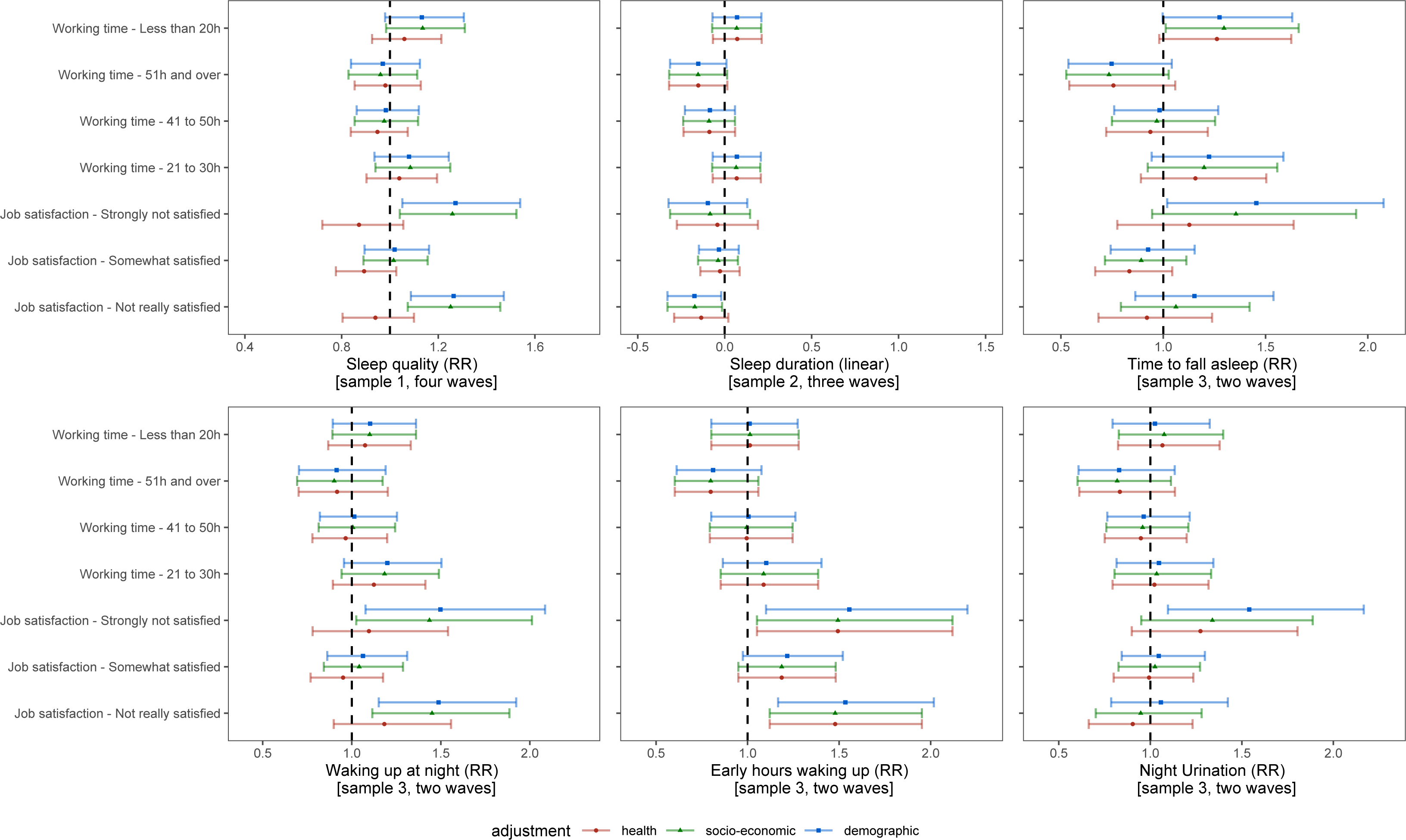
Association between working time and job satisfaction and sleep duration and quality (model 2 with multiple imputations)

Looking at working time, we observe no significant difference between working 51 hours and over, 41 to 50 hours and 21 to 30 hours per week in comparison with those working 31 to 40 hours a week. Sleep quality and job satisfaction are not associated with working time pattern, indicating that that those working below 50 hours a week do not trade their working time for sleep duration and do not report a better sleep quality. However, highest levels of working time are associated with slightly lower sleep duration (−0.152 [95%CI: −0.320; 0.015]), independently of individuals’ demographic, socio-economic or health characteristics.

What plays a major role in explaining sleep duration and, particularly, sleep quality is job satisfaction. Compared with those satisfied with their job, those not really satisfied or strongly not satisfied report higher risks of poor sleep quality in the socio-economic adjusted model (respectively, RR=0.224 [95%CI: 0.071; 0.376] and RR=0.230 [95%CI: 0.040; 0.421]). However, after controlling for self-reported health and psychological distress, the associations are reversed and the confidence intervals overlap the null hypothesis (respectively, RR=-0.062 [95%CI: −0.218; 0.095] and RR=-0.137 [95%CI: −0.328; 0.054]). There is also some evidence that those who report not being really satisfied with their job have a lower sleep duration compared with those satisfied with their job (socio-demographic adjustment: −0.172 [95%CI: −0.329; −0.015]), after controlling for working time, which may indicate that job satisfaction affects sleep duration independently of the time spent in work activities. Similarly, one observes a strong association between the risk of waking up in the early hours and being strongly not satisfied with one’s job (RR=0.401[95%CI: 0.050; 0.751]).

Analyses were replicated using complete cases (i.e., before multiple imputations) with no major difference across estimates, as can be seen in <u>supplementary files 5 and 6</u>.

## Discussion

Individual traits are often targeted as the cause of poor sleep and low sleep duration. Alcohol consumption, cigarette smoking or unhealthy dietary habits ^39^ as well as being unmarried ^40^ indeed play a role in explaining sleep problems within the Japanese population. Policy interventions have long tried to address sleep issues in Japan with interventions targeting individual behaviours or sleep patterns (e.g. napping) ^41^. Whilst these are fruitful to some extent, improving sleep among the Japanese 50+ population would also require addressing the social determinants of sleep among which work and employment play a key role.

The Japanese labour market among the 50+ population is fragmented across different employment statuses. A significant part of such a population is out of the labour market with 22 percent keeping house and 12 percent retired. Housekeeping, particularly among the female population, remains an important status for such older cohorts and is associated with both increased sleeping time and poor sleep quality, independently of socio-demographic characteristics and physical and mental health responses. The same is observed among the retired population but with lower intensity in terms of poor sleep quality and the cofounding effect of physical and mental health. Housekeepers appear to be at particular risk of poor sleep quality and this has not been observed in previous studies.

When focusing on the population in employment, we unsurprisingly observe that high working time tend to be associated with shorter sleep duration but better sleep quality. What is new in this study is that we show that poor job satisfaction, after controlling for working time and employment status, is strongly associated with greater risks of poor sleep quality. However, such a relationship is somehow mediated by poor physical and mental health indicating that poor sleep quality is part of a more general process in which job dissatisfaction leads to a decline in both health and sleep quality. Addressing high working hour patterns (more than 40 hours per week) and improving job satisfaction would contribute to increase sleep duration, on the one hand, and sleep quality, on the other.

This study is not without limitations. A first limitation concerns the dataset itself. JSTAR contains only four waves with questions on sleep quality which were not systematically replicated across waves. That is why we have utilised three specific sub-samples. Associations for the main exposure variables (sleep duration and overall sleep quality) were replicated across these sub-samples, indicating no fundamental differences across estimates. This also explains broader confidence intervals for the variables not replicated in the first two waves. A second limitation pertains to causation. We used a directed acyclic graph (DAG) (s<u>upplementary file 2</u>) to illustrate how adjustment levels control for cofounders and competing exposures. However, sample sizes did not allow for stratification or interaction effects, limiting our ability to infer causal pathways. It has been evidenced that there is a reciprocal relationship between work stress and poor sleep ^42^ and that low sleep quality is associated with an increase in work-related stress ^43^. Similarly, poor sleep leads to fatigue, which may in return affect work ^44,45^. By contrast, job satisfaction is recognised to be influenced by workplace determinants more than by workers’ characteristics ^46^, thus limiting the risk of bidirectionality. Similarly, employment status and working time cannot be considered as the product of poor sleep. Nevertheless, we refrained from using causal language avoid misleading interpretations of our findings. Finally, the used of mixed effects instead of a fixed effect modelling is justified by the small number of respondents transitioning from one status (or working time pattern or job satisfaction category) to another over time. Mixed effect are also more flexible to produce relative risks ratios.

## Data Availability

All data produced in the present study are available upon request to the Japanese Research Institute of Economy, Trade & Industry (RIETI) (https://www.rieti.go.jp/en/projects/jstar/)

## Authors contribution

Conceptualisation: JW; Supervision: JW, RF; Project Administration: JW; Investigation: JW; Formal Analysis: JW; Software: JW; Methodology: JW, RF; Validation: RF; Data Curation: JW; Resources: JW; Funding Acquisition: JW; Writing - Original Draft Preparation: JW; Writing - Review & Editing: RF; Visualization: JW

