## Supplementary files 1-6 for "Associations of employment status, working time and job satisfaction with sleep duration and sleep quality among the Japanese 50+ population"

### Supplementary file S1. Longitudinal samples composition

#### Sample 1. Sample composition

| **WAVE 1** |  | **WAVE 2** |  | **WAVE 3** |  | **WAVE 4** |
| --- | --- | --- | --- | --- | --- | --- |
| 2007 |  | 2009 |  |  |  |  |
| (5 prefectures) |  | (5 prefectures) |  |  |  |  |
| N=3,862 |  | N=2,718 |  |  |  |  |
|  |  | 2009 |  | 2011 |  |  |
|  |  | (2 prefectures) |  | (7 prefectures) |  |  |
|  |  | N=1,440 |  | N=3,847 |  |  |
|  |  |  |  | 2011 |  | 2013 |
|  |  |  |  | (3 prefectures) |  | (10 prefectures) |
|  |  |  |  | N=1,966 |  | N=4,929 |

#### Sample 2. Sample composition

| **WAVE 1** |  | **WAVE 2** |  | **WAVE 3** |  | **WAVE 4** |
| --- | --- | --- | --- | --- | --- | --- |
|  |  | 2009 |  |  |  |  |
|  |  | (5 prefectures) |  |  |  |  |
|  |  | N=2,718 |  |  |  |  |
|  |  | 2009 |  | 2011 |  |  |
|  |  | (2 prefectures) |  | (7 prefectures) |  |  |
|  |  | N=1,440 |  | N=3,847 |  |  |
|  |  |  |  | 2011 |  | 2013 |
|  |  |  |  | (3 prefectures) |  | (10 prefectures) |
|  |  |  |  | N=1,966 |  | N=4,929 |

#### Sample 3. Sample composition

| **WAVE 1** |  | **WAVE 2** |  | **WAVE 3** |  | **WAVE 4** |
| --- | --- | --- | --- | --- | --- | --- |
|  |  |  |  | 2011 |  |  |
|  |  |  |  | (7 prefectures) |  |  |
|  |  |  |  | N=3,847 |  |  |
|  |  |  |  |  |  | 2013 |
|  |  |  |  |  |  | (10 prefectures) |
|  |  |  |  |  |  | N=4,929 |

### Supplementary file 2. Sleep variables availability and coding within the JSTAR survey

**Availability of the variables by sweep**

| **Variable** | **2007** | **2009** | | **2011** | | **2013** | **Samples** |
| --- | --- | --- | --- | --- | --- | --- | --- |
|  |  | ***follow-up*** | ***Refresh-ment*** | ***Follow-up*** | ***Refresh-ment*** |  |  |
| Poor sleep (part of GHQ) | • | • | • | • | • | • | Sample 1  Sample 2  Sample 3 |
| Sleep time on weekdays (in hrs) |  | • | • | • | • | • | Sample 2  Sample 3 |
| Time to fall asleep |  |  |  | • |  | • | Sample 3 |
| Times waking up during the night |  |  |  | • |  | • | Sample 3 |
| Waking up in the early hours |  |  |  | • |  | • | Sample 3 |
| Waking up to urinate |  |  |  | • |  | • | Sample 3 |

**Variables coding (all available sweeps)**

| **Variable** | **JSTAR coding** | **Study’s coding** | **Variable type** |
| --- | --- | --- | --- |
| Poor sleep (part of GHQ) | 1. Not at all  2. 1~2 days  3. 3~4 days  4. 5 days or more | 0: Not at all  1: 1~2 days, 3~4 days, 5 days or more | Binary |
| Sleep time on weekdays (in hrs) | Numeric | Numeric | Numeric |
| Time to fall asleep | 1. No more than 10 minutes  2. 11-30 minutes  3. 31-59 minutes  4. 1-2 hours  5. More than 2 hours | 0. 30 minutes or less  1. More than 30 minutes | Binary |
| Times waking up during the night | 1. Hardly or never  2. 1-2 times  3. 1-2 times a week  4. 3 times or more a week  5. Almost every day" | 0.Never or one or two times a week  1. More than one or two times a week | Binary |
| Waking up in the early hours | 1. Hardly or never  2. 1-2 times  3. 1-2 times a week  4. 3 times or more a week  5. Almost every day" | 0.Never or one or two times a week  1. More than one or two times a week | Binary |
| Waking up to urinate | 1. Never  2. Once per night  3. 2-3 times per night  4. 4-5 times per night  5. 6 times or more per night | 0. Never or once per night  1. Two times per night or more | Binary |

### Supplementary file 3. Directed acyclic graph of the relationship between employment and sleep including the three sets of cofounders

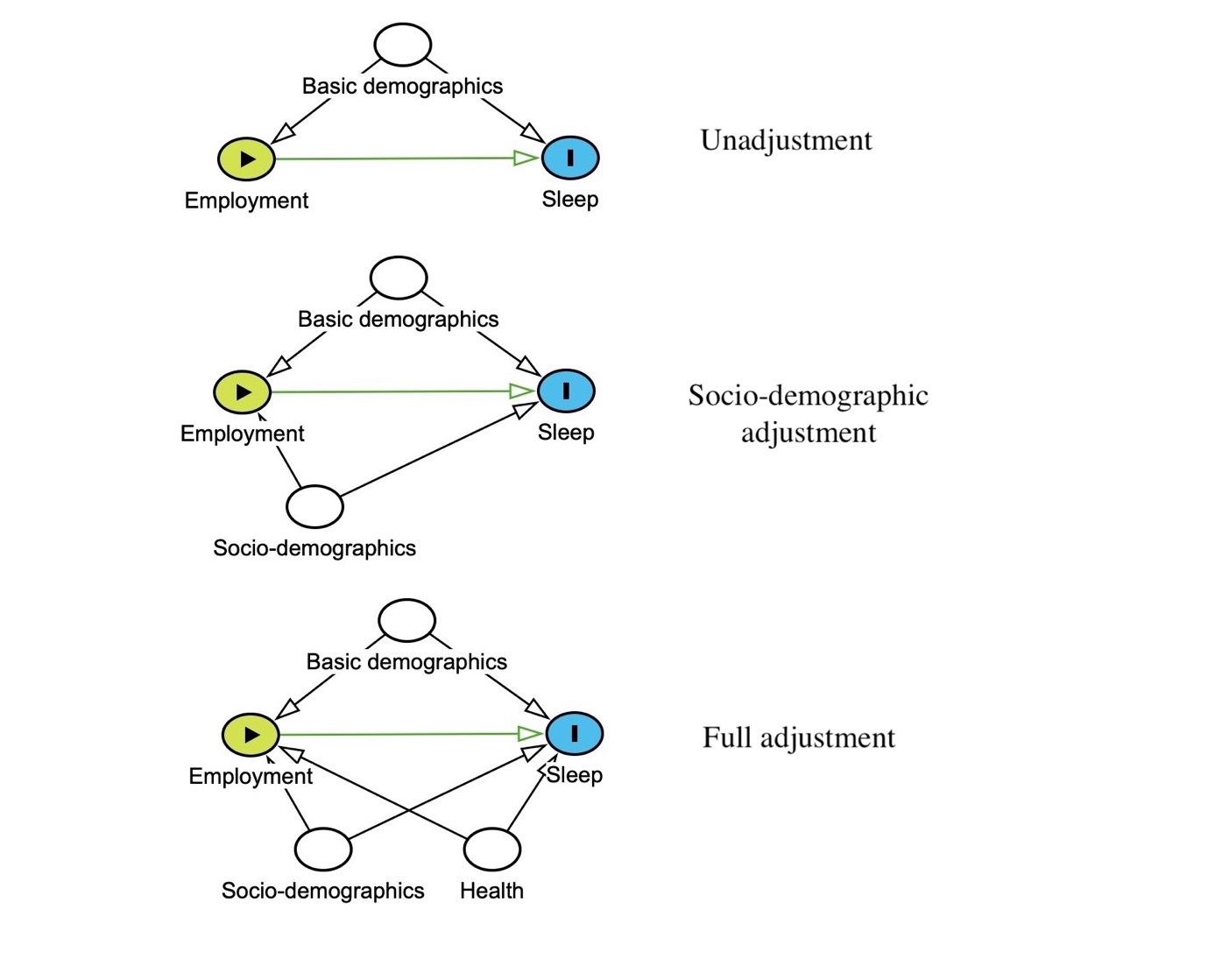

### Supplementary file 4. Descriptive statistics

#### Descriptive statistics for sample 1

| Four waves | |  |  |  |  |  |  |
| --- | --- | --- | --- | --- | --- | --- | --- |
|  |  | **2007,**  **N=3,862** | **2009,**  **N=2,718** | **2011,**  **N=3,847** | **2013,**  **N=4,929** | **2009 (refresh), N=1,440** | **2011 (refresh), N=1,966** |
| Poor sleep |  |  |  |  |  |  |  |
|  | NA | 225 | 251 | 910 | 1202 | 259 | 180 |
|  | No | 2600 (71.5%) | 1726 (69.8%) | 2107 (71.7%) | 2700 (72.4%) | 824 (69.8%) | 1173 (65.7%) |
|  | Yes | 1037 (28.5%) | 747 (30.2%) | 830 (28.3%) | 1027 (27.6%) | 357 (30.2%) | 613 (34.3%) |
| Employment |  |  |  |  |  |  |  |
|  | NA | 132 | 24 | 767 | 952 | 42 | 68 |
|  | Employed full-time | 668 (17.9%) | 389 (14.4%) | 394 (12.8%) | 438 (11.0%) | 234 (16.7%) | 347 (18.3%) |
|  | Company executive | 136 (3.6%) | 66 (2.4%) | 87 (2.8%) | 113 (2.8%) | 35 (2.5%) | 78 (4.1%) |
|  | Employed contract | 126 (3.4%) | 116 (4.3%) | 141 (4.6%) | 196 (4.9%) | 46 (3.3%) | 114 (6.0%) |
|  | Employed other | 36 (1.0%) | 32 (1.2%) | 21 (0.7%) | 62 (1.6%) | 15 (1.1%) | 8 (0.4%) |
|  | Employed part-time | 429 (11.5%) | 308 (11.4%) | 307 (10.0%) | 450 (11.3%) | 158 (11.3%) | 279 (14.7%) |
|  | Employed temporary | 24 (0.6%) | 24 (0.9%) | 12 (0.4%) | 18 (0.5%) | 4 (0.3%) | 7 (0.4%) |
|  | Help in independent business | 181 (4.9%) | 111 (4.1%) | 151 (4.9%) | 183 (4.6%) | 33 (2.4%) | 77 (4.1%) |
|  | Not retired, keep house | 831 (22.3%) | 649 (24.1%) | 798 (25.9%) | 1101 (27.7%) | 349 (25.0%) | 470 (24.8%) |
|  | Not retired, other reasons | 167 (4.5%) | 103 (3.8%) | 96 (3.1%) | 96 (2.4%) | 86 (6.2%) | 94 (5.0%) |
|  | Not retired, receiving medical care | 133 (3.6%) | 84 (3.1%) | 130 (4.2%) | 155 (3.9%) | 74 (5.3%) | 48 (2.5%) |
|  | Owner of independent business | 513 (13.8%) | 351 (13.0%) | 347 (11.3%) | 403 (10.1%) | 154 (11.0%) | 144 (7.6%) |
|  | Retired | 448 (12.0%) | 439 (16.3%) | 570 (18.5%) | 739 (18.6%) | 203 (14.5%) | 225 (11.9%) |
|  | Side job at home | 38 (1.0%) | 22 (0.8%) | 26 (0.8%) | 23 (0.6%) | 7 (0.5%) | 7 (0.4%) |
| Age |  |  |  |  |  |  |  |
|  | NA | 120 | 0 | 2 | 1 | 31 | 1 |
|  | Mean (SD) | 63.359 (7.034) | 65.559 (7.002) | 66.870 (7.183) | 67.232 (7.335) | 63.527 (7.258) | 62.455 (6.844) |
|  | Range | 50.000 - 77.000 | 52.000 - 79.000 | 53.000 - 81.000 | 49.000 - 82.000 | 51.000 - 76.000 | 47.000 - 75.000 |
| Gender |  |  |  |  |  |  |  |
|  | NA | 117 | 0 | 0 | 1 | 29 | 1 |
|  | Male | 1874 (50.0%) | 1387 (51.0%) | 1926 (50.1%) | 2378 (48.3%) | 664 (47.1%) | 877 (44.6%) |
|  | Female | 1871 (50.0%) | 1331 (49.0%) | 1921 (49.9%) | 2550 (51.7%) | 747 (52.9%) | 1088 (55.4%) |
| Highest education level | |  |  |  |  |  |  |
|  | NA | 139 | 12 | 21 | 26 | 39 | 10 |
|  | University | 495 (13.3%) | 338 (12.5%) | 551 (14.4%) | 882 (18.0%) | 240 (17.1%) | 473 (24.2%) |
|  | Elementary to middle school | 1199 (32.2%) | 901 (33.3%) | 1135 (29.7%) | 1140 (23.3%) | 317 (22.6%) | 239 (12.2%) |
|  | High School | 1601 (43.0%) | 1148 (42.4%) | 1660 (43.4%) | 2153 (43.9%) | 647 (46.2%) | 875 (44.7%) |
|  | Junior college | 151 (4.1%) | 105 (3.9%) | 188 (4.9%) | 348 (7.1%) | 97 (6.9%) | 213 (10.9%) |
|  | Vocational school | 277 (7.4%) | 214 (7.9%) | 292 (7.6%) | 380 (7.8%) | 100 (7.1%) | 156 (8.0%) |
| Marital Status |  |  |  |  |  |  |  |
|  | NA | 122 | 0 | 1 | 4 | 30 | 3 |
|  | Married or common law spouse | 3042 (81.3%) | 2226 (81.9%) | 3057 (79.5%) | 3917 (79.5%) | 1058 (75.0%) | 1527 (77.8%) |
|  | Not Married | 698 (18.7%) | 492 (18.1%) | 789 (20.5%) | 1008 (20.5%) | 352 (25.0%) | 436 (22.2%) |
| Expenses covered by friend or family | |  |  |  |  |  |  |
|  | NA | 202 | 137 | 743 | 1042 | 63 | 52 |
|  | No | 396 (10.8%) | 282 (10.9%) | 382 (12.3%) | 431 (11.1%) | 165 (12.0%) | 166 (8.7%) |
|  | Yes | 3238 (88.5%) | 2283 (88.5%) | 2699 (87.0%) | 3427 (88.2%) | 1201 (87.2%) | 1739 (90.9%) |
| Rent or own accommodation | factor(rent) |  |  |  |  |  |  |
|  | NA | 237 | 0 | 789 | 1052 | 89 | 84 |
|  | Own | 3144 (86.7%) | 132 (76.7%) | 2615 (85.5%) | 3290 (84.9%) | 992 (73.4%) | 1472 (78.2%) |
|  | Rent | 467 (12.9%) | 38 (22.1%) | 435 (14.2%) | 574 (14.8%) | 349 (25.8%) | 405 (21.5%) |
| Private health care insurance | |  |  |  |  |  |  |
|  | NA | 219 | 56 | 87 | 123 | 82 | 72 |
|  | No | 1603 (44.0%) | 1149 (43.2%) | 1754 (46.6%) | 1998 (41.6%) | 725 (53.4%) | 673 (35.5%) |
|  | Yes | 2040 (56.0%) | 1513 (56.8%) | 2006 (53.4%) | 2808 (58.4%) | 633 (46.6%) | 1221 (64.5%) |
| Self-reported health | |  |  |  |  |  |  |
|  | NA | 132 | 4 | 690 | 925 | 37 | 15 |
|  | Very good | 813 (21.8%) | 587 (21.6%) | 559 (17.7%) | 831 (20.8%) | 316 (22.5%) | 494 (25.3%) |
|  | Good | 975 (26.1%) | 626 (23.1%) | 903 (28.6%) | 1094 (27.3%) | 272 (19.4%) | 476 (24.4%) |
|  | Average | 1237 (33.2%) | 1078 (39.7%) | 1201 (38.0%) | 1509 (37.7%) | 502 (35.8%) | 688 (35.3%) |
|  | Poor | 567 (15.2%) | 354 (13.0%) | 416 (13.2%) | 460 (11.5%) | 256 (18.2%) | 216 (11.1%) |
|  | Very poor | 138 (3.7%) | 69 (2.5%) | 78 (2.5%) | 110 (2.7%) | 57 (4.1%) | 77 (3.9%) |
| Comorbidity |  |  |  |  |  |  |  |
|  | NA | 168 | 61 | 693 | 954 | 60 | 31 |
|  | No | 2787 (75.4%) | 2038 (76.7%) | 2514 (79.7%) | 3107 (78.2%) | 1043 (75.6%) | 1467 (75.8%) |
|  | Yes | 907 (24.6%) | 619 (23.3%) | 640 (20.3%) | 868 (21.8%) | 337 (24.4%) | 468 (24.2%) |
| GHQ casness |  |  |  |  |  |  |  |
|  | NA | 578 | 497 | 1183 | 1513 | 381 | 325 |
|  | No | 2297 (69.9%) | 1531 (68.9%) | 1838 (69.0%) | 2189 (64.1%) | 688 (65.0%) | 1120 (68.3%) |
|  | Yes | 987 (30.1%) | 690 (31.1%) | 826 (31.0%) | 1227 (35.9%) | 371 (35.0%) | 521 (31.7%) |
| Outpatient at clinic or hospital | |  |  |  |  |  |  |
|  | NA | 172 | 57 | 2281 | 2805 | 65 | 40 |
|  | No | 1143 (31.0%) | 640 (24.1%) | 525 (33.5%) | 576 (27.1%) | 520 (37.8%) | 549 (28.5%) |
|  | Yes | 2547 (69.0%) | 2021 (75.9%) | 1041 (66.5%) | 1548 (72.9%) | 855 (62.2%) | 1377 (71.5%) |
| Night at hospital | factor(night) |  |  |  |  |  |  |
|  | NA | 171 | 52 | 2277 | 2794 | 61 | 36 |
|  | No | 3341 (90.5%) | 2403 (90.1%) | 1404 (89.4%) | 1888 (88.4%) | 1251 (90.7%) | 1779 (92.2%) |
|  | Yes | 350 (9.5%) | 263 (9.9%) | 166 (10.6%) | 247 (11.6%) | 128 (9.3%) | 151 (7.8%) |
| Life satisfaction | |  |  |  |  |  |  |
|  | NA | 136 | 191 | 874 | 1172 | 224 | 162 |
|  | Not poor | 2962 (79.5%) | 2066 (81.8%) | 2505 (84.3%) | 3167 (84.3%) | 958 (78.8%) | 1437 (79.7%) |
|  | Poor | 764 (20.5%) | 461 (18.2%) | 468 (15.7%) | 590 (15.7%) | 258 (21.2%) | 367 (20.3%) |

#### Descriptive statistics for sample 2

| Three waves | |  |  |  |  |  |
| --- | --- | --- | --- | --- | --- | --- |
|  |  | **2009,**  **N=2,718** | **2011,**  **N=3,847** | **2013,**  **N=4,929** | **2009 (refresh), N=1,440** | **2011 (refresh), N=1,966** |
| Poor sleep |  |  |  |  |  |  |
|  | NA | 251 | 910 | 1202 | 259 | 180 |
|  | No | 1722 (69.8%) | 2107 (71.7%) | 2700 (72.4%) | 824 (69.8%) | 1173 (65.7%) |
|  | Yes | 745 (30.2%) | 830 (28.3%) | 1027 (27.6%) | 357 (30.2%) | 613 (34.3%) |
| Sleeping time during weekdays | |  |  |  |  |  |
|  | NA | 1048 | 1953 | 2506 | 690 | 737 |
|  | Mean (SD) | 7.004 (1.230) | 6.917 (1.286) | 6.782 (1.830) | 6.685 (1.204) | 6.612 (1.721) |
|  | Range | 1.000 - 15.000 | 0.000 - 13.000 | 0.000 - 70.000 | 1.000 - 12.000 | 1.000 - 42.000 |
| Employmet |  |  |  |  |  |  |
|  | NA | 24 | 767 | 952 | 42 | 68 |
|  | Employed full-time | 389 (14.4%) | 394 (12.8%) | 438 (11.0%) | 234 (16.7%) | 347 (18.3%) |
|  | Company executive | 66 (2.4%) | 87 (2.8%) | 113 (2.8%) | 35 (2.5%) | 78 (4.1%) |
|  | Employed contract | 116 (4.3%) | 141 (4.6%) | 196 (4.9%) | 46 (3.3%) | 114 (6.0%) |
|  | Employed other | 32 (1.2%) | 21 (0.7%) | 62 (1.6%) | 15 (1.1%) | 8 (0.4%) |
|  | Employed part-time | 308 (11.4%) | 307 (10.0%) | 450 (11.3%) | 158 (11.3%) | 279 (14.7%) |
|  | Employed temporary | 24 (0.9%) | 12 (0.4%) | 18 (0.5%) | 4 (0.3%) | 7 (0.4%) |
|  | Help in independent business | 111 (4.1%) | 151 (4.9%) | 183 (4.6%) | 33 (2.4%) | 77 (4.1%) |
|  | Not retired , keep house | 649 (24.1%) | 798 (25.9%) | 1101 (27.7%) | 349 (25.0%) | 470 (24.8%) |
|  | Not retired, other reasons | 103 (3.8%) | 96 (3.1%) | 96 (2.4%) | 86 (6.2%) | 94 (5.0%) |
|  | Not retired, receiving medical care | 84 (3.1%) | 130 (4.2%) | 155 (3.9%) | 74 (5.3%) | 48 (2.5%) |
|  | Owner of independent business | 351 (13.0%) | 347 (11.3%) | 403 (10.1%) | 154 (11.0%) | 144 (7.6%) |
|  | Retired | 439 (16.3%) | 570 (18.5%) | 739 (18.6%) | 203 (14.5%) | 225 (11.9%) |
|  | Side job at home | 22 (0.8%) | 26 (0.8%) | 23 (0.6%) | 7 (0.5%) | 7 (0.4%) |
| Age |  |  |  |  |  |  |
|  | NA | 0 | 2 | 1 | 31 | 1 |
|  | Mean (SD) | 65.559 (7.002) | 66.870 (7.183) | 67.232 (7.335) | 63.527 (7.258) | 62.455 (6.844) |
|  | Range | 52.000 - 79.000 | 53.000 - 81.000 | 49.000 - 82.000 | 51.000 - 76.000 | 47.000 - 75.000 |
| Gender |  |  |  |  |  |  |
|  | NA | 0 | 0 | 1 | 29 | 1 |
|  | Male | 1387 (51.0%) | 1926 (50.1%) | 2378 (48.3%) | 664 (47.1%) | 877 (44.6%) |
|  | Female | 1331 (49.0%) | 1921 (49.9%) | 2550 (51.7%) | 747 (52.9%) | 1088 (55.4%) |
| Highest education level | |  |  |  |  |  |
|  | NA | 12 | 21 | 26 | 39 | 10 |
|  | University | 338 (12.5%) | 551 (14.4%) | 882 (18.0%) | 240 (17.1%) | 473 (24.2%) |
|  | Elementary to middle school | 901 (33.3%) | 1135 (29.7%) | 1140 (23.3%) | 317 (22.6%) | 239 (12.2%) |
|  | High School | 1148 (42.4%) | 1660 (43.4%) | 2153 (43.9%) | 647 (46.2%) | 875 (44.7%) |
|  | Junior college | 105 (3.9%) | 188 (4.9%) | 348 (7.1%) | 97 (6.9%) | 213 (10.9%) |
|  | Vocational school | 214 (7.9%) | 292 (7.6%) | 380 (7.8%) | 100 (7.1%) | 156 (8.0%) |
| Marital Status |  |  |  |  |  |  |
|  | NA | 0 | 1 | 4 | 30 | 3 |
|  | Married or common law spouse | 2226 (81.9%) | 3057 (79.5%) | 3917 (79.5%) | 1058 (75.0%) | 1527 (77.8%) |
|  | Not Married | 492 (18.1%) | 789 (20.5%) | 1008 (20.5%) | 352 (25.0%) | 436 (22.2%) |
| Expenses covered by friend or family | |  |  |  |  |  |
|  | NA | 137 | 743 | 1042 | 63 | 52 |
|  | No | 282 (10.9%) | 382 (12.3%) | 431 (11.1%) | 165 (12.0%) | 166 (8.7%) |
|  | Yes | 2283 (88.5%) | 2699 (87.0%) | 3427 (88.2%) | 1201 (87.2%) | 1739 (90.9%) |
| Rent or own accommodation | factor(rent) |  |  |  |  |  |
|  | NA | 2546 | 789 | 1052 | 89 | 84 |
|  | Own | 132 (76.7%) | 2615 (85.5%) | 3290 (84.9%) | 992 (73.4%) | 1472 (78.2%) |
|  | Rent | 38 (22.1%) | 435 (14.2%) | 574 (14.8%) | 349 (25.8%) | 405 (21.5%) |
| Private health care insurance | |  |  |  |  |  |
|  | NA | 56 | 87 | 123 | 82 | 72 |
|  | No | 1149 (43.2%) | 0.534 (0.499) | 0.584 (0.493) | 0.466 (0.499) | 0.645 (0.479) |
|  | Yes | 1513 (56.8%) | 0.000 - 1.000 | 0.000 - 1.000 | 0.000 - 1.000 | 0.000 - 1.000 |
| Self-reported health | |  |  |  |  |  |
|  | NA | 4 | 690 | 925 | 37 | 15 |
|  | Very good | 587 (21.6%) | 559 (17.7%) | 831 (20.8%) | 316 (22.5%) | 494 (25.3%) |
|  | Good | 626 (23.1%) | 903 (28.6%) | 1094 (27.3%) | 272 (19.4%) | 476 (24.4%) |
|  | Average | 1078 (39.7%) | 1201 (38.0%) | 1509 (37.7%) | 502 (35.8%) | 688 (35.3%) |
|  | Poor | 354 (13.0%) | 416 (13.2%) | 460 (11.5%) | 256 (18.2%) | 216 (11.1%) |
|  | Very poor | 69 (2.5%) | 78 (2.5%) | 110 (2.7%) | 57 (4.1%) | 77 (3.9%) |
| Comorbidity |  |  |  |  |  |  |
|  | NA | 61 | 693 | 954 | 60 | 31 |
|  | No | 2038 (76.7%) | 0.203 (0.402) | 0.218 (0.413) | 0.244 (0.430) | 0.242 (0.428) |
|  | Yes | 619 (23.3%) | 0.000 - 1.000 | 0.000 - 1.000 | 0.000 - 1.000 | 0.000 - 1.000 |
| GHQ casness |  |  |  |  |  |  |
|  | NA | 497 | 1183 | 1513 | 381 | 325 |
|  | No | 1531 (68.9%) | 0.310 (0.463) | 0.359 (0.480) | 0.350 (0.477) | 0.317 (0.466) |
|  | Yes | 690 (31.1%) | 0.000 - 1.000 | 0.000 - 1.000 | 0.000 - 1.000 | 0.000 - 1.000 |
| Outpatient at clinic or hospital | |  |  |  |  |  |
|  | NA | 57 | 2281 | 2805 | 65 | 40 |
|  | No | 640 (24.1%) | 0.665 (0.472) | 0.729 (0.445) | 0.622 (0.485) | 0.715 (0.452) |
|  | Yes | 2021 (75.9%) | 0.000 - 1.000 | 0.000 - 1.000 | 0.000 - 1.000 | 0.000 - 1.000 |
| Night at hospital | factor(night) |  |  |  |  |  |
|  | NA | 52 | 2277 | 2794 | 61 | 36 |
|  | No | 2403 (90.1%) | 0.106 (0.308) | 0.116 (0.320) | 0.093 (0.290) | 0.078 (0.269) |
|  | Yes | 263 (9.9%) | 0.000 - 1.000 | 0.000 - 1.000 | 0.000 - 1.000 | 0.000 - 1.000 |
| Life satisfaction | |  |  |  |  |  |
|  | NA | 191 | 874 | 1172 | 224 | 162 |
|  | Not poor | 2066 (81.8%) | 0.157 (0.364) | 0.157 (0.364) | 0.212 (0.409) | 0.203 (0.403) |
|  | Poor | 461 (18.2%) | 0.000 - 1.000 | 0.000 - 1.000 | 0.000 - 1.000 | 0.000 - 1.000 |

#### Descriptive statistics for sample 3

| Two waves | |  |  |
| --- | --- | --- | --- |
|  |  | **2011, N=3,847** | **2013, N=4,929** |
| Poor sleep |  |  |  |
|  | NA | 910 | 1202 |
|  | No | 2107 (71.7%) | 2700 (72.4%) |
|  | Yes | 830 (28.3%) | 1027 (27.6%) |
| Sleeping time during weekdays | |  |  |
|  | NA | 1953 | 2506 |
|  | Mean (SD) | 6.917 (1.286) | 6.782 (1.830) |
|  | Range | 0.000 - 13.000 | 0.000 - 70.000 |
| Time to fall asleep | |  |  |
|  | NA | 842 | 1138 |
|  | 30 minutes or less | 2491 (82.9%) | 2813 (74.2%) |
|  | More than 30 minutes | 514 (17.1%) | 978 (25.8%) |
| Waking up at night | |  |  |
|  | NA | 849 | 1145 |
|  | Never or one or two times a week | 2281 (76.1%) | 2614 (69.1%) |
|  | More than one or two times a week | 717 (23.9%) | 1170 (30.9%) |
| Waking up in the early hours | factor(earlyhours_bi) |  |  |
|  | NA | 850 | 1139 |
|  | Never or one or two times a week | 2372 (79.1%) | 2787 (73.5%) |
|  | More than one or two times a week | 625 (20.9%) | 1003 (26.5%) |
| Night urination | factor(nighturination_bi) |  |  |
|  | NA | 850 | 1135 |
|  | 0 to 1 time per night | 2221 (74.1%) | 2962 (78.1%) |
|  | 2 times per night or more | 776 (25.9%) | 832 (21.9%) |
| Employmet |  |  |  |
|  | NA | 767 | 952 |
|  | Employed full-time | 394 (12.8%) | 438 (11.0%) |
|  | Company executive | 87 (2.8%) | 113 (2.8%) |
|  | Employed contract | 141 (4.6%) | 196 (4.9%) |
|  | Employed other | 21 (0.7%) | 62 (1.6%) |
|  | Employed part-time | 307 (10.0%) | 450 (11.3%) |
|  | Employed temporary | 12 (0.4%) | 18 (0.5%) |
|  | Help in independent business | 151 (4.9%) | 183 (4.6%) |
|  | Not retired , keep house | 798 (25.9%) | 1101 (27.7%) |
|  | Not retired, other reasons | 96 (3.1%) | 96 (2.4%) |
|  | Not retired, receiving medical care | 130 (4.2%) | 155 (3.9%) |
|  | Owner of independent business | 347 (11.3%) | 403 (10.1%) |
|  | Retired | 570 (18.5%) | 739 (18.6%) |
|  | Side job at home | 26 (0.8%) | 23 (0.6%) |
| Age |  |  |  |
|  | NA | 2 | 1 |
|  | Mean (SD) | 66.870 (7.183) | 67.232 (7.335) |
|  | Range | 53.000 - 81.000 | 49.000 - 82.000 |
| Gender |  |  |  |
|  | NA | 0 | 1 |
|  | Male | 1926 (50.1%) | 2378 (48.3%) |
|  | Female | 1921 (49.9%) | 2550 (51.7%) |
| Highest education level | |  |  |
|  | NA | 21 | 26 |
|  | University | 551 (14.4%) | 882 (18.0%) |
|  | Elementary to middle school | 1135 (29.7%) | 1140 (23.3%) |
|  | High School | 1660 (43.4%) | 2153 (43.9%) |
|  | Junior college | 188 (4.9%) | 348 (7.1%) |
|  | Vocational school | 292 (7.6%) | 380 (7.8%) |
| Marital Status |  |  |  |
|  | NA | 1 | 4 |
|  | Married or common law spouse | 3057 (79.5%) | 3917 (79.5%) |
|  | Not Married | 789 (20.5%) | 1008 (20.5%) |
| Expenses covered by friend or family | |  |  |
|  | NA | 743 | 1042 |
|  | No | 382 (12.3%) | 431 (11.1%) |
|  | Yes | 2699 (87.0%) | 3427 (88.2%) |
| Rent or own accommodation | factor(rent) |  |  |
|  | NA | 789 | 1052 |
|  | Own | 2615 (85.5%) | 3290 (84.9%) |
|  | Rent | 435 (14.2%) | 574 (14.8%) |
| Private health care insurance | |  |  |
|  | NA | 87 | 123 |
|  | No | 1754 (46.6%) | 1998 (41.6%) |
|  | Yes | 2006 (53.4%) | 2808 (58.4%) |
| Self-reported health | |  |  |
|  | NA | 690 | 925 |
|  | Very good | 559 (17.7%) | 831 (20.8%) |
|  | Good | 903 (28.6%) | 1094 (27.3%) |
|  | Average | 1201 (38.0%) | 1509 (37.7%) |
|  | Poor | 416 (13.2%) | 460 (11.5%) |
|  | Very poor | 78 (2.5%) | 110 (2.7%) |
| Comorbidity |  |  |  |
|  | NA | 693 | 954 |
|  | No | 2514 (79.7%) | 3107 (78.2%) |
|  | Yes | 640 (20.3%) | 868 (21.8%) |
| GHQ casness |  |  |  |
|  | NA | 1183 | 1513 |
|  | No | 1838 (69.0%) | 2189 (64.1%) |
|  | Yes | 826 (31.0%) | 1227 (35.9%) |
| Outpatient at clinic or hospital | |  |  |
|  | NA | 2281 | 2805 |
|  | No | 525 (33.5%) | 576 (27.1%) |
|  | Yes | 1041 (66.5%) | 1548 (72.9%) |
| Night at hospital | factor(night) |  |  |
|  | NA | 2277 | 2794 |
|  | No | 1404 (89.4%) | 1888 (88.4%) |
|  | Yes | 166 (10.6%) | 247 (11.6%) |
| Life satisfaction | |  |  |
|  | NA | 874 | 1172 |
|  | Not poor | 2505 (84.3%) | 3167 (84.3%) |
|  | Poor | 468 (15.7%) | 590 (15.7%) |

### Supplementary file 5. Model 1 results

#### Model 1 – complete case

##### Sample 1

|  |  | Sleep quality | | |
| --- | --- | --- | --- | --- |
|  |  | (Modified Poisson) | | |
|  |  | **Demo Adj.** | **Socio-eco Adj.** | **Health ad.** |
|  | (Intercept) | -1.056^***^ | -1.050^***^ | -2.364^***^ |
|  |  | (-1.370, -0.741) | (-1.413, -0.686) | (-2.821, -1.907) |
| Employment | |  |  |  |
|  | Company executive | 0.143 | **0.190^*^** | **0.191^*^** |
|  |  | (-0.056, 0.341) | **(-0.016, 0.397)** | **(-0.033, 0.414)** |
|  | Employed contract | 0.019 | 0.092 | 0.06 |
|  |  | (-0.158, 0.196) | (-0.095, 0.279) | (-0.152, 0.271) |
|  | Employed other | 0.172 | 0.131 | 0.199 |
|  |  | (-0.132, 0.477) | (-0.216, 0.478) | (-0.226, 0.623) |
|  | Employed part-time | 0.063 | 0.049 | 0.038 |
|  |  | (-0.070, 0.195) | (-0.093, 0.190) | (-0.123, 0.200) |
|  | Employed temporary | 0.216 | 0.357 | 0.398 |
|  |  | (-0.191, 0.623) | (-0.072, 0.786) | (-0.103, 0.900) |
|  | Help in independent business | 0.106 | 0.128 | 0.165 |
|  |  | (-0.072, 0.283) | (-0.062, 0.317) | (-0.064, 0.393) |
|  | Not retired – keep house | **0.181^*^** | **0.229^*^** | 0.098 |
|  |  | **(0.050, 0.311)** | **(0.091, 0.367)** | (-0.063, 0.258) |
|  | Not retired – other reasons | **0.216^*^** | **0.268^*^** | 0.071 |
|  |  | **(0.041, 0.391)** | **(0.079, 0.456)** | (-0.147, 0.289) |
|  | Not retired – receiving medical care | **0.599^*^** | **0.630^*^** | 0.037 |
|  |  | **(0.426, 0.771)** | **(0.449, 0.811)** | (-0.195, 0.269) |
|  | Owner of independent business | 0.047 | 0.029 | 0.048 |
|  |  | (-0.087, 0.181) | (-0.115, 0.174) | (-0.123, 0.218) |
|  | Retired | **0.158^*^** | **0.178^*^** | 0.048 |
|  |  | **(0.023, 0.293)** | **(0.035, 0.321)** | (-0.124, 0.220) |
|  | Side job at home | -0.134 | -0.074 | -0.078 |
|  |  | (-0.546, 0.279) | (-0.509, 0.360) | (-0.684, 0.527) |
|  | Full-time employed (Ref.) |  |  |  |
| age | | **-0.008^***^** | **-0.007^**^** | -0.001 |
|  |  | **(-0.013, -0.002)** | **(-0.013, -0.001)** | (-0.008, 0.006) |
| gender: Female | | **0.168^*^** | **0.163^*^** | **0.108^*^** |
|  |  | **(0.082, 0.253)** | **(0.070, 0.255)** | **(-0.003, 0.219)** |
| Education | |  |  |  |
|  | Elementary to middle school |  | -0.068 | -0.014 |
|  |  |  | (-0.183, 0.048) | (-0.156, 0.128) |
|  | High School |  | -0.05 | 0.021 |
|  |  |  | (-0.151, 0.050) | (-0.096, 0.138) |
|  | Junior college |  | **-0.197^**^** | -0.109 |
|  |  |  | **(-0.361, -0.033)** | (-0.301, 0.082) |
|  | Vocational school |  | 0.038 | 0.062 |
|  |  |  | (-0.103, 0.179) | (-0.106, 0.230) |
|  | University degree (Ref.) |  |  |  |
| Marital Satus | |  |  |  |
|  | Not Married and no common law spouse |  | **0.077^*^** | 0.001 |
|  |  |  | **(-0.007, 0.162)** | (-0.102, 0.104) |
| Friend or family cover expenses: Yes | |  | 0.031 | 0.046 |
|  |  |  | (-0.074, 0.136) | (-0.082, 0.175) |
| Renting accommodation: Yes | |  | **0.133^***^** | 0.003 |
|  |  |  | **(0.043, 0.224)** | (-0.108, 0.114) |
| Private health insurance: Yes | |  | 0.018 | 0.057 |
|  |  |  | (-0.051, 0.088) | (-0.030, 0.145) |
| Self-reported health | |  |  | **0.092^***^** |
|  |  |  |  | **(0.049, 0.135)** |
| Comorbidity | |  |  | 0.053 |
|  |  |  |  | (-0.044, 0.150) |
| GHQ-case | |  |  | **1.304^***^** |
|  |  |  |  | **(1.214, 1.393)** |
| Outpatient at clinic or hospital | |  |  | 0.074 |
|  |  |  |  | (-0.021, 0.170) |
| Night at hospital | |  |  | -0.002 |
|  |  |  |  | (-0.136, 0.131) |
| Life satisfaction | |  |  | **0.127^***^** |
|  |  |  |  | **(0.032, 0.221)** |
|  | nobs | 15,389 | 12,366 | 8,448 |

##### Sample 2

|  |  | Sleep quality | | | Weekdays sleep duration | | |
| --- | --- | --- | --- | --- | --- | --- | --- |
|  |  | (Modified Poisson) | | | (Linear) | | |
|  |  | **Demo Adj.** | **Socio-eco Adj.** | **Health ad.** | **Demo Adj.** | **Socio-eco Adj.** | **Health ad.** |
|  | (Intercept) | -0.905^***^ | -1.008^***^ | -2.317^***^ | 4.943^***^ | 5.226^***^ | 5.326^***^ |
|  |  | (-1.248, -0.561) | (-1.440, -0.577) | (-2.885, -1.749) | (4.563, 5.323) | (4.741, 5.711) | (4.659, 5.994) |
| Employment | |  |  |  |  |  |  |
|  | Company executive | 0.067 | 0.092 | 0.132 | **0.223^**^** | **0.334^***^** | **0.387^***^** |
|  |  | (-0.163, 0.297) | (-0.163, 0.347) | (-0.150, 0.414) | **(0.035, 0.411)** | **(0.108, 0.559)** | **(0.106, 0.669)** |
|  | Employed contract | -0.026 | 0.029 | 0.054 | 0.068 | 0.08 | 0.021 |
|  |  | (-0.219, 0.167) | (-0.185, 0.243) | (-0.188, 0.295) | (-0.082, 0.217) | (-0.102, 0.262) | (-0.206, 0.247) |
|  | Employed other | 0.209 | 0.162 | 0.143 | 0.201 | **0.316^*^** | 0.063 |
|  |  | (-0.118, 0.536) | (-0.228, 0.551) | (-0.366, 0.652) | (-0.083, 0.485) | **(-0.036, 0.668)** | (-0.473, 0.599) |
|  | Employed part-time | 0.059 | 0.034 | 0.038 | **0.161^**^** | **0.155^**^** | 0.146 |
|  |  | (-0.086, 0.204) | (-0.130, 0.199) | (-0.153, 0.230) | **(0.038, 0.285)** | **(0.005, 0.305)** | (-0.049, 0.342) |
|  | Employed temporary | 0.07 | 0.193 | 0.068 | 0.332 | 0.281 | 0.157 |
|  |  | (-0.424, 0.565) | (-0.384, 0.771) | (-0.744, 0.879) | (-0.074, 0.738) | (-0.270, 0.831) | (-0.645, 0.959) |
|  | Help in independent business | 0.097 | 0.076 | **0.258^*^** | **0.427^***^** | **0.435^***^** | **0.577^***^** |
|  |  | (-0.100, 0.294) | (-0.150, 0.303) | **(-0.031, 0.546)** | **(0.255, 0.599)** | **(0.226, 0.643)** | **(0.263, 0.892)** |
|  | Not retired – keep house | **0.143^*^** | **0.164^**^** | 0.037 | **0.303^***^** | **0.264^***^** | **0.334^***^** |
|  |  | **(-0.0003, 0.286)** | **(0.003, 0.325)** | (-0.156, 0.229) | **(0.161, 0.445)** | **(0.093, 0.434)** | **(0.105, 0.563)** |
|  | Not retired – other reasons | **0.183^*^** | **0.218^*^** | 0.006 | **0.520^***^** | **0.607^***^** | **0.951^***^** |
|  |  | **(-0.016, 0.382)** | **(-0.009, 0.445)** | (-0.268, 0.280) | **(0.279, 0.761)** | **(0.305, 0.908)** | **(0.526, 1.376)** |
|  | Not retired – receiving medical care | **0.577^***^** | **0.569^***^** | 0.008 | **0.916^***^** | **0.920^***^** | **0.537^*^** |
|  |  | **(0.385, 0.768)** | **(0.355, 0.784)** | (-0.274, 0.289) | **(0.634, 1.199)** | **(0.574, 1.266)** | **(-0.005, 1.079)** |
|  | Owner of independent business | 0.001 | -0.064 | -0.106 | **0.223^***^** | **0.212^***^** | **0.247^**^** |
|  |  | (-0.150, 0.153) | (-0.241, 0.113) | (-0.339, 0.126) | **(0.096, 0.350)** | **(0.058, 0.367)** | **(0.029, 0.465)** |
|  | Retired | **0.155^**^** | **0.156^*^** | 0.079 | **0.413^***^** | **0.514^***^** | **0.541^***^** |
|  |  | **(0.008, 0.302)** | **(-0.009, 0.322)** | (-0.129, 0.287) | **(0.243, 0.583)** | **(0.309, 0.719)** | **(0.233, 0.850)** |
|  | Side job at home | -0.026 | 0.031 | -0.01 | **0.521^***^** | **0.530^**^** | **0.887^**^** |
|  |  | (-0.481, 0.430) | (-0.479, 0.540) | (-0.900, 0.880) | **(0.131, 0.911)** | **(0.056, 1.004)** | **(0.019, 1.755)** |
|  | Full-time employed (Ref.) |  |  |  |  |  |  |
| age | | **-0.009^***^** | **-0.008^**^** | -0.003 | **0.029^***^** | **0.023^***^** | **0.021^***^** |
|  |  | **(-0.014, -0.003)** | **(-0.014, -0.001)** | (-0.011, 0.006) | **(0.023, 0.035)** | **(0.016, 0.031)** | **(0.011, 0.032)** |
| gender: Female | | **0.164^***^** | **0.165^***^** | 0.112 | **-0.423^***^** | **-0.390^***^** | **-0.374^***^** |
|  |  | **(0.071, 0.258)** | **(0.056, 0.274)** | (-0.024, 0.248) | **(-0.515, -0.332)** | **(-0.504, -0.277)** | **(-0.530, -0.218)** |
| Education | |  |  |  |  |  |  |
|  | Elementary to middle school |  | -0.033 | -0.005 |  | **0.199^**^** | 0.162 |
|  |  |  | (-0.168, 0.103) | (-0.186, 0.175) |  | **(0.046, 0.352)** | (-0.056, 0.380) |
|  | High School |  | -0.011 | 0.036 |  | 0.044 | 0.047 |
|  |  |  | (-0.126, 0.104) | (-0.103, 0.174) |  | (-0.083, 0.170) | (-0.114, 0.208) |
|  | Junior college |  | **-0.154^*^** | -0.098 |  | -0.025 | -0.072 |
|  |  |  | **(-0.335, 0.028)** | (-0.314, 0.119) |  | (-0.218, 0.168) | (-0.317, 0.173) |
|  | Vocational school |  | 0.039 | 0.039 |  | 0.151 | 0.179 |
|  |  |  | (-0.124, 0.203) | (-0.165, 0.242) |  | (-0.032, 0.334) | (-0.060, 0.418) |
|  | University degree (Ref.) |  |  |  |  |  |  |
| Marital Satus | |  |  |  |  |  |  |
|  | Not Married and no common law spouse |  | 0.034 | -0.028 |  | **-0.112^*^** | 0.009 |
|  |  |  | (-0.064, 0.132) | (-0.155, 0.098) |  | **(-0.230, 0.005)** | (-0.151, 0.168) |
| Friend or family cover expenses: Yes | |  | 0.077 | 0.127 |  | 0.028 | 0.054 |
|  |  |  | (-0.048, 0.201) | (-0.036, 0.291) |  | (-0.106, 0.161) | (-0.138, 0.247) |
| Renting accommodation: Yes | |  | **0.144^***^** | 0.027 |  | **-0.186^***^** | **-0.174^**^** |
|  |  |  | **(0.041, 0.248)** | (-0.105, 0.159) |  | **(-0.306, -0.066)** | **(-0.334, -0.014)** |
| Private health insurance: Yes | |  | 0.014 | 0.068 |  | 0.022 | 0.024 |
|  |  |  | (-0.068, 0.095) | (-0.039, 0.176) |  | (-0.072, 0.116) | (-0.106, 0.154) |
| Self-reported health | |  |  | **0.104^***^** |  |  | 0.01 |
|  |  |  |  | **(0.051, 0.158)** |  |  | (-0.052, 0.072) |
| Comorbidity | |  |  | 0.055 |  |  | 0.004 |
|  |  |  |  | (-0.066, 0.176) |  |  | (-0.151, 0.159) |
| GHQ-case | |  |  | **1.249^***^** |  |  | -0.063 |
|  |  |  |  | **(1.138, 1.360)** |  |  | (-0.190, 0.065) |
| Outpatient at clinic or hospital | |  |  | 0.076 |  |  | -0.061 |
|  |  |  |  | (-0.041, 0.194) |  |  | (-0.187, 0.065) |
| Night at hospital | |  |  | -0.003 |  |  | 0.113 |
|  |  |  |  | (-0.168, 0.161) |  |  | (-0.102, 0.329) |
| Life satisfaction | |  |  | 0.098 |  |  | -0.081 |
|  |  |  |  | (-0.019, 0.216) |  |  | (-0.241, 0.079) |
|  | nobs | 11,869 | 9,030 | 5,436 | 7,806 | 5,880 | 3,683 |

##### Sample 3 (part a)

|  |  | Sleep quality | | | Weekdays sleep duration | | | Time to fall asleep | | |
| --- | --- | --- | --- | --- | --- | --- | --- | --- | --- | --- |
|  |  | (Modified Poisson) | | | (Linear) |  |  | (Modified Poisson) | | |
|  |  | **Demo Adj.** | **Socio-eco Adj.** | **Health ad.** | **Demo Adj.** | **Socio-eco Adj.** | **Health ad.** | **Demo Adj.** | **Socio-eco Adj.** | **Health ad.** |
|  | (Intercept) | -1.112^***^ | -1.220^***^ | -2.718^***^ | 5.046^***^ | 5.189^***^ | 5.291^***^ | -2.606^***^ | -2.570^***^ | -3.433^***^ |
|  |  | (-1.586, -0.638) | (-1.769, -0.671) | (-3.534, -1.901) | (4.526, 5.565) | (4.595, 5.783) | (4.310, 6.272) | (-3.206, -2.006) | (-3.256, -1.883) | (-4.404, -2.462) |
| Employment | |  |  |  |  |  |  |  |  |  |
|  | Company executive | 0.142 | 0.12 | 0.143 | **0.259^*^** | **0.270^*^** | 0.287 | 0.181 | 0.31 | **0.369^*^** |
|  |  | (-0.185, 0.469) | (-0.218, 0.457) | (-0.246, 0.532) | **(-0.009, 0.527)** | **(-0.009, 0.549)** | (-0.099, 0.673) | (-0.190, 0.552) | (-0.065, 0.684) | **(-0.070, 0.807)** |
|  | Employed contract | 0.042 | 0.03 | -0.035 | 0.021 | 0.045 | -0.055 | -0.125 | -0.126 | -0.225 |
|  |  | (-0.232, 0.316) | (-0.253, 0.312) | (-0.381, 0.310) | (-0.192, 0.234) | (-0.179, 0.268) | (-0.363, 0.254) | (-0.461, 0.210) | (-0.475, 0.224) | (-0.677, 0.227) |
|  | Employed other | 0.267 | 0.254 | 0.183 | **0.400^**^** | **0.404^*^** | 0.017 | 0.029 | 0.016 | 0.32 |
|  |  | (-0.172, 0.706) | (-0.203, 0.712) | (-0.507, 0.874) | **(0.007, 0.792)** | **(-0.011, 0.819)** | (-0.744, 0.778) | (-0.497, 0.554) | (-0.544, 0.576) | (-0.461, 1.101) |
|  | Employed part-time | 0.123 | 0.08 | -0.014 | **0.185^**^** | **0.157^*^** | 0.091 | -0.067 | -0.079 | 0.036 |
|  |  | (-0.089, 0.335) | (-0.138, 0.298) | (-0.294, 0.266) | **(0.006, 0.363)** | **(-0.028, 0.342)** | (-0.192, 0.374) | (-0.324, 0.190) | (-0.343, 0.185) | (-0.302, 0.374) |
|  | Employed temporary | 0.308 | 0.356 | 0.128 | 0.387 | 0.263 | -0.048 | 0.158 | 0.214 | 0.77 |
|  |  | (-0.363, 0.979) | (-0.317, 1.028) | (-1.021, 1.276) | (-0.227, 1.000) | (-0.379, 0.905) | (-1.180, 1.084) | (-0.663, 0.978) | (-0.609, 1.037) | (-0.239, 1.778) |
|  | Help in independent business | 0.109 | 0.096 | 0.262 | **0.574^***^** | **0.543^***^** | **1.073^***^** | 0.025 | 0.008 | 0.124 |
|  |  | (-0.164, 0.382) | (-0.189, 0.381) | (-0.174, 0.699) | **(0.338, 0.810)** | **(0.295, 0.790)** | **(0.606, 1.541)** | (-0.292, 0.341) | (-0.326, 0.341) | (-0.407, 0.654) |
|  | Not retired – keep house | **0.257^**^** | **0.255^**^** | 0.05 | **0.347^***^** | **0.295^***^** | **0.415^**^** | **0.266^**^** | **0.286^**^** | 0.173 |
|  |  | **(0.053, 0.462)** | **(0.045, 0.466)** | (-0.226, 0.325) | **(0.146, 0.547)** | **(0.085, 0.504)** | **(0.078, 0.752)** | **(0.030, 0.501)** | **(0.043, 0.530)** | (-0.153, 0.500) |
|  | Not retired – other reasons | 0.252 | 0.201 | -0.384 | 0.291 | 0.282 | 0.385 | 0.333^*^ | 0.292 | 0.242 |
|  |  | (-0.063, 0.566) | (-0.133, 0.534) | (-0.939, 0.171) | (-0.098, 0.681) | (-0.119, 0.682) | (-0.321, 1.091) | (-0.015, 0.682) | (-0.077, 0.662) | (-0.274, 0.758) |
|  | Not retired – receiving medical care | **0.687^***^** | **0.631^***^** | 0.009 | **1.124^***^** | **1.028^***^** | **0.701^*^** | **0.716^***^** | **0.690^***^** | 0.102 |
|  |  | **(0.426, 0.948)** | **(0.358, 0.904)** | (-0.392, 0.410) | **(0.725, 1.523)** | **(0.611, 1.445)** | **(-0.111, 1.513)** | **(0.426, 1.006)** | **(0.391, 0.990)** | (-0.364, 0.568) |
|  | Owner of independent business | 0.142 | 0.094 | 0.055 | **0.276^***^** | **0.219^**^** | **0.347^**^** | 0.085 | 0.096 | -0.021 |
|  |  | (-0.072, 0.356) | (-0.126, 0.314) | (-0.276, 0.386) | **(0.097, 0.455)** | **(0.033, 0.405)** | **(0.014, 0.680)** | (-0.168, 0.339) | (-0.165, 0.356) | (-0.428, 0.386) |
|  | Retired | **0.262^**^** | **0.219^**^** | 0.081 | **0.570^***^** | **0.579^***^** | **0.618^***^** | **0.223^*^** | **0.245^*^** | 0.08 |
|  |  | **(0.056, 0.468)** | **(0.008, 0.430)** | (-0.214, 0.376) | **(0.341, 0.800)** | **(0.341, 0.817)** | **(0.203, 1.033)** | **(-0.015, 0.462)** | **(-0.0004, 0.490)** | (-0.265, 0.425) |
|  | Side job at home | 0.00001 | 0.031 | -18.813 | **0.648^**^** | **0.591^**^** | **1.324^**^** | -0.059 | -0.069 | -15.262 |
|  |  | (-0.617, 0.617) | (-0.590, 0.651) | (-373.605, 335.978) | **(0.139, 1.157)** | **(0.070, 1.113)** | **(0.074, 2.574)** | (-0.745, 0.627) | (-0.759, 0.621) | (-115.839, 85.316) |
|  | Full-time employed (Ref.) |  |  |  |  |  |  |  |  |  |
| age | | **-0.007^*^** | -0.006 | 0 | **0.027^***^** | **0.022^***^** | **0.017^**^** | **0.011^**^** | 0.005 | 0.009 |
|  |  | **(-0.014, 0.001)** | (-0.014, 0.002) | (-0.012, 0.012) | **(0.018, 0.035)** | **(0.013, 0.031)** | **(0.002, 0.032)** | **(0.002, 0.019)** | (-0.004, 0.014) | (-0.005, 0.023) |
| gender: Female | | **0.171^***^** | **0.156^**^** | 0.154 | **-0.431^***^** | **-0.394^***^** | **-0.359^***^** | **0.274^***^** | **0.203^***^** | **0.208^*^** |
|  |  | **(0.043, 0.298)** | **(0.018, 0.293)** | (-0.044, 0.353) | **(-0.554, -0.307)** | **(-0.529, -0.259)** | **(-0.582, -0.136)** | **(0.129, 0.420)** | **(0.049, 0.357)** | **(-0.023, 0.439)** |
| Education | |  |  |  |  |  |  |  |  |  |
|  | Elementary to middle school |  | -0.006 | 0.011 |  | 0.088 | 0.084 |  | **0.438^*^** | **0.429^*^** |
|  |  |  | [-0.157; 0.145] | [-0.145; 0.167] |  | [-0.039; 0.215] | [-0.044; 0.211] |  | **[ 0.210; 0.666]** | **[ 0.198; 0.659]** |
|  | High School |  | 0.009 | 0.03 |  | 0.031 | 0.028 |  | **0.323^*^** | **0.321^*^** |
|  |  |  | [-0.124; 0.142] | [-0.114; 0.174] |  | [-0.088; 0.149] | [-0.092; 0.148] |  | **[ 0.121; 0.524]** | **[ 0.120; 0.523]** |
|  | Junior college |  | -0.113 | -0.091 |  | -0.066 | -0.068 |  | 0.114 | 0.12 |
|  |  |  | [-0.326; 0.100] | [-0.311; 0.128] |  | [-0.261; 0.129] | [-0.262; 0.126] |  | [-0.171; 0.398] | [-0.163; 0.403] |
|  | Vocational school |  | 0.069 | 0.077 |  | 0.043 | 0.042 |  | **0.390^*^** | **0.381^*^** |
|  |  |  | [-0.108; 0.247] | [-0.104; 0.259] |  | [-0.124; 0.211] | [-0.125; 0.209] |  | **[ 0.107; 0.672]** | **[ 0.100; 0.663]** |
|  | University degree (Ref.) |  |  |  |  |  |  |  |  |  |
| Marital Satus | |  |  |  |  |  |  |  |  |  |
|  | Not Married and no common law spouse |  | 0.051 | -0.001 |  | **-0.210^***^** | -0.146 |  | **0.161^**^** | 0.121 |
|  |  |  | (-0.072, 0.174) | (-0.185, 0.183) |  | **(-0.350, -0.069)** | (-0.378, 0.086) |  | **(0.030, 0.291)** | (-0.081, 0.324) |
| Friend or family cover expenses: Yes | |  | 0.082 | 0.151 |  | 0.065 | 0.138 |  | 0.082 | 0.156 |
|  |  |  | (-0.070, 0.235) | (-0.079, 0.381) |  | (-0.093, 0.223) | (-0.130, 0.406) |  | (-0.083, 0.247) | (-0.104, 0.415) |
| Renting accommodation: Yes | |  | 0.101 | -0.049 |  | -0.101 | 0.02 |  | **0.126^*^** | -0.009 |
|  |  |  | (-0.037, 0.240) | (-0.257, 0.160) |  | (-0.253, 0.051) | (-0.231, 0.270) |  | **(-0.024, 0.275)** | (-0.244, 0.225) |
| Private health insurance: Yes | |  | 0.037 | 0.055 |  | 0.061 | 0.093 |  | -0.029 | **-0.167^*^** |
|  |  |  | (-0.064, 0.138) | (-0.103, 0.212) |  | (-0.050, 0.171) | (-0.093, 0.280) |  | (-0.140, 0.083) | **(-0.343, 0.010)** |
| Self-reported health | |  |  | **0.113^***^** |  |  | 0.038 |  |  | **0.183^***^** |
|  |  |  |  | **(0.034, 0.192)** |  |  | (-0.055, 0.131) |  |  | **(0.093, 0.273)** |
| Comorbidity | |  |  | 0.004 |  |  | -0.009 |  |  | 0.114 |
|  |  |  |  | (-0.175, 0.184) |  |  | (-0.241, 0.224) |  |  | (-0.082, 0.309) |
| GHQ-case | |  |  | **1.347^***^** |  |  | -0.121 |  |  | **0.432^***^** |
|  |  |  |  | **(1.182, 1.511)** |  |  | (-0.307, 0.065) |  |  | **(0.256, 0.608)** |
| Outpatient at clinic or hospital | |  |  | 0.087 |  |  | 0.02 |  |  | 0.097 |
|  |  |  |  | (-0.082, 0.257) |  |  | (-0.164, 0.205) |  |  | (-0.102, 0.296) |
| Night at hospital | |  |  | 0.009 |  |  | 0.17 |  |  | 0.128 |
|  |  |  |  | (-0.216, 0.233) |  |  | (-0.131, 0.472) |  |  | (-0.113, 0.370) |
| Life satisfaction | |  |  | **0.193^**^** |  |  | -0.148 |  |  | 0.134 |
|  |  |  |  | **(0.025, 0.361)** |  |  | (-0.391, 0.095) |  |  | (-0.079, 0.347) |
|  | nobs | 6,557 | 6,179 | 2,866 | 4,233 | 3,995 | 1,994 | 6,681 | 6,295 | 2,852 |

##### Sample 3 (part b)

|  |  | Waking up at night | | | Early hours waking up | | | Night urination | | |
| --- | --- | --- | --- | --- | --- | --- | --- | --- | --- | --- |
|  |  | (Modified Poisson) | | | (Modified Poisson) | | | (Modified Poisson) | | |
|  |  | **Demo Adj.** | **Socio-eco Adj.** | **Health ad.** | **Demo Adj.** | **Socio-eco Adj.** | **Health ad.** | **Demo Adj.** | **Socio-eco Adj.** | **Health ad.** |
|  | (Intercept) | 0.284^*^ | 0.301^*^ | 0.637^**^ | 0.263^*^ | 0.249 | 0.249 | -3.431^***^ | -3.449^***^ | -3.083^***^ |
|  |  | (-0.015, 0.583) | (-0.045, 0.646) | (0.141, 1.134) | (-0.025, 0.552) | (-0.085, 0.583) | (-0.085, 0.583) | (-3.974, -2.887) | (-4.063, -2.835) | (-3.996, -2.170) |
| Employment | |  |  |  |  |  |  |  |  |  |
|  | Company executive | -0.019 | -0.036 | -0.024 | -0.082 | -0.09 | -0.09 | -0.203 | -0.019 | 0.051 |
|  |  | (-0.204, 0.166) | (-0.229, 0.158) | (-0.252, 0.204) | (-0.272, 0.108) | (-0.288, 0.108) | (-0.288, 0.108) | (-0.602, 0.197) | (-0.423, 0.385) | (-0.429, 0.530) |
|  | Employed contract | 0.0003 | 0.001 | 0.031 | 0.004 | 0.008 | 0.008 | 0.085 | 0.179 | 0.213 |
|  |  | (-0.146, 0.147) | (-0.152, 0.155) | (-0.149, 0.212) | (-0.142, 0.151) | (-0.146, 0.161) | (-0.146, 0.161) | (-0.212, 0.382) | (-0.128, 0.486) | (-0.154, 0.580) |
|  | Employed other | -0.064 | -0.031 | -0.016 | 0.01 | 0.031 | 0.031 | -0.02 | 0.059 | 0.344 |
|  |  | (-0.338, 0.210) | (-0.316, 0.253) | (-0.459, 0.426) | (-0.254, 0.274) | (-0.244, 0.306) | (-0.244, 0.306) | (-0.544, 0.503) | (-0.500, 0.618) | (-0.436, 1.125) |
|  | Employed part-time | -0.017 | -0.004 | 0.018 | 0.058 | 0.072 | 0.072 | 0.08 | 0.17 | 0.186 |
|  |  | (-0.136, 0.103) | (-0.128, 0.120) | (-0.147, 0.183) | (-0.059, 0.175) | (-0.049, 0.193) | (-0.049, 0.193) | (-0.163, 0.324) | (-0.084, 0.423) | (-0.149, 0.520) |
|  | Employed temporary | -0.028 | -0.047 | 0.062 | 0.012 | -0.005 | -0.005 | 0.092 | 0.181 | 0.375 |
|  |  | (-0.453, 0.398) | (-0.495, 0.400) | (-0.569, 0.692) | (-0.405, 0.428) | (-0.441, 0.432) | (-0.441, 0.432) | (-0.726, 0.911) | (-0.641, 1.004) | (-0.781, 1.532) |
|  | Help in independent business | -0.035 | -0.045 | -0.016 | 0.026 | 0.02 | 0.02 | -0.058 | 0.144 | -0.048 |
|  |  | (-0.193, 0.123) | (-0.212, 0.121) | (-0.293, 0.262) | (-0.128, 0.179) | (-0.142, 0.181) | (-0.142, 0.181) | (-0.371, 0.255) | (-0.180, 0.468) | (-0.631, 0.535) |
|  | Not retired – keep house | -0.068 | -0.073 | 0.027 | -0.027 | -0.028 | -0.028 | 0.142 | **0.270^**^** | 0.229 |
|  |  | (-0.188, 0.053) | (-0.199, 0.052) | (-0.143, 0.197) | (-0.145, 0.091) | (-0.151, 0.095) | (-0.151, 0.095) | (-0.094, 0.378) | **(0.026, 0.514)** | (-0.102, 0.559) |
|  | Not retired – other reasons | -0.068 | -0.033 | 0.068 | -0.023 | -0.013 | -0.013 | **0.374^**^** | **0.327^*^** | 0.287 |
|  |  | (-0.260, 0.124) | (-0.234, 0.168) | (-0.215, 0.350) | (-0.212, 0.165) | (-0.213, 0.186) | (-0.213, 0.186) | **(0.050, 0.698)** | **(-0.016, 0.670)** | (-0.227, 0.801) |
|  | Not retired – receiving medical care | **-0.376^***^** | **-0.381^***^** | -0.202 | **-0.267^***^** | **-0.258^**^** | **-0.258^**^** | **0.414^***^** | **0.429^***^** | **0.420^*^** |
|  |  | **(-0.578, -0.173)** | **(-0.595, -0.167)** | (-0.557, 0.153) | **(-0.459, -0.074)** | **(-0.460, -0.055)** | **(-0.460, -0.055)** | **(0.119, 0.708)** | **(0.123, 0.735)** | **(-0.050, 0.891)** |
|  | Owner of independent business | -0.063 | -0.059 | 0.001 | -0.007 | -0.003 | -0.003 | 0.008 | 0.126 | 0.124 |
|  |  | (-0.184, 0.057) | (-0.184, 0.066) | (-0.192, 0.193) | (-0.126, 0.111) | (-0.126, 0.120) | (-0.126, 0.120) | (-0.235, 0.250) | (-0.126, 0.378) | (-0.264, 0.513) |
|  | Retired | -0.047 | -0.046 | -0.006 | 0.005 | 0.007 | 0.007 | **0.220^*^** | **0.282^**^** | **0.392^**^** |
|  |  | (-0.165, 0.070) | (-0.168, 0.077) | (-0.181, 0.169) | (-0.111, 0.121) | (-0.113, 0.128) | (-0.113, 0.128) | **(-0.003, 0.444)** | **(0.050, 0.515)** | **(0.072, 0.713)** |
|  | Side job at home | -0.189 | -0.195 | -0.296 | -0.086 | -0.08 | -0.08 | -0.005 | -0.052 | -0.496 |
|  |  | (-0.560, 0.183) | (-0.575, 0.185) | (-1.185, 0.593) | (-0.436, 0.265) | (-0.438, 0.278) | (-0.438, 0.278) | (-0.658, 0.648) | (-0.741, 0.638) | (-2.477, 1.484) |
|  | Full-time employed (Ref.) |  |  |  |  |  |  |  |  |  |
| age | | **-0.008^***^** | **-0.007^***^** | **-0.009^**^** | **-0.008^***^** | **-0.007^***^** | **-0.007^***^** | **0.028^***^** | **0.026^***^** | **0.012^*^** |
|  |  | **(-0.013, -0.003)** | **(-0.013, -0.002)** | **(-0.017, -0.002)** | **(-0.013, -0.003)** | **(-0.012, -0.002)** | **(-0.012, -0.002)** | **(0.019, 0.036)** | **(0.016, 0.035)** | **(-0.002, 0.025)** |
| gender: Female | | -0.013 | -0.005 | -0.041 | 0.028 | 0.034 | 0.034 | -0.027 | -0.195^***^ | -0.156 |
|  |  | (-0.091, 0.064) | (-0.089, 0.079) | (-0.163, 0.080) | (-0.047, 0.104) | (-0.047, 0.116) | (-0.047, 0.116) | (-0.167, 0.114) | (-0.343, -0.047) | (-0.382, 0.070) |
| Education | |  |  |  |  |  |  |  |  |  |
|  | Elementary to middle school |  | 0.139 | 0.134 |  | 0.137 | 0.137 |  | -0.03 | -0.032 |
|  |  |  | [-0.017; 0.296] | [-0.022; 0.290] |  | [-0.031; 0.305] | [-0.031; 0.305] |  | [-0.190; 0.129] | [-0.191; 0.128] |
|  | High School |  | 0.074 | 0.075 |  | 0.056 | 0.056 |  | -0.093 | -0.091 |
|  |  |  | [-0.070; 0.218] | [-0.069; 0.219] |  | [-0.100; 0.213] | [-0.100; 0.213] |  | [-0.244; 0.058] | [-0.242; 0.059] |
|  | Junior college |  | -0.008 | 0.003 |  | 0.003 | 0.003 |  | 0.063 | 0.064 |
|  |  |  | [-0.229; 0.214] | [-0.217; 0.222] |  | [-0.249; 0.255] | [-0.249; 0.255] |  | [-0.198; 0.325] | [-0.194; 0.323] |
|  | Vocational school |  | 0.127 | 0.121 |  | 0.091 | 0.091 |  | 0.031 | 0.027 |
|  |  |  | [-0.064; 0.318] | [-0.068; 0.310] |  | [-0.128; 0.310] | [-0.128; 0.310] |  | [-0.175; 0.237] | [-0.177; 0.232] |
|  | University degree (Ref.) |  |  |  |  |  |  |  |  |  |
| Marital Satus | |  |  |  |  |  |  |  |  |  |
|  | Not Married and no common law spouse |  | -0.007 | 0.009 |  | -0.004 | -0.004 |  | **0.728^***^** | **0.758^***^** |
|  |  |  | (-0.086, 0.072) | (-0.109, 0.127) |  | (-0.080, 0.073) | (-0.080, 0.073) |  | **(0.611, 0.845)** | **(0.576, 0.940)** |
| Friend or family cover expenses: Yes | |  | -0.038 | -0.042 |  | -0.024 | -0.024 |  | 0.075 | 0.187 |
|  |  |  | (-0.129, 0.053) | (-0.179, 0.094) |  | (-0.113, 0.065) | (-0.113, 0.065) |  | (-0.086, 0.236) | (-0.072, 0.446) |
| Renting accommodation: Yes | |  | -0.072 | -0.005 |  | -0.063 | -0.063 |  | -0.023 | -0.141 |
|  |  |  | (-0.163, 0.019) | (-0.138, 0.127) |  | (-0.151, 0.025) | (-0.151, 0.025) |  | (-0.170, 0.123) | (-0.374, 0.092) |
| Private health insurance: Yes | |  | -0.005 | -0.027 |  | 0.009 | 0.009 |  | -0.08 | **-0.160^*^** |
|  |  |  | (-0.067, 0.057) | (-0.121, 0.067) |  | (-0.052, 0.070) | (-0.052, 0.070) |  | (-0.188, 0.028) | **(-0.331, 0.012)** |
| Self-reported health | |  |  | **-0.053^**^** |  |  |  |  |  | 0.045 |
|  |  |  |  | **(-0.101, -0.005)** |  |  |  |  |  | (-0.042, 0.132) |
| Comorbidity | |  |  | 0.014 |  |  |  |  |  | 0.159 |
|  |  |  |  | (-0.103, 0.131) |  |  |  |  |  | (-0.034, 0.351) |
| GHQ-case | |  |  | **-0.209^***^** |  |  |  |  |  | **0.175^**^** |
|  |  |  |  | **(-0.309, -0.109)** |  |  |  |  |  | **(0.002, 0.348)** |
| Outpatient at clinic or hospital | |  |  | -0.037 |  |  |  |  |  | **0.225^**^** |
|  |  |  |  | (-0.134, 0.060) |  |  |  |  |  | **(0.031, 0.420)** |
| Night at hospital | |  |  | -0.025 |  |  |  |  |  | -0.114 |
|  |  |  |  | (-0.179, 0.128) |  |  |  |  |  | (-0.370, 0.142) |
| Life satisfaction | |  |  | -0.11 |  |  |  |  |  | -0.015 |
|  |  |  |  | (-0.244, 0.023) |  |  |  |  |  | (-0.237, 0.207) |
|  | nobs | 6,666 | 6,279 | 2,843 | 6,672 | 6,283 | 6,283 | 6,674 | 6,287 | 2,844 |

#### Model 1 – multiple imputations

##### Sample 1

|  |  | Sleep quality | | |
| --- | --- | --- | --- | --- |
|  |  | (Modified Poisson) | | |
|  |  | **Demo Adj.** | **Socio-eco Adj.** | **Health ad.** |
|  | (Intercept) | -1.005^*^ | -1.039^*^ | -1.683^*^ |
|  |  | [-1.315; -0.694] | [-1.351; -0.727] | [-2.033; -1.332] |
| Employment | |  |  |  |
|  | Company executive | 0.068 | 0.07 | 0.085 |
|  |  | [-0.079; 0.216] | [-0.078; 0.217] | [-0.064; 0.234] |
|  | Employed contract | 0.009 | 0.008 | 0.024 |
|  |  | [-0.143; 0.161] | [-0.146; 0.163] | [-0.123; 0.171] |
|  | Employed other | 0.056 | 0.058 | 0.059 |
|  |  | [-0.235; 0.346] | [-0.227; 0.343] | [-0.248; 0.366] |
|  | Employed part-time | 0.054 | 0.053 | 0.049 |
|  |  | [-0.058; 0.166] | [-0.061; 0.167] | [-0.064; 0.162] |
|  | Employed temporary | 0.112 | 0.113 | 0.09 |
|  |  | [-0.243; 0.466] | [-0.252; 0.477] | [-0.302; 0.482] |
|  | Help in independent business | 0.089 | 0.099 | 0.09 |
|  |  | [-0.039; 0.216] | [-0.026; 0.224] | [-0.037; 0.217] |
|  | Not retired – keep house | **0.140^*^** | **0.145^*^** | **0.123^*^** |
|  |  | **[ 0.056; 0.224]** | **[ 0.062; 0.228]** | **[ 0.041; 0.205]** |
|  | Not retired – other reasons | 0.126 | 0.124 | 0.063 |
|  |  | [-0.001; 0.254] | [-0.005; 0.254] | [-0.065; 0.192] |
|  | Not retired – receiving medical care | **0.329^*^** | **0.327^*^** | 0.114 |
|  |  | **[ 0.188; 0.470]** | **[ 0.187; 0.467]** | [-0.029; 0.257] |
|  | Owner of independent business | 0.022 | 0.027 | 0.033 |
|  |  | [-0.097; 0.140] | [-0.089; 0.142] | [-0.079; 0.146] |
|  | Retired | 0.089 | 0.092 | 0.051 |
|  |  | [-0.004; 0.183] | [-0.002; 0.186] | [-0.043; 0.144] |
|  | Side job at home | 0.056 | 0.063 | 0.091 |
|  |  | [-0.255; 0.366] | [-0.244; 0.369] | [-0.207; 0.389] |
|  | Full-time employed (Ref.) |  |  |  |
| age | | **-0.006^*^** | **-0.005^*^** | -0.003 |
|  |  | **[-0.010; -0.002]** | **[-0.009; -0.001]** | [-0.007; 0.002] |
| gender: Female | | **0.075^*^** | **0.077^*^** | **0.063^*^** |
|  |  | **[ 0.023; 0.126]** | **[ 0.019; 0.135]** | **[ 0.011; 0.116]** |
| Education | |  |  |  |
|  | Elementary to middle school |  | -0.045 | -0.033 |
|  |  |  | [-0.139; 0.049] | [-0.127; 0.060] |
|  | High School |  | -0.019 | -0.007 |
|  |  |  | [-0.119; 0.080] | [-0.102; 0.089] |
|  | Junior college |  | -0.104 | -0.088 |
|  |  |  | [-0.233; 0.025] | [-0.213; 0.036] |
|  | Vocational school |  | 0.02 | 0.023 |
|  |  |  | [-0.108; 0.148] | [-0.100; 0.146] |
|  | University degree (Ref.) |  |  |  |
| Marital Satus | |  |  |  |
|  | Not Married and no common law spouse |  | 0.03 | -0.002 |
|  |  |  | [-0.035; 0.096] | [-0.065; 0.061] |
| Friend or family cover expenses: Yes | |  | -0.019 | -0.003 |
|  |  |  | [-0.094; 0.055] | [-0.076; 0.069] |
| Renting accommodation: Yes | |  | **0.082^*^** | 0.043 |
|  |  |  | **[ 0.012; 0.152]** | [-0.028; 0.114] |
| Private health insurance: Yes | |  | 0.02 | 0.029 |
|  |  |  | [-0.028; 0.068] | [-0.019; 0.077] |
| Self-reported health | |  |  | **0.082^*^** |
|  |  |  |  | **[ 0.054; 0.109]** |
| Comorbidity | |  |  | 0.041 |
|  |  |  |  | [-0.021; 0.104] |
| GHQ-case | |  |  | **0.617^*^** |
|  |  |  |  | **[ 0.561; 0.673]** |
| Outpatient at clinic or hospital | |  |  | 0.058 |
|  |  |  |  | [-0.000; 0.115] |
| Night at hospital | |  |  | 0.037 |
|  |  |  |  | [-0.056; 0.130] |
| Life satisfaction | |  |  | **0.198^*^** |
|  |  |  |  | **[ 0.142; 0.254]** |
|  | nobs | 29072 | 29072 | 29072 |

##### Sample 2

|  |  | Sleep quality | | | Weekdays sleep duration | | |
| --- | --- | --- | --- | --- | --- | --- | --- |
|  |  | (Modified Poisson) | | | (Linear) | | |
|  |  | **Demo Adj.** | **Socio-eco Adj.** | **Health ad.** | **Demo Adj.** | **Socio-eco Adj.** | **Health ad.** |
|  | (Intercept) | -1.074^*^ | -1.161^*^ | -2.138^*^ | 5.006^*^ | 5.089^*^ | 5.113^*^ |
|  |  | [-1.497; -0.651] | [-1.655; -0.667] | [-2.609; -1.667] | [ 4.696; 5.315] | [ 4.739; 5.439] | [ 4.764; 5.462] |
| Employment | |  |  |  |  |  |  |
|  | Company executive | 0.081 | 0.089 | 0.113 | 0.195 | 0.198 | 0.199 |
|  |  | [-0.120; 0.283] | [-0.115; 0.292] | [-0.092; 0.319] | [-0.010; 0.401] | [-0.011; 0.407] | [-0.013; 0.410] |
|  | Employed contract | -0.042 | -0.041 | -0.003 | 0.048 | 0.048 | 0.046 |
|  |  | [-0.229; 0.145] | [-0.228; 0.146] | [-0.187; 0.180] | [-0.096; 0.192] | [-0.096; 0.191] | [-0.097; 0.190] |
|  | Employed other | 0.156 | 0.169 | 0.144 | 0.164 | 0.16 | 0.162 |
|  |  | [-0.207; 0.518] | [-0.192; 0.531] | [-0.221; 0.510] | [-0.119; 0.448] | [-0.124; 0.443] | [-0.123; 0.447] |
|  | Employed part-time | 0.047 | 0.043 | 0.04 | **0.155^*^** | **0.154^*^** | **0.157^*^** |
|  |  | [-0.092; 0.186] | [-0.096; 0.182] | [-0.100; 0.180] | **[ 0.026; 0.285]** | **[ 0.022; 0.285]** | **[ 0.025; 0.288]** |
|  | Employed temporary | -0.034 | -0.031 | -0.121 | 0.292 | 0.299 | 0.307 |
|  |  | [-0.485; 0.417] | [-0.482; 0.420] | [-0.573; 0.332] | [-0.129; 0.714] | [-0.122; 0.721] | [-0.111; 0.724] |
|  | Help in independent business | 0.073 | 0.094 | 0.082 | **0.431^*^** | **0.411^*^** | **0.411^*^** |
|  |  | [-0.104; 0.251] | [-0.085; 0.274] | [-0.095; 0.259] | **[ 0.253; 0.609]** | **[ 0.232; 0.590]** | **[ 0.232; 0.590]** |
|  | Not retired – keep house | **0.132^*^** | **0.142^*^** | 0.098 | **0.276^*^** | **0.273^*^** | **0.275^*^** |
|  |  | **[ 0.002; 0.262]** | **[ 0.010; 0.273]** | [-0.027; 0.224] | **[ 0.085; 0.467]** | **[ 0.081; 0.464]** | **[ 0.085; 0.466]** |
|  | Not retired – other reasons | **0.201^*^** | **0.194^*^** | 0.094 | **0.481^*^** | **0.490^*^** | **0.496^*^** |
|  |  | **[ 0.011; 0.390]** | **[ 0.004; 0.385]** | [-0.095; 0.283] | **[ 0.262; 0.701]** | **[ 0.269; 0.710]** | **[ 0.275; 0.716]** |
|  | Not retired – receiving medical care | **0.514^*^** | **0.505^*^** | 0.092 | **0.968^*^** | **0.973^*^** | **0.977^*^** |
|  |  | **[ 0.308; 0.720]** | **[ 0.298; 0.712]** | [-0.095; 0.279] | **[ 0.493; 1.443]** | **[ 0.490; 1.456]** | **[ 0.504; 1.450]** |
|  | Owner of independent business | -0.011 | -0.005 | -0.008 | **0.214^*^** | **0.204^*^** | **0.204^*^** |
|  |  | [-0.144; 0.123] | [-0.138; 0.128] | [-0.141; 0.125] | **[ 0.086; 0.342]** | **[ 0.075; 0.332]** | **[ 0.077; 0.332]** |
|  | Retired | 0.126 | 0.131 | 0.06 | **0.334^*^** | **0.338^*^** | **0.339^*^** |
|  |  | [-0.023; 0.275] | [-0.016; 0.278] | [-0.090; 0.210] | **[ 0.209; 0.459]** | **[ 0.215; 0.461]** | **[ 0.218; 0.460]** |
|  | Side job at home | -0.024 | -0.007 | 0.092 | **0.524^*^** | **0.508^*^** | **0.505^*^** |
|  |  | [-0.439; 0.391] | [-0.421; 0.406] | [-0.325; 0.510] | **[ 0.214; 0.834]** | **[ 0.198; 0.818]** | **[ 0.195; 0.815]** |
|  | Full-time employed (Ref.) |  |  |  |  |  |  |
| age | | -0.006 | -0.005 | -0.002 | **0.028^*^** | **0.026^*^** | **0.026^*^** |
|  |  | [-0.012; 0.001] | [-0.012; 0.002] | [-0.008; 0.004] | **[ 0.024; 0.033]** | **[ 0.021; 0.032]** | **[ 0.020; 0.031]** |
| gender: Female | | **0.131^*^** | **0.129^*^** | 0.092 | **-0.420^*^** | **-0.417^*^** | **-0.417^*^** |
|  |  | **[ 0.030; 0.232]** | **[ 0.028; 0.230]** | [-0.002; 0.187] | **[-0.501; -0.339]** | **[-0.507; -0.327]** | **[-0.510; -0.324]** |
| Education | |  |  |  |  |  |  |
|  | Elementary to middle school |  | -0.034 | -0.031 |  | **0.134^*^** | **0.130^*^** |
|  |  |  | [-0.152; 0.084] | [-0.147; 0.084] |  | **[ 0.026; 0.242]** | **[ 0.023; 0.237]** |
|  | High School |  | 0.007 | 0.008 |  | 0.034 | 0.032 |
|  |  |  | [-0.093; 0.107] | [-0.093; 0.109] |  | [-0.043; 0.112] | [-0.045; 0.110] |
|  | Junior college |  | -0.097 | -0.082 |  | -0.009 | -0.012 |
|  |  |  | [-0.262; 0.067] | [-0.254; 0.091] |  | [-0.133; 0.115] | [-0.136; 0.112] |
|  | Vocational school |  | 0.042 | 0.045 |  | 0.108 | 0.106 |
|  |  |  | [-0.100; 0.183] | [-0.103; 0.192] |  | [-0.060; 0.275] | [-0.063; 0.276] |
|  | University degree (Ref.) |  |  |  |  |  |  |
| Marital Satus | |  |  |  |  |  |  |
|  | Not Married and no common law spouse |  | 0.019 | -0.03 |  | -0.053 | -0.051 |
|  |  |  | [-0.057; 0.096] | [-0.110; 0.051] |  | [-0.126; 0.020] | [-0.123; 0.022] |
| Friend or family cover expenses: Yes | |  | 0.008 | 0.043 |  | 0.027 | 0.026 |
|  |  |  | [-0.094; 0.109] | [-0.057; 0.142] |  | [-0.065; 0.119] | [-0.067; 0.118] |
| Renting accommodation: Yes | |  | **0.143^*^** | 0.071 |  | -0.094 | -0.089 |
|  |  |  | **[ 0.047; 0.239]** | [-0.033; 0.175] |  | [-0.188; 0.000] | [-0.183; 0.005] |
| Private health insurance: Yes | |  | 0.012 | 0.021 |  | 0.015 | 0.015 |
|  |  |  | [-0.058; 0.082] | [-0.051; 0.093] |  | [-0.071; 0.101] | [-0.072; 0.102] |
| Self-reported health | |  |  | **0.124^*^** |  |  | 0.013 |
|  |  |  |  | **[ 0.070; 0.179]** |  |  | [-0.016; 0.042] |
| Comorbidity | |  |  | 0.01 |  |  | -0.044 |
|  |  |  |  | [-0.076; 0.096] |  |  | [-0.128; 0.039] |
| GHQ-case | |  |  | **0.917^*^** |  |  | -0.045 |
|  |  |  |  | **[ 0.764; 1.070]** |  |  | [-0.113; 0.023] |
| Outpatient at clinic or hospital | |  |  | 0.061 |  |  | -0.016 |
|  |  |  |  | [-0.017; 0.138] |  |  | [-0.077; 0.046] |
| Night at hospital | |  |  | 0.033 |  |  | 0.041 |
|  |  |  |  | [-0.073; 0.139] |  |  | [-0.070; 0.153] |
| Life satisfaction | |  |  | **0.144^*^** |  |  | -0.048 |
|  |  |  |  | **[ 0.057; 0.232]** |  |  | [-0.121; 0.024] |
|  | nobs | 16044 | 16044 | 16044 | 16044 | 16044 | 16044 |

##### Sample 3 (part a)

|  |  | Sleep quality | | | Weekdays sleep duration | | | Time to fall asleep | | |
| --- | --- | --- | --- | --- | --- | --- | --- | --- | --- | --- |
|  |  | (Modified Poisson) | | | (Linear) |  |  | (Modified Poisson) | | |
|  |  | **Demo Adj.** | **Socio-eco Adj.** | **Health ad.** | **Demo Adj.** | **Socio-eco Adj.** | **Health ad.** | **Demo Adj.** | **Socio-eco Adj.** | **Health ad.** |
|  | (Intercept) | -1.008^*^ | -1.053^*^ | -1.925^*^ | **4.963^*^** | 5.023^*^ | 5.055^*^ | -3.029^*^ | -2.970^*^ | -3.493^*^ |
|  |  | [-1.508; -0.508] | [-1.633; -0.473] | [-2.538; -1.313] | **[ 4.517; 5.409]** | [ 4.534; 5.512] | [ 4.564; 5.546] | [-3.678; -2.381] | [-3.673; -2.267] | [-4.210; -2.776] |
| Employment | |  |  |  |  |  |  |  |  |  |
|  | Company executive | 0.103 | 0.11 | 0.116 | 0.239 | 0.241 | 0.241 | 0.188 | 0.236 | 0.219 |
|  |  | [-0.180; 0.386] | [-0.173; 0.394] | [-0.185; 0.417] | [-0.018; 0.495] | [-0.022; 0.503] | [-0.021; 0.502] | [-0.207; 0.584] | [-0.157; 0.630] | [-0.194; 0.631] |
|  | Employed contract | 0.008 | 0.009 | 0.052 | 0.024 | 0.021 | 0.02 | -0.058 | -0.061 | -0.025 |
|  |  | [-0.250; 0.267] | [-0.247; 0.265] | [-0.203; 0.308] | [-0.182; 0.230] | [-0.183; 0.226] | [-0.183; 0.222] | [-0.377; 0.262] | [-0.381; 0.260] | [-0.348; 0.297] |
|  | Employed other | 0.202 | 0.213 | 0.21 | **0.401^*^** | **0.401^*^** | **0.404^*^** | 0.091 | 0.089 | 0.104 |
|  |  | [-0.225; 0.630] | [-0.214; 0.641] | [-0.233; 0.654] | **[ 0.062; 0.741]** | **[ 0.060; 0.741]** | **[ 0.063; 0.745]** | [-0.407; 0.589] | [-0.412; 0.590] | [-0.403; 0.610] |
|  | Employed part-time | 0.07 | 0.068 | 0.047 | 0.164 | 0.162 | 0.165 | -0.012 | -0.035 | -0.05 |
|  |  | [-0.116; 0.256] | [-0.123; 0.258] | [-0.145; 0.239] | [-0.039; 0.366] | [-0.042; 0.366] | [-0.038; 0.369] | [-0.270; 0.246] | [-0.295; 0.225] | [-0.311; 0.210] |
|  | Employed temporary | 0.275 | 0.282 | 0.145 | 0.334 | 0.35 | 0.36 | 0.176 | 0.158 | 0.07 |
|  |  | [-0.410; 0.960] | [-0.415; 0.978] | [-0.540; 0.829] | [-0.242; 0.910] | [-0.222; 0.921] | [-0.221; 0.940] | [-0.635; 0.987] | [-0.658; 0.975] | [-0.754; 0.894] |
|  | Help in independent business | 0.082 | 0.095 | 0.101 | **0.546^*^** | **0.529^*^** | **0.530^*^** | 0.085 | 0.087 | 0.094 |
|  |  | [-0.172; 0.336] | [-0.160; 0.351] | [-0.146; 0.349] | **[ 0.344; 0.747]** | **[ 0.325; 0.733]** | **[ 0.326; 0.733]** | [-0.215; 0.386] | [-0.215; 0.389] | [-0.208; 0.397] |
|  | Not retired – keep house | **0.221^*^** | **0.227^*^** | 0.189 | **0.344^*^** | **0.340^*^** | **0.343^*^** | **0.323^*^** | **0.327^*^** | **0.288^*^** |
|  |  | **[ 0.038; 0.405]** | **[ 0.039; 0.415]** | [-0.003; 0.382] | **[ 0.169; 0.520]** | **[ 0.160; 0.520]** | **[ 0.162; 0.524]** | **[ 0.096; 0.549]** | **[ 0.102; 0.552]** | **[ 0.062; 0.514]** |
|  | Not retired – other reasons | 0.168 | 0.167 | 0.07 | 0.311 | 0.318 | 0.325 | **0.386^*^** | **0.361^*^** | 0.29 |
|  |  | [-0.129; 0.465] | [-0.135; 0.469] | [-0.244; 0.384] | [-0.250; 0.872] | [-0.238; 0.874] | [-0.232; 0.881] | **[ 0.054; 0.718]** | **[ 0.029; 0.693]** | [-0.043; 0.622] |
|  | Not retired – receiving medical care | **0.509^*^** | **0.506^*^** | 0.137 | **1.137^*^** | **1.136^*^** | **1.142^*^** | **0.561^*^** | **0.532^*^** | 0.202 |
|  |  | **[ 0.274; 0.743]** | **[ 0.274; 0.738]** | [-0.117; 0.391] | **[ 0.388; 1.887]** | **[ 0.396; 1.877]** | **[ 0.379; 1.904]** | **[ 0.255; 0.866]** | **[ 0.222; 0.841]** | [-0.101; 0.506] |
|  | Owner of independent business | 0.088 | 0.09 | 0.09 | **0.304^*^** | **0.294^*^** | **0.294^*^** | 0.131 | 0.123 | 0.124 |
|  |  | [-0.109; 0.286] | [-0.109; 0.289] | [-0.106; 0.285] | **[ 0.150; 0.458]** | **[ 0.138; 0.449]** | **[ 0.140; 0.449]** | [-0.119; 0.380] | [-0.125; 0.372] | [-0.127; 0.374] |
|  | Retired | 0.175 | 0.181 | 0.113 | **0.502^*^** | **0.504^*^** | **0.508^*^** | 0.215 | 0.23 | 0.175 |
|  |  | [-0.007; 0.358] | [-0.003; 0.365] | [-0.072; 0.297] | **[ 0.279; 0.724]** | **[ 0.283; 0.726]** | **[ 0.288; 0.728]** | [-0.018; 0.447] | [-0.002; 0.461] | [-0.059; 0.409] |
|  | Side job at home | 0.006 | 0.018 | 0.14 | **0.618^*^** | **0.609^*^** | **0.606^*^** | 0.077 | 0.042 | 0.116 |
|  |  | [-0.563; 0.575] | [-0.555; 0.590] | [-0.441; 0.720] | **[ 0.122; 1.115]** | **[ 0.119; 1.099]** | **[ 0.117; 1.094]** | [-0.577; 0.731] | [-0.612; 0.695] | [-0.543; 0.775] |
|  | Full-time employed (Ref.) |  |  |  |  |  |  |  |  |  |
| age | | -0.007 | -0.007 | -0.004 | **0.028^*^** | **0.026^*^** | **0.026^*^** | **0.014^*^** | 0.008 | 0.008 |
|  |  | [-0.015; 0.001] | [-0.015; 0.001] | [-0.012; 0.005] | **[ 0.021; 0.035]** | **[ 0.018; 0.034]** | **[ 0.018; 0.033]** | **[ 0.005; 0.023]** | [-0.001; 0.017] | [-0.002; 0.017] |
| gender: Female | | 0.114 | 0.113 | 0.077 | **-0.444^*^** | **-0.430^*^** | **-0.428^*^** | **0.218^*^** | **0.158^*^** | **0.152^*^** |
|  |  | [-0.003; 0.230] | [-0.006; 0.232] | [-0.048; 0.203] | **[-0.602; -0.287]** | **[-0.593; -0.267]** | **[-0.592; -0.264]** | **[ 0.096; 0.341]** | **[ 0.031; 0.284]** | **[ 0.022; 0.283]** |
| Education | |  |  |  |  |  |  |  |  |  |
|  | Elementary to middle school |  | -0.006 | 0.011 |  | 0.088 | 0.084 |  | **0.438^*^** | **0.429^*^** |
|  |  |  | [-0.157; 0.145] | [-0.145; 0.167] |  | [-0.039; 0.215] | [-0.044; 0.211] |  | **[ 0.210; 0.666]** | **[ 0.198; 0.659]** |
|  | High School |  | 0.009 | 0.03 |  | 0.031 | 0.028 |  | **0.323^*^** | **0.321^*^** |
|  |  |  | [-0.124; 0.142] | [-0.114; 0.174] |  | [-0.088; 0.149] | [-0.092; 0.148] |  | **[ 0.121; 0.524]** | **[ 0.120; 0.523]** |
|  | Junior college |  | -0.113 | -0.091 |  | -0.066 | -0.068 |  | 0.114 | 0.12 |
|  |  |  | [-0.326; 0.100] | [-0.311; 0.128] |  | [-0.261; 0.129] | [-0.262; 0.126] |  | [-0.171; 0.398] | [-0.163; 0.403] |
|  | Vocational school |  | 0.069 | 0.077 |  | 0.043 | 0.042 |  | **0.390^*^** | **0.381^*^** |
|  |  |  | [-0.108; 0.247] | [-0.104; 0.259] |  | [-0.124; 0.211] | [-0.125; 0.209] |  | **[ 0.107; 0.672]** | **[ 0.100; 0.663]** |
|  | University degree (Ref.) |  |  |  |  |  |  |  |  |  |
| Marital Satus | |  |  |  |  |  |  |  |  |  |
|  | Not Married and no common law spouse |  | 0.03 | -0.003 |  | -0.075 | -0.073 |  | **0.129^*^** | 0.119 |
|  |  |  | [-0.079; 0.140] | [-0.112; 0.106] |  | [-0.171; 0.022] | [-0.170; 0.024] |  | **[ 0.005; 0.252]** | [-0.006; 0.243] |
| Friend or family cover expenses: Yes | |  | 0.025 | 0.036 |  | 0.028 | 0.027 |  | 0.024 | 0.035 |
|  |  |  | [-0.114; 0.164] | [-0.103; 0.175] |  | [-0.083; 0.138] | [-0.084; 0.138] |  | [-0.133; 0.181] | [-0.123; 0.193] |
| Renting accommodation: Yes | |  | 0.078 | 0.002 |  | -0.055 | -0.051 |  | 0.065 | 0.013 |
|  |  |  | [-0.077; 0.232] | [-0.148; 0.151] |  | [-0.178; 0.069] | [-0.176; 0.074] |  | [-0.083; 0.212] | [-0.134; 0.161] |
| Private health insurance: Yes | |  | 0.004 | -0.003 |  | 0.019 | 0.02 |  | 0.006 | 0.009 |
|  |  |  | [-0.090; 0.097] | [-0.107; 0.101] |  | [-0.077; 0.115] | [-0.075; 0.115] |  | [-0.101; 0.113] | [-0.098; 0.115] |
| Self-reported health | |  |  | **0.093^*^** |  |  | 0.01 |  |  | **0.129^*^** |
|  |  |  |  | **[ 0.048; 0.137]** |  |  | [-0.037; 0.057] |  |  | **[ 0.072; 0.187]** |
| Comorbidity | |  |  | -0.029 |  |  | 0.006 |  |  | 0.108 |
|  |  |  |  | [-0.143; 0.085] |  |  | [-0.119; 0.132] |  |  | [-0.027; 0.244] |
| GHQ-case | |  |  | **0.930^*^** |  |  | -0.064 |  |  | **0.441^*^** |
|  |  |  |  | **[ 0.836; 1.024]** |  |  | [-0.152; 0.024] |  |  | **[ 0.281; 0.601]** |
| Outpatient at clinic or hospital | |  |  | 0.004 |  |  | -0.01 |  |  | 0.082 |
|  |  |  |  | [-0.116; 0.124] |  |  | [-0.106; 0.086] |  |  | [-0.076; 0.240] |
| Night at hospital | |  |  | 0.016 |  |  | 0.011 |  |  | 0.019 |
|  |  |  |  | [-0.143; 0.176] |  |  | [-0.135; 0.157] |  |  | [-0.184; 0.221] |
| Life satisfaction | |  |  | **0.198^*^** |  |  | -0.026 |  |  | **0.143^*^** |
|  |  |  |  | **[ 0.085; 0.310]** |  |  | [-0.140; 0.088] |  |  | **[ 0.011; 0.274]** |
|  | nobs | 8776 | 8776 | 8776 | 8776 | 8776 | 8776 | 8776 | 8776 | 8776 |

##### Sample 3 (part b)

|  |  | Waking up at night | | | Early hours waking up | | | Night urination | | |
| --- | --- | --- | --- | --- | --- | --- | --- | --- | --- | --- |
|  |  | (Modified Poisson) | | | (Modified Poisson) | | | (Modified Poisson) | | |
|  |  | **Demo Adj.** | **Socio-eco Adj.** | **Health ad.** | **Demo Adj.** | **Socio-eco Adj.** | **Health ad.** | **Demo Adj.** | **Socio-eco Adj.** | **Health ad.** |
|  | (Intercept) | -2.929^*^ | -2.979^*^ | -3.495^*^ | -3.148^*^ | -3.172^*^ | -3.172^*^ | -3.522^*^ | -3.559^*^ | -3.714^*^ |
|  |  | [-3.409; -2.448] | [-3.534; -2.425] | [-4.033; -2.958] | [-3.631; -2.664] | [-3.712; -2.632] | [-3.712; -2.632] | [-4.046; -2.999] | [-4.188; -2.931] | [-4.346; -3.082] |
| Employment | |  |  |  |  |  |  |  |  |  |
|  | Company executive | 0.095 | 0.116 | 0.113 | 0.196 | 0.214 | 0.214 | -0.095 | -0.038 | -0.045 |
|  |  | [-0.258; 0.449] | [-0.243; 0.475] | [-0.238; 0.463] | [-0.138; 0.530] | [-0.123; 0.552] | [-0.123; 0.552] | [-0.464; 0.274] | [-0.404; 0.328] | [-0.420; 0.330] |
|  | Employed contract | 0.004 | 0.01 | 0.038 | 0.006 | 0.013 | 0.013 | 0.058 | 0.083 | 0.096 |
|  |  | [-0.256; 0.264] | [-0.250; 0.270] | [-0.220; 0.297] | [-0.317; 0.330] | [-0.311; 0.338] | [-0.311; 0.338] | [-0.221; 0.337] | [-0.195; 0.360] | [-0.180; 0.371] |
|  | Employed other | 0.168 | 0.169 | 0.183 | -0.036 | -0.036 | -0.036 | -0.035 | 0.026 | 0.03 |
|  |  | [-0.308; 0.644] | [-0.309; 0.648] | [-0.294; 0.659] | [-0.514; 0.442] | [-0.516; 0.443] | [-0.516; 0.443] | [-0.502; 0.433] | [-0.444; 0.496] | [-0.442; 0.501] |
|  | Employed part-time | 0.08 | 0.067 | 0.057 | -0.141 | -0.15 | -0.15 | 0.057 | 0.108 | 0.103 |
|  |  | [-0.132; 0.291] | [-0.143; 0.278] | [-0.153; 0.266] | [-0.448; 0.166] | [-0.456; 0.156] | [-0.456; 0.156] | [-0.172; 0.286] | [-0.119; 0.336] | [-0.126; 0.332] |
|  | Employed temporary | 0.125 | 0.114 | 0.038 | 0.022 | 0.014 | 0.014 | 0.144 | 0.132 | 0.108 |
|  |  | [-0.564; 0.813] | [-0.574; 0.802] | [-0.644; 0.719] | [-0.862; 0.906] | [-0.872; 0.900] | [-0.872; 0.900] | [-0.608; 0.896] | [-0.630; 0.895] | [-0.649; 0.865] |
|  | Help in independent business | 0.135 | 0.148 | 0.152 | -0.043 | -0.034 | -0.034 | -0.047 | 0.061 | 0.073 |
|  |  | [-0.111; 0.382] | [-0.100; 0.395] | [-0.092; 0.395] | [-0.352; 0.266] | [-0.350; 0.282] | [-0.350; 0.282] | [-0.326; 0.231] | [-0.215; 0.338] | [-0.204; 0.351] |
|  | Not retired – keep house | **0.209^*^** | **0.216^*^** | 0.182 | 0.08 | 0.086 | 0.086 | 0.099 | 0.171 | 0.15 |
|  |  | **[ 0.011; 0.407]** | **[ 0.017; 0.414]** | [-0.015; 0.379] | [-0.170; 0.331] | [-0.171; 0.344] | [-0.171; 0.344] | [-0.144; 0.342] | [-0.068; 0.410] | [-0.091; 0.391] |
|  | Not retired – other reasons | 0.223 | 0.216 | 0.153 | 0.106 | 0.098 | 0.098 | **0.318^*^** | 0.264 | 0.238 |
|  |  | [-0.080; 0.526] | [-0.089; 0.521] | [-0.147; 0.452] | [-0.277; 0.489] | [-0.285; 0.481] | [-0.285; 0.481] | **[ 0.001; 0.635]** | [-0.052; 0.580] | [-0.083; 0.559] |
|  | Not retired – receiving medical care | **0.509^*^** | **0.500^*^** | 0.211 | **0.411^*^** | **0.398^*^** | **0.398^*^** | 0.308 | 0.275 | 0.169 |
|  |  | **[ 0.283; 0.735]** | **[ 0.273; 0.726]** | [-0.029; 0.450] | **[ 0.139; 0.684]** | **[ 0.122; 0.675]** | **[ 0.122; 0.675]** | [-0.002; 0.618] | [-0.029; 0.579] | [-0.158; 0.497] |
|  | Owner of independent business | 0.174 | 0.177 | 0.177 | 0.053 | 0.056 | 0.056 | 0.015 | 0.075 | 0.079 |
|  |  | [-0.024; 0.372] | [-0.022; 0.376] | [-0.019; 0.374] | [-0.183; 0.288] | [-0.181; 0.292] | [-0.181; 0.292] | [-0.214; 0.244] | [-0.152; 0.302] | [-0.149; 0.307] |
|  | Retired | 0.156 | 0.166 | 0.116 | 0.01 | 0.02 | 0.02 | 0.178 | 0.213 | 0.192 |
|  |  | [-0.041; 0.353] | [-0.031; 0.362] | [-0.081; 0.313] | [-0.235; 0.255] | [-0.228; 0.269] | [-0.228; 0.269] | [-0.041; 0.396] | [-0.003; 0.430] | [-0.026; 0.410] |
|  | Side job at home | 0.389 | 0.387 | 0.454 | 0.25 | 0.244 | 0.244 | -0.045 | -0.016 | 0.013 |
|  |  | [-0.088; 0.866] | [-0.089; 0.864] | [-0.021; 0.929] | [-0.294; 0.793] | [-0.303; 0.791] | [-0.303; 0.791] | [-0.722; 0.633] | [-0.668; 0.635] | [-0.630; 0.657] |
|  | Full-time employed (Ref.) |  |  |  |  |  |  |  |  |  |
| age | | **0.022^*^** | **0.020^*^** | **0.021^*^** | **0.025^*^** | **0.023^*^** | **0.023^*^** | **0.029^*^** | **0.028^*^** | **0.027^*^** |
|  |  | **[ 0.015; 0.029]** | **[ 0.013; 0.028]** | **[ 0.013; 0.028]** | **[ 0.017; 0.033]** | **[ 0.015; 0.032]** | **[ 0.015; 0.032]** | **[ 0.021; 0.038]** | **[ 0.019; 0.036]** | **[ 0.018; 0.035]** |
| gender: Female | | 0.007 | -0.009 | -0.018 | -0.062 | -0.075 | -0.075 | 0.01 | **-0.139^*^** | **-0.136^*^** |
|  |  | [-0.110; 0.123] | [-0.132; 0.113] | [-0.141; 0.105] | [-0.194; 0.070] | [-0.214; 0.063] | [-0.214; 0.063] | [-0.113; 0.134] | **[-0.260; -0.019]** | **[-0.257; -0.015]** |
| Education | |  |  |  |  |  |  |  |  |  |
|  | Elementary to middle school |  | 0.139 | 0.134 |  | 0.137 | 0.137 |  | -0.03 | -0.032 |
|  |  |  | [-0.017; 0.296] | [-0.022; 0.290] |  | [-0.031; 0.305] | [-0.031; 0.305] |  | [-0.190; 0.129] | [-0.191; 0.128] |
|  | High School |  | 0.074 | 0.075 |  | 0.056 | 0.056 |  | -0.093 | -0.091 |
|  |  |  | [-0.070; 0.218] | [-0.069; 0.219] |  | [-0.100; 0.213] | [-0.100; 0.213] |  | [-0.244; 0.058] | [-0.242; 0.059] |
|  | Junior college |  | -0.008 | 0.003 |  | 0.003 | 0.003 |  | 0.063 | 0.064 |
|  |  |  | [-0.229; 0.214] | [-0.217; 0.222] |  | [-0.249; 0.255] | [-0.249; 0.255] |  | [-0.198; 0.325] | [-0.194; 0.323] |
|  | Vocational school |  | 0.127 | 0.121 |  | 0.091 | 0.091 |  | 0.031 | 0.027 |
|  |  |  | [-0.064; 0.318] | [-0.068; 0.310] |  | [-0.128; 0.310] | [-0.128; 0.310] |  | [-0.175; 0.237] | [-0.177; 0.232] |
|  | University degree (Ref.) |  |  |  |  |  |  |  |  |  |
| Marital Satus | |  |  |  |  |  |  |  |  |  |
|  | Not Married and no common law spouse |  | 0.022 | 0.006 |  | 0.019 | 0.019 |  | **0.707^*^** | **0.701^*^** |
|  |  |  | [-0.089; 0.134] | [-0.105; 0.118] |  | [-0.109; 0.148] | [-0.109; 0.148] |  | **[ 0.603; 0.811]** | **[ 0.597; 0.806]** |
| Friend or family cover expenses: Yes | |  | 0.025 | 0.036 |  | 0.048 | 0.048 |  | 0.072 | 0.077 |
|  |  |  | [-0.111; 0.162] | [-0.103; 0.175] |  | [-0.102; 0.199] | [-0.102; 0.199] |  | [-0.083; 0.227] | [-0.079; 0.232] |
| Renting accommodation: Yes | |  | **0.157^*^** | 0.11 |  | **0.138^*^** | **0.138^*^** |  | -0.018 | -0.037 |
|  |  |  | **[ 0.030; 0.283]** | [-0.017; 0.238] |  | **[ 0.012; 0.264]** | **[ 0.012; 0.264]** |  | [-0.152; 0.115] | [-0.170; 0.096] |
| Private health insurance: Yes | |  | 0.025 | 0.026 |  | 0.01 | 0.01 |  | -0.057 | -0.058 |
|  |  |  | [-0.064; 0.115] | [-0.062; 0.114] |  | [-0.092; 0.112] | [-0.092; 0.112] |  | [-0.156; 0.042] | [-0.157; 0.041] |
| Self-reported health | |  |  | **0.108^*^** |  |  |  |  |  | 0.046 |
|  |  |  |  | **[ 0.064; 0.152]** |  |  |  |  |  | [-0.002; 0.093] |
| Comorbidity | |  |  | 0.052 |  |  |  |  |  | **0.130^*^** |
|  |  |  |  | [-0.048; 0.151] |  |  |  |  |  | **[ 0.019; 0.242]** |
| GHQ-case | |  |  | **0.461^*^** |  |  |  |  |  | **0.192^*^** |
|  |  |  |  | **[ 0.376; 0.547]** |  |  |  |  |  | **[ 0.082; 0.302]** |
| Outpatient at clinic or hospital | |  |  | 0.035 |  |  |  |  |  | 0.05 |
|  |  |  |  | [-0.069; 0.138] |  |  |  |  |  | [-0.068; 0.169] |
| Night at hospital | |  |  | 0.015 |  |  |  |  |  | -0.047 |
|  |  |  |  | [-0.131; 0.161] |  |  |  |  |  | [-0.226; 0.132] |
| Life satisfaction | |  |  | **0.146^*^** |  |  |  |  |  | -0.007 |
|  |  |  |  | **[ 0.035; 0.257]** |  |  |  |  |  | [-0.134; 0.120] |
|  | nobs | 8776 | 8776 | 8776 | 8776 | 8776 | 8776 | 8776 | 8776 | 8776 |

### Supplementary file 6. Model 2 results

#### Model 2 – complete case

##### Sample 1

|  |  | Sleep quality | | |
| --- | --- | --- | --- | --- |
|  |  | (Modified Poisson) | | |
|  |  | **Demo Adj.** | **Socio-eco Adj.** | **Health ad.** |
|  | (Intercept) | -1.087^***^ | -1.236^***^ | -2.569^***^ |
|  |  | (-1.598, -0.575) | (-1.831, -0.641) | (-3.282, -1.857) |
| Employment | |  |  |  |
|  | Company executive | 0.125 | 0.161 | 0.167 |
|  |  | (-0.076, 0.326) | (-0.049, 0.371) | (-0.061, 0.394) |
|  | Employed contract | -0.012 | 0.054 | 0.022 |
|  |  | (-0.183, 0.160) | (-0.127, 0.235) | (-0.185, 0.229) |
|  | Employed other | 0.126 | 0.08 | 0.115 |
|  |  | (-0.192, 0.445) | (-0.284, 0.444) | (-0.349, 0.579) |
|  | Employed temporary | 0.181 | **0.378^*^** | 0.406 |
|  |  | (-0.224, 0.587) | **(-0.047, 0.804)** | (-0.094, 0.906) |
|  | Help in independent business | 0.06 | 0.069 | 0.039 |
|  |  | (-0.118, 0.237) | (-0.124, 0.261) | (-0.201, 0.279) |
|  | Owner of independent business | 0.029 | 0.015 | 0.031 |
|  |  | (-0.102, 0.161) | (-0.127, 0.157) | (-0.141, 0.203) |
|  | Side job at home | -0.024 | -0.021 | -0.06 |
|  |  | (-0.465, 0.417) | (-0.496, 0.455) | (-0.725, 0.605) |
|  | Employed (ref.) |  |  |  |
| Working time | |  |  |  |
|  | 21 to 30 | 0.086 | 0.078 | -0.05 |
|  |  | (-0.064, 0.237) | (-0.084, 0.239) | (-0.244, 0.143) |
|  | 41 to 50 | -0.026 | -0.027 | -0.079 |
|  |  | (-0.159, 0.107) | (-0.171, 0.117) | (-0.238, 0.080) |
|  | 51 and over | -0.028 | -0.03 | 0.002 |
|  |  | (-0.185, 0.129) | (-0.199, 0.139) | (-0.187, 0.192) |
|  | Less than 20 | **0.159^**^** | 0.126^*^ | 0.04 |
|  |  | **(0.023, 0.295)** | (-0.021, 0.273) | (-0.131, 0.212) |
|  | 21 to 40 (ref.) |  |  |  |
| Job satisfaction | |  |  |  |
|  | 2 | 0.038 | 0.036 | -0.075 |
|  |  | (-0.088, 0.163) | (-0.101, 0.173) | (-0.234, 0.084) |
|  | 3 | **0.303^***^** | **0.297^***^** | 0.004 |
|  |  | **(0.143, 0.463)** | **(0.122, 0.471)** | (-0.203, 0.210) |
|  | 4 | **0.280^***^** | **0.340^***^** | -0.056 |
|  |  | **(0.073, 0.488)** | **(0.114, 0.567)** | (-0.314, 0.201) |
|  | 1 (Ref.) |  |  |  |
| Age |  | **0.173^***^** | **0.180^***^** | **0.142^**^** |
|  |  | **(0.067, 0.279)** | **(0.062, 0.298)** | **(0.005, 0.278)** |
| gender: Female | | **0.173^***^** | **0.180^***^** | **0.142^**^** |
|  |  | **(0.067, 0.279)** | **(0.062, 0.298)** | **(0.005, 0.278)** |
| Education | |  |  |  |
|  | Elementary to middle school |  | -0.037 | 0.047 |
|  |  |  | (-0.205, 0.131) | (-0.154, 0.249) |
|  | High School |  | -0.084 | -0.006 |
|  |  |  | (-0.219, 0.051) | (-0.156, 0.143) |
|  | Junior college |  | -0.168 | -0.095 |
|  |  |  | (-0.398, 0.063) | (-0.354, 0.164) |
|  | Vocational school |  | 0.001 | 0.037 |
|  |  |  | (-0.191, 0.193) | (-0.179, 0.254) |
|  | University degree (ref.) |  |  |  |
| Marital Satus | |  |  |  |
|  | Not Married and no common law spouse |  | 0.063 | -0.04 |
|  |  |  | (-0.069, 0.194) | (-0.190, 0.111) |
| Friend or family cover expenses: Yes | |  | 0.068 | 0.09 |
|  |  |  | (-0.102, 0.238) | (-0.108, 0.288) |
| Renting accommodation: Yes | |  | 0.094 | 0.022 |
|  |  |  | (-0.040, 0.227) | (-0.132, 0.177) |
| Private health insurance: Yes | |  | 0.075 | 0.066 |
|  |  |  | (-0.033, 0.183) | (-0.062, 0.193) |
| Self-reported health | |  |  | **0.092^***^** |
|  |  |  |  | **(0.031, 0.154)** |
| Comorbidity | |  |  | **0.063** |
|  |  |  |  | **(-0.086, 0.212)** |
| GHQ-case | |  |  | **1.365^***^** |
|  |  |  |  | **(1.240, 1.490)** |
| Outpatient at clinic or hospital | |  |  | 0.043 |
|  |  |  |  | (-0.083, 0.170) |
| Night at hospital | |  |  | 0.119 |
|  |  |  |  | (-0.083, 0.321) |
| Life satisfaction | |  |  | 0.093 |
|  |  |  |  | (-0.045, 0.231) |
|  | nobs | 7,436 | 5,959 | 4,501 |

##### Sample 2

|  |  | Sleep quality | | | Weekdays sleep duration | | |
| --- | --- | --- | --- | --- | --- | --- | --- |
|  |  | (Modified Poisson) | | | (Linear) | | |
|  |  | **Demo Adj.** | **Socio-eco Adj.** | **Health ad.** | **Demo Adj.** | **Socio-eco Adj.** | **Health ad.** |
|  | (Intercept) | -0.934^***^ | -1.296^***^ | -2.755^***^ | 5.372^***^ | 5.413^***^ | 5.374^***^ |
|  |  | (-1.508, -0.360) | (-2.029, -0.564) | (-3.654, -1.857) | (4.877, 5.868) | (4.769, 6.056) | (4.510, 6.238) |
| Employment | |  |  |  |  |  |  |
|  | Company executive | 0.068 | 0.074 | 0.084 | 0.169^*^ | 0.293^**^ | 0.335^**^ |
|  |  | (-0.161, 0.297) | (-0.187, 0.334) | (-0.205, 0.373) | (-0.017, 0.355) | (0.066, 0.520) | (0.051, 0.618) |
|  | Employed contract | -0.06 | -0.003 | 0.023 | -0.02 | 0.004 | -0.029 |
|  |  | (-0.244, 0.124) | (-0.209, 0.204) | (-0.214, 0.260) | (-0.162, 0.122) | (-0.171, 0.178) | (-0.248, 0.189) |
|  | Employed other | 0.178 | 0.115 | 0.022 | 0.101 | 0.207 | -0.024 |
|  |  | (-0.154, 0.509) | (-0.284, 0.514) | (-0.521, 0.564) | (-0.190, 0.393) | (-0.154, 0.569) | (-0.583, 0.535) |
|  | Employed temporary | 0.012 | 0.204 | 0.045 | 0.183 | 0.076 | -0.021 |
|  |  | (-0.471, 0.494) | (-0.370, 0.777) | (-0.767, 0.857) | (-0.224, 0.590) | (-0.491, 0.642) | (-0.839, 0.798) |
|  | Help in independent business | 0.057 | 0.015 | 0.132 | **0.353^***^** | **0.329^***^** | **0.495^***^** |
|  |  | (-0.139, 0.253) | (-0.220, 0.250) | (-0.185, 0.449) | **(0.180, 0.526)** | **(0.117, 0.542)** | **(0.166, 0.825)** |
|  | Owner of independent business | -0.011 | -0.065 | -0.126 | **0.187^***^** | **0.163^**^** | 0.166 |
|  |  | (-0.157, 0.136) | (-0.240, 0.110) | (-0.366, 0.114) | **(0.065, 0.308)** | **(0.012, 0.313)** | (-0.055, 0.387) |
|  | Side job at home | 0.153 | 0.185 | 0.328 | 0.413^*^ | 0.449 | 0.715 |
|  |  | (-0.324, 0.631) | (-0.378, 0.749) | (-0.672, 1.327) | (-0.061, 0.888) | (-0.139, 1.037) | (-0.435, 1.864) |
|  | Employed (ref.) |  |  |  |  |  |  |
| Working time | |  |  |  |  |  |  |
|  | 21 to 30 | 0.09 | 0.079 | 0.02 | 0.097 | 0.062 | 0.106 |
|  |  | (-0.073, 0.253) | (-0.108, 0.266) | (-0.212, 0.252) | (-0.037, 0.230) | (-0.101, 0.225) | (-0.116, 0.328) |
|  | 41 to 50 | -0.014 | 0.014 | -0.041 | -0.110^*^ | -0.1 | -0.047 |
|  |  | (-0.163, 0.135) | (-0.157, 0.186) | (-0.236, 0.154) | (-0.225, 0.005) | (-0.243, 0.043) | (-0.230, 0.135) |
|  | 51 and over | -0.038 | -0.031 | 0.067 | **-0.198^***^** | **-0.148^*^** | -0.072 |
|  |  | (-0.217, 0.141) | (-0.241, 0.180) | (-0.179, 0.312) | **(-0.338, -0.059)** | **(-0.322, 0.027)** | (-0.304, 0.159) |
|  | Less than 20 | 0.136^*^ | 0.084 | 0.067 | 0.102 | 0.106 | 0.109 |
|  |  | (-0.012, 0.284) | (-0.090, 0.257) | (-0.146, 0.280) | (-0.022, 0.227) | (-0.048, 0.259) | (-0.103, 0.321) |
|  | 21 to 40 (ref.) |  |  |  |  |  |  |
| Job satisfaction | |  |  |  |  |  |  |
|  | 2 | 0.018 | 0.03 | -0.09 | -0.058 | -0.025 | -0.021 |
|  |  | (-0.121, 0.156) | (-0.132, 0.193) | (-0.289, 0.108) | (-0.165, 0.049) | (-0.157, 0.108) | (-0.201, 0.159) |
|  | 3 | **0.292^***^** | **0.273^**^** | -0.03 | **-0.221^***^** | **-0.206^**^** | -0.182 |
|  |  | **(0.116, 0.469)** | **(0.064, 0.483)** | (-0.292, 0.231) | **(-0.370, -0.073)** | **(-0.391, -0.021)** | (-0.435, 0.070) |
|  | 4 | **0.227^*^** | **0.287^**^** | -0.113 | **-0.171^*^** | -0.137 | -0.044 |
|  |  | **(-0.005, 0.459)** | **(0.011, 0.564)** | (-0.439, 0.212) | **(-0.366, 0.023)** | (-0.386, 0.112) | (-0.383, 0.295) |
|  | 1 (Ref.) |  |  |  |  |  |  |
| Age |  | **-0.009^*^** | -0.005 | 0.003 | **0.025^***^** | **0.021^***^** | **0.022^***^** |
|  |  | **(-0.018, 0.00002)** | (-0.017, 0.006) | (-0.011, 0.017) | **(0.018, 0.033)** | **(0.012, 0.031)** | **(0.008, 0.035)** |
| gender: Female | | **0.186^***^** | **0.213^***^** | **0.140^*^** | **-0.480^***^** | **-0.437^***^** | **-0.416^***^** |
|  |  | **(0.073, 0.300)** | **(0.074, 0.352)** | **(-0.025, 0.306)** | **(-0.579, -0.381)** | **(-0.565, -0.310)** | **(-0.583, -0.248)** |
| Education | |  |  |  |  |  |  |
|  | Elementary to middle school |  | -0.026 | -0.044 |  | **0.187^**^** | 0.148 |
|  |  |  | (-0.234, 0.183) | (-0.326, 0.237) |  | **(0.001, 0.374)** | (-0.113, 0.408) |
|  | High School |  | -0.048 | -0.005 |  | 0.08 | 0.067 |
|  |  |  | (-0.207, 0.110) | (-0.184, 0.174) |  | (-0.062, 0.223) | (-0.108, 0.243) |
|  | Junior college |  | -0.121 | -0.096 |  | 0.026 | -0.016 |
|  |  |  | (-0.378, 0.137) | (-0.389, 0.198) |  | (-0.211, 0.264) | (-0.308, 0.276) |
|  | Vocational school |  | -0.06 | -0.049 |  | 0.099 | 0.112 |
|  |  |  | (-0.289, 0.170) | (-0.316, 0.218) |  | (-0.112, 0.310) | (-0.154, 0.378) |
|  | University degree (ref.) |  |  |  |  |  |  |
| Marital Satus | |  |  |  |  |  |  |
|  | Not Married and no common law spouse |  | 0.021 | -0.056 |  | -0.096 | -0.034 |
|  |  |  | (-0.134, 0.175) | (-0.239, 0.126) |  | (-0.241, 0.050) | (-0.219, 0.151) |
| Friend or family cover expenses: Yes | |  | 0.097 | 0.193 |  | 0.052 | 0.073 |
|  |  |  | (-0.108, 0.302) | (-0.060, 0.447) |  | (-0.122, 0.225) | (-0.162, 0.309) |
| Renting accommodation: Yes | |  | 0.072 | 0.048 |  | -0.078 | -0.052 |
|  |  |  | (-0.083, 0.227) | (-0.135, 0.231) |  | (-0.222, 0.067) | (-0.236, 0.132) |
| Private health insurance: Yes | |  | 0.099 | 0.131 |  | 0.035 | 0.052 |
|  |  |  | (-0.032, 0.229) | (-0.031, 0.293) |  | (-0.082, 0.152) | (-0.103, 0.206) |
| Self-reported health | |  |  | **0.125^*^** |  |  | 0.002 |
|  |  |  |  | **(0.048, 0.201)** |  |  | (-0.071, 0.076) |
| Comorbidity | |  |  | 0.068 |  |  | -0.009 |
|  |  |  |  | (-0.120, 0.255) |  |  | (-0.198, 0.181) |
| GHQ-case | |  |  | **1.311^*^** |  |  | -0.083 |
|  |  |  |  | **(1.155, 1.466)** |  |  | (-0.234, 0.067) |
| Outpatient at clinic or hospital | |  |  | 0.08 |  |  | -0.012 |
|  |  |  |  | (-0.080, 0.239) |  |  | (-0.157, 0.133) |
| Night at hospital | |  |  | 0.128 |  |  | -0.027 |
|  |  |  |  | (-0.115, 0.371) |  |  | (-0.287, 0.233) |
| Life satisfaction | |  |  | 0.069 |  |  | **-0.164^*^** |
|  |  |  |  | (-0.105, 0.244) |  |  | **(-0.354, 0.025)** |
|  | nobs | 5,521 | 4,140 | 2,845 | 5,244 | 3,932 | 2,684 |

##### Sample 3 (part a)

|  |  | **Model 3** |  |  |  |  |  |  |  |  |
| --- | --- | --- | --- | --- | --- | --- | --- | --- | --- | --- |
|  |  | Sleep quality | | | Weekdays sleep duration | | | Time to fall asleep | | |
|  |  | (Modified Poisson) | | | (Linear) |  |  | (Modified Poisson) | | |
|  |  | **Demo Adj.** | **Socio-eco Adj.** | **Health ad.** | **Demo Adj.** | **Socio-eco Adj.** | **Health ad.** | **Demo Adj.** | **Socio-eco Adj.** | **Health ad.** |
|  | (Intercept) | -1.317^***^ | -1.392^***^ | -2.841^***^ | 5.288^***^ | 5.318^***^ | 5.189^***^ | -2.550^***^ | -2.433^***^ | -3.998^***^ |
|  |  | (-2.159, -0.475) | (-2.345, -0.438) | (-4.222, -1.461) | (4.545, 6.032) | (4.470, 6.166) | (3.791, 6.588) | (-3.607, -1.493) | (-3.616, -1.249) | (-5.693, -2.304) |
| Employment | |  |  |  |  |  |  |  |  |  |
|  | Company executive | 0.073 | 0.064 | 0.091 | 0.198 | 0.22 | 0.241 | 0.194 | 0.319^*^ | 0.379^*^ |
|  |  | (-0.254, 0.400) | (-0.278, 0.405) | (-0.311, 0.493) | (-0.079, 0.474) | (-0.072, 0.513) | (-0.172, 0.655) | (-0.177, 0.566) | (-0.060, 0.699) | (-0.061, 0.818) |
|  | Employed contract | -0.025 | -0.019 | -0.01 | -0.089 | -0.05 | -0.097 | -0.115 | -0.126 | -0.236 |
|  |  | (-0.285, 0.234) | (-0.288, 0.251) | (-0.351, 0.330) | (-0.301, 0.122) | (-0.274, 0.175) | (-0.420, 0.226) | (-0.435, 0.206) | (-0.464, 0.213) | (-0.678, 0.206) |
|  | Employed other | 0.139 | 0.139 | 0.022 | 0.238 | 0.263 | -0.027 | -0.109 | -0.007 | 0.226 |
|  |  | (-0.308, 0.587) | (-0.330, 0.609) | (-0.766, 0.810) | (-0.181, 0.657) | (-0.186, 0.711) | (-0.896, 0.841) | (-0.654, 0.436) | (-0.576, 0.561) | (-0.616, 1.067) |
|  | Employed temporary | 0.241 | 0.327 | 0.114 | 0.216 | 0.064 | -0.111 | 0.135 | 0.248 | 0.791 |
|  |  | (-0.422, 0.904) | (-0.340, 0.994) | (-1.043, 1.270) | (-0.437, 0.870) | (-0.628, 0.756) | (-1.323, 1.101) | (-0.674, 0.943) | (-0.568, 1.064) | (-0.220, 1.803) |
|  | Help in independent business | -0.033 | -0.022 | -0.053 | **0.459^***^** | **0.440^***^** | **1.069^***^** | 0.006 | 0.024 | 0.017 |
|  |  | (-0.315, 0.249) | (-0.318, 0.275) | (-0.591, 0.486) | **(0.209, 0.710)** | **(0.175, 0.706)** | **(0.525, 1.614)** | (-0.324, 0.335) | (-0.325, 0.372) | (-0.575, 0.609) |
|  | Owner of independent business | 0.103 | 0.064 | 0.057 | 0.209^**^ | 0.175^*^ | 0.291 | 0.151 | 0.166 | -0.074 |
|  |  | (-0.101, 0.307) | (-0.147, 0.275) | (-0.291, 0.406) | (0.030, 0.389) | (-0.013, 0.362) | (-0.078, 0.661) | (-0.094, 0.395) | (-0.084, 0.417) | (-0.515, 0.367) |
|  | Side job at home | 0.121 | 0.148 | -19.043 | 0.531 | 0.519 | 1.546 | -0.099 | -0.098 | -15.357 |
|  |  | (-0.554, 0.795) | (-0.537, 0.834) | (-520.795, 482.708) | (-0.125, 1.187) | (-0.167, 1.205) | (-0.457, 3.548) | (-0.921, 0.724) | (-0.935, 0.740) | (-212.163, 181.449) |
|  | Employed (ref.) |  |  |  |  |  |  |  |  |  |
| Working time | |  |  |  |  |  |  |  |  |  |
|  | 21 to 30 | 0.175 | 0.168 | -0.023 | 0.077 | 0.076 | 0.081 | 0.228^*^ | 0.231 | 0.114 |
|  |  | (-0.056, 0.406) | (-0.070, 0.405) | (-0.365, 0.318) | (-0.126, 0.280) | (-0.136, 0.288) | (-0.269, 0.431) | (-0.042, 0.499) | (-0.047, 0.509) | (-0.276, 0.504) |
|  | 41 to 50 | 0.118 | 0.115 | -0.012 | -0.101 | -0.11 | -0.06 | -0.065 | -0.048 | -0.105 |
|  |  | (-0.096, 0.332) | (-0.103, 0.334) | (-0.281, 0.258) | (-0.280, 0.079) | (-0.299, 0.078) | (-0.335, 0.215) | (-0.335, 0.205) | (-0.325, 0.230) | (-0.450, 0.241) |
|  | 51 and over | -0.024 | -0.078 | 0.022 | **-0.186^*^** | -0.156 | -0.115 | **-0.426^**^** | **-0.394^**^** | -0.425 |
|  |  | (-0.290, 0.241) | (-0.356, 0.200) | (-0.340, 0.384) | **(-0.403, 0.030)** | (-0.384, 0.072) | (-0.475, 0.245) | **(-0.791, -0.061)** | **(-0.768, -0.020)** | (-0.954, 0.104) |
|  | Less than 20 | 0.148 | 0.113 | 0.027 | 0.159 | 0.164 | 0.198 | **0.267^**^** | **0.279^**^** | 0.209 |
|  |  | (-0.070, 0.366) | (-0.113, 0.339) | (-0.304, 0.359) | (-0.033, 0.350) | (-0.036, 0.365) | (-0.158, 0.554) | **(0.014, 0.520)** | **(0.017, 0.542)** | (-0.171, 0.588) |
|  | 21 to 40 (ref.) |  |  |  |  |  |  |  |  |  |
| Job satisfaction | |  |  |  |  |  |  |  |  |  |
|  | 2 | 0.031 | -0.004 | -0.244^*^ | -0.031 | -0.026 | -0.006 | -0.094 | -0.164 | -0.235 |
|  |  | (-0.169, 0.232) | (-0.210, 0.202) | (-0.535, 0.046) | (-0.197, 0.135) | (-0.201, 0.149) | (-0.293, 0.280) | (-0.326, 0.138) | (-0.404, 0.076) | (-0.588, 0.118) |
|  | 3 | **0.318^**^** | **0.249^*^** | -0.194 | -0.18 | -0.179 | -0.089 | 0.095 | 0.035 | -0.063 |
|  |  | **(0.063, 0.572)** | **(-0.017, 0.515)** | (-0.575, 0.187) | (-0.410, 0.049) | (-0.421, 0.064) | (-0.483, 0.306) | (-0.219, 0.409) | (-0.287, 0.357) | (-0.526, 0.400) |
|  | 4 | 0.171 | 0.177 | -0.319 | -0.115 | -0.158 | -0.019 | 0.380^*^ | 0.27 | 0.076 |
|  |  | (-0.192, 0.533) | (-0.199, 0.554) | (-0.823, 0.185) | (-0.430, 0.201) | (-0.495, 0.178) | (-0.574, 0.537) | (-0.006, 0.766) | (-0.138, 0.678) | (-0.507, 0.660) |
|  | 1 (Ref.) |  |  |  |  |  |  |  |  |  |
| Age |  | -0.005 | -0.005 | 0.003 | **0.025^***^** | **0.021^***^** | **0.020^*^** | 0.009 | 0.003 | 0.019 |
|  |  | (-0.018, 0.008) | (-0.019, 0.010) | (-0.018, 0.024) | **(0.014, 0.036)** | **(0.008, 0.034)** | **(-0.002, 0.041)** | (-0.006, 0.024) | (-0.014, 0.020) | (-0.006, 0.044) |
| gender: Female | | **0.221^***^** | **0.211^**^** | 0.154 | **-0.455^***^** | **-0.455^***^** | **-0.430^***^** | 0.142 | 0.09 | 0.165 |
|  |  | **(0.059, 0.382)** | **(0.032, 0.390)** | (-0.085, 0.392) | **(-0.599, -0.312)** | **(-0.616, -0.294)** | **(-0.688, -0.173)** | (-0.054, 0.338) | (-0.125, 0.304) | (-0.129, 0.460) |
| Education | |  |  |  |  |  |  |  |  |  |
|  | Elementary to middle school |  | 0.047 | 0.161 |  | **0.250^**^** | 0.192 |  | **0.286^*^** | 0.113 |
|  |  |  | (-0.211, 0.305) | (-0.238, 0.560) |  | **(0.019, 0.481)** | (-0.205, 0.589) |  | **(-0.044, 0.616)** | (-0.376, 0.602) |
|  | High School |  | -0.061 | 0.034 |  | 0.145 | 0.143 |  | **0.266^*^** | 0.159 |
|  |  |  | (-0.273, 0.151) | (-0.231, 0.299) |  | (-0.040, 0.329) | (-0.125, 0.412) |  | **(-0.011, 0.542)** | (-0.189, 0.507) |
|  | Junior college |  | -0.188 | -0.125 |  | 0.07 | 0.008 |  | -0.147 | -0.185 |
|  |  |  | (-0.544, 0.169) | (-0.596, 0.347) |  | (-0.239, 0.380) | (-0.458, 0.473) |  | (-0.625, 0.331) | (-0.812, 0.441) |
|  | Vocational school |  | -0.04 | 0.061 |  | 0.184 | 0.181 |  | **0.428^**^** | **0.517^**^** |
|  |  |  | (-0.338, 0.258) | (-0.310, 0.432) |  | (-0.083, 0.451) | (-0.220, 0.583) |  | **(0.067, 0.788)** | **(0.072, 0.962)** |
|  | University degree (ref.) |  |  |  |  |  |  |  |  |  |
| Marital Satus | |  |  |  |  |  |  |  |  |  |
|  | Not Married and no common law spouse |  | 0.047 | -0.003 |  | -0.136 | -0.126 |  | **0.242^**^** | 0.178 |
|  |  |  | (-0.154, 0.248) | (-0.269, 0.262) |  | (-0.322, 0.049) | (-0.414, 0.163) |  | **(0.010, 0.474)** | (-0.139, 0.494) |
| Friend or family cover expenses: Yes | |  | 0.084 | 0.179 |  | 0.099 | 0.178 |  | 0.147 | 0.268 |
|  |  |  | (-0.167, 0.336) | (-0.166, 0.525) |  | (-0.121, 0.320) | (-0.169, 0.525) |  | (-0.162, 0.457) | (-0.177, 0.714) |
| Renting accommodation: Yes | |  | 0.082 | 0.019 |  | -0.03 | 0.054 |  | 0.043 | -0.087 |
|  |  |  | (-0.129, 0.293) | (-0.264, 0.302) |  | (-0.223, 0.164) | (-0.254, 0.363) |  | (-0.210, 0.295) | (-0.447, 0.274) |
| Private health insurance: Yes | |  | 0.059 | 0.034 |  | 0.051 | 0.111 |  | -0.084 | -0.247 |
|  |  |  | (-0.106, 0.225) | (-0.202, 0.270) |  | (-0.096, 0.198) | (-0.128, 0.349) |  | (-0.279, 0.110) | (-0.521, 0.027) |
| Self-reported health | |  |  | **0.121^**^** |  |  | 0.022 |  |  | **0.221^***^** |
|  |  |  |  | **(0.006, 0.236)** |  |  | (-0.097, 0.140) |  |  | **(0.079, 0.364)** |
| Comorbidity | |  |  | -0.042 |  |  | -0.04 |  |  | 0.047 |
|  |  |  |  | (-0.335, 0.250) |  |  | (-0.347, 0.266) |  |  | (-0.299, 0.394) |
| GHQ-case | |  |  | **1.489^***^** |  |  | -0.111 |  |  | **0.545^***^** |
|  |  |  |  | **(1.255, 1.723)** |  |  | (-0.346, 0.125) |  |  | **(0.276, 0.815)** |
| Outpatient at clinic or hospital | |  |  | 0.11 |  |  | 0.056 |  |  | 0.03 |
|  |  |  |  | (-0.121, 0.342) |  |  | (-0.169, 0.282) |  |  | (-0.254, 0.314) |
| Night at hospital | |  |  | 0.194 |  |  | 0.007 |  |  | -0.038 |
|  |  |  |  | (-0.136, 0.524) |  |  | (-0.381, 0.396) |  |  | (-0.458, 0.382) |
| Life satisfaction | |  |  | 0.127 |  |  | -0.245 |  |  | 0.123 |
|  |  |  |  | (-0.126, 0.380) |  |  | (-0.552, 0.061) |  |  | (-0.209, 0.455) |
|  | nobs | 2,904 | 2,734 | 1,537 | 2,770 | 2,611 | 1,468 | 2,938 | 2,767 | 1,532 |

##### Sample 3 (part b)

|  |  | Waking up at night | | | Early hours waking up | | | Night urination | | |
| --- | --- | --- | --- | --- | --- | --- | --- | --- | --- | --- |
|  |  | (Modified Poisson) | | | (Modified Poisson) | | | (Modified Poisson) | | |
|  |  | **Demo Adj.** | **Socio-eco Adj.** | **Health ad.** | **Demo Adj.** | **Socio-eco Adj.** | **Health ad.** | **Demo Adj.** | **Socio-eco Adj.** | **Health ad.** |
|  | (Intercept) | 0.323 | 0.307 | 0.672^*^ | 0.301 | 0.271 | 0.271 | -3.557^***^ | -3.825^***^ | -3.515^***^ |
|  |  | (-0.162, 0.808) | (-0.245, 0.859) | (-0.112, 1.456) | (-0.170, 0.772) | (-0.266, 0.807) | (-0.266, 0.807) | (-4.501, -2.613) | (-4.881, -2.769) | (-5.096, -1.934) |
| Employment | |  |  |  |  |  |  |  |  |  |
|  | Company executive | -0.024 | -0.043 | -0.038 | -0.109 | -0.117 | -0.117 | -0.304 | -0.135 | -0.062 |
|  |  | (-0.207, 0.160) | (-0.236, 0.150) | (-0.270, 0.194) | (-0.296, 0.079) | (-0.313, 0.079) | (-0.313, 0.079) | (-0.708, 0.099) | (-0.543, 0.274) | (-0.551, 0.426) |
|  | Employed contract | 0.005 | 0.004 | 0.03 | -0.028 | -0.024 | -0.024 | 0.013 | 0.043 | 0.145 |
|  |  | (-0.135, 0.145) | (-0.143, 0.151) | (-0.146, 0.207) | (-0.167, 0.112) | (-0.170, 0.122) | (-0.170, 0.122) | (-0.264, 0.291) | (-0.244, 0.331) | (-0.209, 0.499) |
|  | Employed other | -0.047 | -0.016 | 0.056 | -0.017 | -0.004 | -0.004 | -0.146 | -0.119 | 0.145 |
|  |  | (-0.327, 0.234) | (-0.305, 0.272) | (-0.398, 0.510) | (-0.285, 0.251) | (-0.282, 0.274) | (-0.282, 0.274) | (-0.674, 0.383) | (-0.685, 0.448) | (-0.695, 0.985) |
|  | Employed temporary | -0.014 | -0.043 | 0.066 | -0.018 | -0.044 | -0.044 | 0.025 | 0.028 | 0.379 |
|  |  | (-0.447, 0.418) | (-0.499, 0.413) | (-0.565, 0.696) | (-0.441, 0.404) | (-0.489, 0.400) | (-0.489, 0.400) | (-0.785, 0.836) | (-0.787, 0.843) | (-0.778, 1.536) |
|  | Help in independent business | -0.041 | -0.058 | 0.056 | -0.016 | -0.029 | -0.029 | -0.109 | 0.1 | -0.253 |
|  |  | (-0.206, 0.123) | (-0.232, 0.115) | (-0.246, 0.357) | (-0.174, 0.141) | (-0.196, 0.137) | (-0.196, 0.137) | (-0.430, 0.212) | (-0.232, 0.431) | (-0.906, 0.399) |
|  | Owner of independent business | -0.062 | -0.059 | 0.018 | -0.05 | -0.046 | -0.046 | -0.063 | 0.017 | -0.07 |
|  |  | (-0.181, 0.056) | (-0.181, 0.064) | (-0.186, 0.223) | (-0.166, 0.065) | (-0.165, 0.073) | (-0.165, 0.073) | (-0.296, 0.169) | (-0.221, 0.256) | (-0.501, 0.361) |
|  | Side job at home | -0.352 | -0.39 | -1.042 | -0.166 | -0.183 | -0.183 | -0.039 | -0.064 | -14.042 |
|  |  | (-0.851, 0.148) | (-0.910, 0.130) | (-3.014, 0.929) | (-0.605, 0.272) | (-0.637, 0.271) | (-0.637, 0.271) | (-0.805, 0.726) | (-0.896, 0.769) | (-2,283.041, 2,254.956) |
|  | Employed (ref.) |  |  |  |  |  |  |  |  |  |
| Working time | |  |  |  |  |  |  |  |  |  |
|  | 21 to 30 | -0.067 | -0.073 | -0.039 | -0.029 | -0.019 | -0.019 | 0.05 | 0.119 | 0.089 |
|  |  | (-0.204, 0.069) | (-0.213, 0.068) | (-0.235, 0.157) | (-0.161, 0.103) | (-0.155, 0.117) | (-0.155, 0.117) | (-0.213, 0.313) | (-0.149, 0.387) | (-0.299, 0.476) |
|  | 41 to 50 | -0.004 | -0.012 | -0.021 | 0.003 | -0.007 | -0.007 | -0.012 | 0.004 | -0.098 |
|  |  | (-0.122, 0.115) | (-0.135, 0.111) | (-0.173, 0.131) | (-0.114, 0.121) | (-0.129, 0.115) | (-0.129, 0.115) | (-0.256, 0.232) | (-0.247, 0.255) | (-0.429, 0.232) |
|  | 51 and over | 0.039 | 0.027 | 0.006 | 0.065 | 0.062 | 0.062 | -0.199 | -0.18 | -0.05 |
|  |  | (-0.103, 0.180) | (-0.121, 0.175) | (-0.191, 0.202) | (-0.074, 0.205) | (-0.083, 0.207) | (-0.083, 0.207) | (-0.511, 0.114) | (-0.503, 0.142) | (-0.484, 0.384) |
|  | Less than 20 | -0.045 | -0.037 | -0.026 | -0.008 | 0.002 | 0.002 | 0.049 | 0.102 | 0.122 |
|  |  | (-0.171, 0.082) | (-0.168, 0.093) | (-0.223, 0.171) | (-0.131, 0.115) | (-0.125, 0.128) | (-0.125, 0.128) | (-0.195, 0.294) | (-0.155, 0.359) | (-0.268, 0.513) |
|  | 21 to 40 (ref.) |  |  |  |  |  |  |  |  |  |
| Job satisfaction | |  |  |  |  |  |  |  |  |  |
|  | 2 | -0.026 | -0.013 | 0.036 | -0.051 | -0.031 | -0.031 | 0.05 | 0.02 | 0.225 |
|  |  | (-0.135, 0.083) | (-0.127, 0.101) | (-0.121, 0.194) | (-0.158, 0.056) | (-0.142, 0.081) | (-0.142, 0.081) | (-0.175, 0.274) | (-0.213, 0.253) | (-0.135, 0.585) |
|  | 3 | **-0.156^**^** | **-0.145^*^** | -0.014 | -0.115 | -0.103 | -0.103 | 0.045 | -0.032 | -0.298 |
|  |  | **(-0.311, -0.001)** | **(-0.308, 0.017)** | (-0.233, 0.204) | (-0.265, 0.034) | (-0.260, 0.053) | (-0.260, 0.053) | (-0.266, 0.357) | (-0.353, 0.290) | (-0.827, 0.231) |
|  | 4 | -0.157 | -0.133 | -0.082 | -0.12 | -0.078 | -0.078 | **0.373^*^** | 0.133 | 0.073 |
|  |  | (-0.375, 0.062) | (-0.365, 0.098) | (-0.409, 0.245) | (-0.329, 0.089) | (-0.298, 0.143) | (-0.298, 0.143) | **(-0.004, 0.751)** | (-0.278, 0.545) | (-0.567, 0.713) |
|  | 1 (Ref.) |  |  |  |  |  |  |  |  |  |
| Age |  | **-0.008^**^** | **-0.007^*^** | **-0.011^*^** | **-0.007^**^** | -0.007 | -0.007 | **0.030^***^** | **0.028^***^** | 0.015 |
|  |  | **(-0.015, -0.0005)** | **(-0.016, 0.001)** | **(-0.023, 0.001)** | **(-0.015, -0.0002)** | (-0.015, 0.001) | (-0.015, 0.001) | **(0.016, 0.044)** | **(0.013, 0.044)** | (-0.009, 0.039) |
| gender: Female | | 0.001 | 0.019 | -0.018 | 0.057 | 0.073 | 0.073 | -0.074 | **-0.335^***^** | -0.19 |
|  |  | (-0.093, 0.095) | (-0.084, 0.123) | (-0.161, 0.124) | (-0.034, 0.148) | (-0.027, 0.174) | (-0.027, 0.174) | (-0.262, 0.113) | **(-0.542, -0.128)** | (-0.482, 0.103) |
| Education | |  |  |  |  |  |  |  |  |  |
|  | Elementary to middle school |  | -0.051 | -0.088 |  | -0.052 | -0.052 |  | 0.124 | 0.21 |
|  |  |  | (-0.200, 0.099) | (-0.309, 0.132) |  | (-0.199, 0.095) | (-0.199, 0.095) |  | (-0.171, 0.420) | (-0.233, 0.653) |
|  | High School |  | -0.033 | -0.038 |  | -0.054 | -0.054 |  | 0.053 | 0.142 |
|  |  |  | (-0.152, 0.086) | (-0.185, 0.109) |  | (-0.170, 0.063) | (-0.170, 0.063) |  | (-0.200, 0.305) | (-0.184, 0.469) |
|  | Junior college |  | -0.032 | -0.004 |  | -0.025 | -0.025 |  | 0.202 | 0.115 |
|  |  |  | (-0.230, 0.166) | (-0.256, 0.248) |  | (-0.215, 0.165) | (-0.215, 0.165) |  | (-0.211, 0.615) | (-0.458, 0.688) |
|  | Vocational school |  | -0.046 | -0.033 |  | -0.078 | -0.078 |  | 0.24 | **0.391^*^** |
|  |  |  | (-0.218, 0.125) | (-0.251, 0.185) |  | (-0.246, 0.090) | (-0.246, 0.090) |  | (-0.106, 0.587) | **(-0.051, 0.832)** |
|  | University degree (ref.) |  |  |  |  |  |  |  |  |  |
| Marital Satus | |  |  |  |  |  |  |  |  |  |
|  | Not Married and no common law spouse |  | -0.027 | -0.012 |  | -0.019 | -0.019 |  | **0.938^***^** | **0.883^***^** |
|  |  |  | (-0.147, 0.094) | (-0.173, 0.150) |  | (-0.136, 0.098) | (-0.136, 0.098) |  | **(0.740, 1.136)** | **(0.608, 1.158)** |
| Friend or family cover expenses: Yes | |  | -0.006 | 0.004 |  | -0.003 | -0.003 |  | 0.145 | 0.192 |
|  |  |  | (-0.149, 0.137) | (-0.193, 0.200) |  | (-0.142, 0.136) | (-0.142, 0.136) |  | (-0.152, 0.442) | (-0.235, 0.619) |
| Renting accommodation: Yes | |  | -0.047 | 0.004 |  | -0.028 | -0.028 |  | 0.03 | -0.087 |
|  |  |  | (-0.173, 0.080) | (-0.167, 0.174) |  | (-0.150, 0.095) | (-0.150, 0.095) |  | (-0.205, 0.265) | (-0.427, 0.254) |
| Private health insurance: Yes | |  | -0.01 | -0.017 |  | 0.011 | 0.011 |  | -0.014 | -0.143 |
|  |  |  | (-0.104, 0.084) | (-0.147, 0.113) |  | (-0.081, 0.103) | (-0.081, 0.103) |  | (-0.199, 0.170) | (-0.407, 0.122) |
| Self-reported health | |  |  | -0.051 |  |  |  |  |  | 0.026 |
|  |  |  |  | (-0.117, 0.014) |  |  |  |  |  | (-0.112, 0.165) |
| Comorbidity | |  |  | 0.034 |  |  |  |  |  | 0.234 |
|  |  |  |  | (-0.136, 0.203) |  |  |  |  |  | (-0.087, 0.556) |
| GHQ-case | |  |  | **-0.219^***^** |  |  |  |  |  | 0.176 |
|  |  |  |  | **(-0.355, -0.084)** |  |  |  |  |  | (-0.086, 0.439) |
| Outpatient at clinic or hospital | |  |  | -0.015 |  |  |  |  |  | **0.275^*^** |
|  |  |  |  | (-0.139, 0.110) |  |  |  |  |  | **(-0.005, 0.554)** |
| Night at hospital | |  |  | 0.008 |  |  |  |  |  | -0.031 |
|  |  |  |  | (-0.209, 0.226) |  |  |  |  |  | (-0.454, 0.391) |
| Life satisfaction | |  |  | -0.066 |  |  |  |  |  | -0.059 |
|  |  |  |  | (-0.242, 0.110) |  |  |  |  |  | (-0.407, 0.289) |
|  | nobs | 2,924 | 2,754 | 1,524 | 2,927 | 2,756 | 2,756 | 2,933 | 2,762 | 1,528 |

#### Model 2 – multiple imputations

##### Sample 1

|  |  | Sleep quality | | |
| --- | --- | --- | --- | --- |
|  |  | (Modified Poisson) | | |
|  |  | **Demo Adj.** | **Socio-eco Adj.** | **Health ad.** |
|  | (Intercept) | -1.025^*^ | -1.097^*^ | -2.166^*^ |
|  |  | [-1.525; -0.524] | [-1.652; -0.543] | [-2.757; -1.575] |
| Employment | |  |  |  |
|  | Company executive | 0.156 | 0.158 | 0.145 |
|  |  | [-0.030; 0.343] | [-0.026; 0.343] | [-0.038; 0.328] |
|  | Employed contract | -0.013 | -0.015 | 0.025 |
|  |  | [-0.177; 0.151] | [-0.176; 0.147] | [-0.133; 0.183] |
|  | Employed other | 0.121 | 0.12 | 0.127 |
|  |  | [-0.168; 0.409] | [-0.170; 0.410] | [-0.152; 0.407] |
|  | Employed temporary | 0.152 | 0.15 | 0.072 |
|  |  | [-0.236; 0.540] | [-0.248; 0.547] | [-0.308; 0.452] |
|  | Help in independent business | 0.06 | 0.093 | 0.072 |
|  |  | [-0.098; 0.218] | [-0.066; 0.253] | [-0.080; 0.223] |
|  | Owner of independent business | 0.043 | 0.052 | 0.045 |
|  |  | [-0.076; 0.161] | [-0.068; 0.173] | [-0.072; 0.161] |
|  | Side job at home | -0.209 | -0.178 | -0.066 |
|  |  | [-0.600; 0.181] | [-0.566; 0.210] | [-0.443; 0.312] |
|  | Employed |  |  |  |
| Working time | |  |  |  |
|  | 21 to 30 | 0.076 | 0.081 | 0.038 |
|  |  | [-0.066; 0.218] | [-0.062; 0.223] | [-0.102; 0.178] |
|  | 41 to 50 | -0.017 | -0.024 | -0.053 |
|  |  | [-0.148; 0.113] | [-0.158; 0.110] | [-0.177; 0.071] |
|  | 51 and over | -0.03 | -0.04 | -0.019 |
|  |  | [-0.176; 0.117] | [-0.188; 0.107] | [-0.158; 0.120] |
|  | Less than 20 | 0.124 | 0.127 | 0.058 |
|  |  | [-0.020; 0.267] | [-0.016; 0.270] | [-0.077; 0.193] |
|  | 21 to 40 (Ref.) |  |  |  |
| Job satisfaction | |  |  |  |
|  | 2 | 0.019 | 0.015 | -0.113 |
|  |  | [-0.111; 0.150] | [-0.116; 0.145] | [-0.253; 0.026] |
|  | 3 | **0.234^*^** | **0.224^*^** | -0.062 |
|  |  | **[ 0.083; 0.386]** | **[ 0.071; 0.376]** | [-0.218; 0.095] |
|  | 4 | **0.240^*^** | **0.230^*^** | -0.137 |
|  |  | **[ 0.050; 0.431]** | **[ 0.040; 0.421]** | [-0.328; 0.054] |
|  | 1 (Ref.) |  |  |  |
| Age |  | **-0.008^*^** | -0.007 | -0.001 |
|  |  | **[-0.016; -0.000]** | [-0.016; 0.001] | [-0.009; 0.007] |
| gender: Female | | **0.161^*^** | **0.153^*^** | 0.078 |
|  |  | **[ 0.053; 0.268]** | **[ 0.034; 0.271]** | [-0.024; 0.179] |
| Education | |  |  |  |
|  | Elementary to middle school |  | -0.041 | 0.043 |
|  |  |  | [-0.198; 0.115] | [-0.105; 0.191] |
|  | High School |  | -0.051 | 0 |
|  |  |  | [-0.183; 0.080] | [-0.122; 0.122] |
|  | Junior college |  | -0.118 | -0.056 |
|  |  |  | [-0.326; 0.089] | [-0.252; 0.141] |
|  | Vocational school |  | 0.005 | 0.044 |
|  |  |  | [-0.169; 0.179] | [-0.121; 0.209] |
|  | University degree (Ref.) |  |  |  |
| Marital Satus | |  |  |  |
|  | Not Married and no common law spouse |  | 0.052 | -0.03 |
|  |  |  | [-0.067; 0.172] | [-0.143; 0.083] |
| Friend or family cover expenses: Yes | |  | -0.011 | 0.03 |
|  |  |  | [-0.157; 0.134] | [-0.108; 0.168] |
| Renting accommodation: Yes | |  | **0.119^*^** | 0.054 |
|  |  |  | **[ 0.001; 0.237]** | [-0.059; 0.167] |
| Private health insurance: Yes | |  | 0.063 | 0.037 |
|  |  |  | [-0.040; 0.165] | [-0.058; 0.131] |
| Self-reported health | |  |  | 0.07 |
|  |  |  |  | [-0.007; 0.147] |
| Comorbidity | |  |  | -0.02 |
|  |  |  |  | [-0.132; 0.093] |
| GHQ-case | |  |  | **1.353^*^** |
|  |  |  |  | **[ 1.249; 1.457]** |
| Outpatient at clinic or hospital | |  |  | 0.08 |
|  |  |  |  | [-0.014; 0.173] |
| Night at hospital | |  |  | 0.085 |
|  |  |  |  | [-0.070; 0.240] |
| Life satisfaction | |  |  | 0.074 |
|  |  |  |  | [-0.044; 0.193] |
|  | nobs | 8689 | 8689 | 8689 |

##### Sample 2

|  |  | Sleep quality | | | Weekdays sleep duration | | |
| --- | --- | --- | --- | --- | --- | --- | --- |
|  |  | (Modified Poisson) | | | (Linear) | | |
|  |  | **Demo Adj.** | **Socio-eco Adj.** | **Health ad.** | **Demo Adj.** | **Socio-eco Adj.** | **Health ad.** |
|  | (Intercept) | -0.943^*^ | -1.067^*^ | -2.350^*^ | 5.233^*^ | 5.243^*^ | 5.351^*^ |
|  |  | [-1.480; -0.405] | [-1.665; -0.470] | [-2.993; -1.707] | [ 4.659; 5.807] | [ 4.626; 5.861] | [ 4.724; 5.977] |
| Employment | |  |  |  |  |  |  |
|  | Company executive | 0.072 | 0.07 | 0.05 | 0.22 | 0.23 | 0.227 |
|  |  | [-0.143; 0.288] | [-0.150; 0.290] | [-0.169; 0.268] | [-0.091; 0.531] | [-0.082; 0.542] | [-0.084; 0.538] |
|  | Employed contract | -0.034 | -0.036 | 0.007 | 0.019 | 0.02 | 0.017 |
|  |  | [-0.218; 0.149] | [-0.220; 0.148] | [-0.174; 0.187] | [-0.140; 0.177] | [-0.140; 0.181] | [-0.143; 0.177] |
|  | Employed other | 0.158 | 0.164 | 0.074 | 0.046 | 0.052 | 0.056 |
|  |  | [-0.157; 0.474] | [-0.152; 0.480] | [-0.239; 0.388] | [-0.231; 0.324] | [-0.225; 0.329] | [-0.221; 0.333] |
|  | Employed temporary | -0.044 | -0.038 | -0.123 | 0.226 | 0.222 | 0.231 |
|  |  | [-0.518; 0.431] | [-0.514; 0.438] | [-0.601; 0.356] | [-0.189; 0.641] | [-0.191; 0.635] | [-0.181; 0.644] |
|  | Help in independent business | 0.078 | 0.101 | 0.082 | **0.301^*^** | **0.274^*^** | **0.275^*^** |
|  |  | [-0.095; 0.252] | [-0.075; 0.277] | [-0.099; 0.264] | **[ 0.146; 0.455]** | **[ 0.117; 0.430]** | **[ 0.118; 0.432]** |
|  | Owner of independent business | -0.019 | -0.011 | -0.045 | **0.149^*^** | **0.140^*^** | **0.143^*^** |
|  |  | [-0.156; 0.117] | [-0.148; 0.126] | [-0.180; 0.090] | **[ 0.033; 0.265]** | **[ 0.024; 0.256]** | **[ 0.027; 0.258]** |
|  | Side job at home | -0.096 | -0.058 | 0.075 | **0.407^*^** | 0.399 | 0.385 |
|  |  | [-0.525; 0.333] | [-0.491; 0.375] | [-0.354; 0.503] | **[ 0.005; 0.809]** | [-0.009; 0.807] | [-0.023; 0.793] |
|  | Employed |  |  |  |  |  |  |
| Working time | |  |  |  |  |  |  |
|  | 21 to 30 | 0.064 | 0.07 | 0.045 | 0.07 | 0.066 | 0.069 |
|  |  | [-0.088; 0.217] | [-0.083; 0.223] | [-0.109; 0.198] | [-0.069; 0.208] | [-0.072; 0.205] | [-0.069; 0.208] |
|  | 41 to 50 | -0.027 | -0.03 | -0.059 | -0.085 | -0.09 | -0.088 |
|  |  | [-0.173; 0.118] | [-0.175; 0.116] | [-0.202; 0.083] | [-0.229; 0.059] | [-0.239; 0.059] | [-0.237; 0.060] |
|  | 51 and over | -0.032 | -0.036 | -0.008 | -0.152 | -0.153 | -0.152 |
|  |  | [-0.202; 0.138] | [-0.206; 0.134] | [-0.187; 0.171] | [-0.314; 0.010] | [-0.319; 0.014] | [-0.320; 0.015] |
|  | Less than 20 | 0.111 | 0.115 | 0.058 | 0.071 | 0.069 | 0.072 |
|  |  | [-0.039; 0.260] | [-0.035; 0.265] | [-0.087; 0.202] | [-0.070; 0.211] | [-0.072; 0.210] | [-0.068; 0.213] |
|  | 21 to 40 (Ref.) |  |  |  |  |  |  |
| Job satisfaction | |  |  |  |  |  |  |
|  | 2 | 0.005 | 0.006 | -0.13 | -0.033 | -0.038 | -0.027 |
|  |  | [-0.125; 0.136] | [-0.125; 0.137] | [-0.264; 0.004] | [-0.148; 0.081] | [-0.153; 0.076] | [-0.140; 0.086] |
|  | 3 | **0.238^*^** | **0.240^*^** | -0.063 | **-0.174^*^** | **-0.172^*^** | -0.135 |
|  |  | **[ 0.073; 0.404]** | **[ 0.073; 0.407]** | [-0.237; 0.111] | **[-0.329; -0.020]** | **[-0.329; -0.015]** | [-0.291; 0.021] |
|  | 4 | 0.202 | 0.194 | -0.199 | -0.097 | -0.084 | -0.042 |
|  |  | [-0.002; 0.406] | [-0.012; 0.400] | [-0.414; 0.016] | [-0.323; 0.130] | [-0.314; 0.146] | [-0.275; 0.191] |
|  | 1 (Ref.) |  |  |  |  |  |  |
| Age |  | -0.008^*^ | -0.008 | 0.001 | **0.027^*^** | **0.024^*^** | **0.023^*^** |
|  |  | [-0.017; -0.000] | [-0.016; 0.001] | [-0.008; 0.010] | **[ 0.018; 0.037]** | **[ 0.014; 0.033]** | **[ 0.013; 0.033]** |
| gender: Female | | **0.178^*^** | **0.179^*^** | 0.073 | **-0.446^*^** | **-0.438^*^** | **-0.434^*^** |
|  |  | **[ 0.068; 0.288]** | **[ 0.059; 0.298]** | [-0.039; 0.186] | **[-0.543; -0.348]** | **[-0.547; -0.330]** | **[-0.542; -0.326]** |
| Education | |  |  |  |  |  |  |
|  | Elementary to middle school |  | -0.032 | 0.026 |  | **0.195^*^** | **0.190^*^** |
|  |  |  | [-0.205; 0.141] | [-0.149; 0.200] |  | **[ 0.012; 0.378]** | **[ 0.006; 0.374]** |
|  | High School |  | -0.016 | 0.008 |  | 0.113 | 0.112 |
|  |  |  | [-0.149; 0.118] | [-0.126; 0.141] |  | [-0.009; 0.235] | [-0.011; 0.235] |
|  | Junior college |  | -0.073 | -0.043 |  | 0.036 | 0.032 |
|  |  |  | [-0.293; 0.147] | [-0.258; 0.173] |  | [-0.160; 0.232] | [-0.164; 0.229] |
|  | Vocational school |  | -0.027 | 0.017 |  | 0.106 | 0.104 |
|  |  |  | [-0.224; 0.171] | [-0.177; 0.211] |  | [-0.072; 0.284] | [-0.074; 0.283] |
|  | University degree (Ref.) |  |  |  |  |  |  |
| Marital Satus | |  |  |  |  |  |  |
|  | Not Married and no common law spouse |  | 0.006 | -0.072 |  | -0.058 | -0.049 |
|  |  |  | [-0.120; 0.131] | [-0.199; 0.056] |  | [-0.191; 0.074] | [-0.181; 0.084] |
| Friend or family cover expenses: Yes | |  | 0.026 | 0.082 |  | 0.107 | 0.102 |
|  |  |  | [-0.136; 0.188] | [-0.079; 0.243] |  | [-0.036; 0.249] | [-0.040; 0.244] |
| Renting accommodation: Yes | |  | 0.087 | 0.046 |  | -0.086 | -0.073 |
|  |  |  | [-0.052; 0.227] | [-0.087; 0.180] |  | [-0.205; 0.033] | [-0.192; 0.046] |
| Private health insurance: Yes | |  | 0.069 | 0.043 |  | 0.049 | 0.048 |
|  |  |  | [-0.037; 0.175] | [-0.064; 0.151] |  | [-0.064; 0.162] | [-0.066; 0.163] |
| Self-reported health | |  |  | **0.090^*^** |  |  | -0.004 |
|  |  |  |  | **[ 0.032; 0.147]** |  |  | [-0.051; 0.042] |
| Comorbidity | |  |  | -0.014 |  |  | -0.035 |
|  |  |  |  | [-0.155; 0.127] |  |  | [-0.162; 0.093] |
| GHQ-case | |  |  | **1.384^*^** |  |  | **-0.094^*^** |
|  |  |  |  | **[ 1.266; 1.503]** |  |  | **[-0.184; -0.004]** |
| Outpatient at clinic or hospital | |  |  | 0.074 |  |  | 0.004 |
|  |  |  |  | [-0.053; 0.202] |  |  | [-0.086; 0.094] |
| Night at hospital | |  |  | 0.128 |  |  | 0.044 |
|  |  |  |  | [-0.057; 0.314] |  |  | [-0.130; 0.217] |
| Life satisfaction | |  |  | 0.046 |  |  | -0.085 |
|  |  |  |  | [-0.075; 0.168] |  |  | [-0.201; 0.030] |
|  | nobs | 6538 | 6538 | 6538 | 6538 | 6538 | 6538 |

##### Sample 3 (part a)

|  |  | Sleep quality | | | Weekdays sleep duration | | | Time to fall asleep | | |
| --- | --- | --- | --- | --- | --- | --- | --- | --- | --- | --- |
|  |  | (Modified Poisson) | | | (Linear) |  |  | (Modified Poisson) | | |
|  |  | **Demo Adj.** | **Socio-eco Adj.** | **Health ad.** | **Demo Adj.** | **Socio-eco Adj.** | **Health ad.** | **Demo Adj.** | **Socio-eco Adj.** | **Health ad.** |
|  | (Intercept) | -1.338^*^ | -1.329^*^ | -2.717^*^ | 5.103^*^ | 5.095^*^ | 5.189^*^ | -2.359^*^ | -2.043^*^ | -2.582^*^ |
|  |  | [-2.099; -0.578] | [-2.170; -0.487] | [-3.622; -1.812] | [ 4.349; 5.857] | [ 4.296; 5.894] | [ 4.362; 6.016] | [-3.382; -1.337] | [ -3.155; -0.930] | [ -3.736; -1.427] |
| Employment | |  |  |  |  |  |  |  |  |  |
|  | Company executive | 0.118 | 0.133 | 0.094 | 0.146 | 0.169 | 0.167 | 0.229 | 0.297 | 0.263 |
|  |  | [-0.189; 0.425] | [-0.175; 0.441] | [-0.216; 0.404] | [-0.125; 0.417] | [-0.106; 0.443] | [-0.108; 0.443] | [-0.116; 0.574] | [ -0.051; 0.644] | [ -0.085; 0.611] |
|  | Employed contract | 0.032 | 0.037 | 0.082 | -0.071 | -0.073 | -0.074 | -0.108 | -0.102 | -0.073 |
|  |  | [-0.218; 0.283] | [-0.214; 0.289] | [-0.171; 0.335] | [-0.287; 0.146] | [-0.289; 0.144] | [-0.290; 0.141] | [-0.422; 0.205] | [ -0.416; 0.213] | [ -0.389; 0.243] |
|  | Employed other | 0.157 | 0.161 | 0.075 | 0.216 | 0.221 | 0.216 | -0.027 | -0.01 | -0.017 |
|  |  | [-0.271; 0.585] | [-0.268; 0.589] | [-0.352; 0.503] | [-0.191; 0.624] | [-0.184; 0.625] | [-0.189; 0.621] | [-0.526; 0.471] | [ -0.512; 0.492] | [ -0.521; 0.488] |
|  | Employed temporary | 0.191 | 0.211 | -0.001 | 0.191 | 0.192 | 0.213 | 0.145 | 0.137 | 0.072 |
|  |  | [-0.473; 0.854] | [-0.456; 0.879] | [-0.689; 0.687] | [-0.452; 0.835] | [-0.455; 0.840] | [-0.440; 0.866] | [-0.641; 0.931] | [ -0.655; 0.930] | [ -0.721; 0.864] |
|  | Help in independent business | 0.03 | 0.041 | 0.033 | **0.395^*^** | **0.376^*^** | **0.374^*^** | 0.07 | 0.081 | 0.066 |
|  |  | [-0.214; 0.274] | [-0.205; 0.288] | [-0.220; 0.285] | **[ 0.155; 0.634]** | **[ 0.134; 0.618]** | **[ 0.134; 0.613]** | [-0.222; 0.362] | [ -0.215; 0.377] | [ -0.233; 0.365] |
|  | Owner of independent business | 0.073 | 0.08 | 0.063 | **0.194^*^** | **0.184^*^** | **0.181^*^** | 0.126 | 0.137 | 0.124 |
|  |  | [-0.115; 0.260] | [-0.109; 0.269] | [-0.128; 0.255] | **[ 0.020; 0.368]** | **[ 0.005; 0.362]** | **[ 0.002; 0.361]** | [-0.099; 0.351] | [ -0.090; 0.364] | [ -0.102; 0.350] |
|  | Side job at home | -0.091 | -0.073 | 0.126 | 0.407 | 0.415 | 0.384 | -0.119 | -0.171 | -0.103 |
|  |  | [-0.678; 0.497] | [-0.666; 0.520] | [-0.509; 0.761] | [-0.146; 0.960] | [-0.144; 0.974] | [-0.170; 0.938] | [-0.800; 0.562] | [ -0.861; 0.519] | [ -0.799; 0.593] |
|  | Employed |  |  |  |  |  |  |  |  |  |
| Working time | |  |  |  |  |  |  |  |  |  |
|  | 21 to 30 | 0.162 | 0.164 | 0.063 | 0.031 | 0.035 | 0.046 | 0.202 | 0.182 | 0.146 |
|  |  | [-0.063; 0.388] | [-0.063; 0.391] | [-0.169; 0.294] | [-0.174; 0.235] | [-0.171; 0.241] | [-0.160; 0.253] | [-0.058; 0.462] | [ -0.079; 0.443] | [ -0.116; 0.408] |
|  | 41 to 50 | 0.097 | 0.087 | -0.018 | -0.076 | -0.079 | -0.066 | -0.018 | -0.032 | -0.065 |
|  |  | [-0.117; 0.311] | [-0.127; 0.301] | [-0.225; 0.189] | [-0.272; 0.121] | [-0.278; 0.121] | [-0.269; 0.137] | [-0.274; 0.238] | [ -0.289; 0.226] | [ -0.327; 0.197] |
|  | 51 and over | -0.002 | -0.015 | 0.023 | -0.161 | -0.164 | -0.156 | -0.291 | -0.308 | -0.279 |
|  |  | [-0.255; 0.251] | [-0.269; 0.238] | [-0.231; 0.277] | [-0.378; 0.055] | [-0.382; 0.055] | [-0.377; 0.066] | [-0.623; 0.041] | [ -0.643; 0.026] | [ -0.615; 0.057] |
|  | Less than 20 | 0.122 | 0.125 | 0.038 | 0.11 | 0.111 | 0.121 | 0.243 | **0.260^*^** | 0.233 |
|  |  | [-0.104; 0.348] | [-0.103; 0.353] | [-0.194; 0.270] | [-0.086; 0.307] | [-0.084; 0.307] | [-0.075; 0.318] | [-0.003; 0.489] | **[ 0.012; 0.508]** | [ -0.020; 0.486] |
|  | 21 to 40 (Ref.) |  |  |  |  |  |  |  |  |  |
| Job satisfaction | |  |  |  |  |  |  |  |  |  |
|  | 2 | 0.017 | 0.011 | -0.153 | 0.006 | -0.004 | 0.009 | -0.077 | -0.114 | -0.181 |
|  |  | [-0.186; 0.219] | [-0.194; 0.215] | [-0.354; 0.047] | [-0.163; 0.175] | [-0.172; 0.164] | [-0.161; 0.178] | [-0.297; 0.143] | [ -0.336; 0.107] | [ -0.404; 0.043] |
|  | 3 | 0.248 | 0.242 | -0.087 | -0.141 | -0.146 | -0.101 | 0.142 | 0.06 | -0.083 |
|  |  | [-0.003; 0.500] | [-0.015; 0.499] | [-0.345; 0.172] | [-0.371; 0.088] | [-0.373; 0.082] | [-0.332; 0.130] | [-0.147; 0.431] | [ -0.232; 0.352] | [ -0.381; 0.214] |
|  | 4 | 0.216 | 0.2 | -0.232 | 0.02 | 0.023 | 0.083 | **0.375^*^** | 0.304 | 0.12 |
|  |  | [-0.100; 0.531] | [-0.119; 0.519] | [-0.556; 0.092] | [-0.488; 0.529] | [-0.451; 0.497] | [-0.385; 0.552] | **[ 0.019; 0.731]** | [ -0.056; 0.664] | [ -0.254; 0.493] |
|  | 1 (Ref.) |  |  |  |  |  |  |  |  |  |
| Age |  | -0.005 | -0.007 | 0.003 | **0.028^*^** | **0.024^*^** | **0.023^*^** | 0.006 | -0.003 | -0.001 |
|  |  | [-0.017; 0.007] | [-0.020; 0.006] | [-0.010; 0.016] | **[ 0.016; 0.039]** | **[ 0.012; 0.037]** | **[ 0.010; 0.036]** | [-0.008; 0.020] | [ -0.018; 0.013] | [ -0.017; 0.015] |
| gender: Female | | 0.213^*^ | 0.213^*^ | 0.088 | **-0.452^*^** | **-0.455^*^** | **-0.451^*^** | 0.153 | 0.088 | 0.063 |
|  |  | [ 0.063; 0.363] | [ 0.052; 0.374] | [-0.075; 0.251] | **[-0.594; -0.311]** | **[-0.607; -0.302]** | **[-0.604; -0.299]** | [-0.033; 0.339] | [ -0.108; 0.284] | [ -0.132; 0.258] |
| Education | |  |  |  |  |  |  |  |  |  |
|  | Elementary to middle school |  | 0.1 | 0.146 |  | 0.219 | 0.209 |  | **0.373^*^** | **0.361^*^** |
|  |  |  | [-0.133; 0.334] | [-0.092; 0.384] |  | [-0.003; 0.441] | [-0.010; 0.429] |  | **[ 0.073; 0.673]** | **[ 0.066; 0.655]** |
|  | High School |  | -0.001 | 0.045 |  | 0.162 | 0.156 |  | **0.311^*^** | **0.299^*^** |
|  |  |  | [-0.200; 0.198] | [-0.156; 0.246] |  | [-0.013; 0.337] | [-0.018; 0.330] |  | **[ 0.059; 0.563]** | **[ 0.048; 0.549]** |
|  | Junior college |  | -0.104 | -0.049 |  | 0.041 | 0.036 |  | 0.036 | 0.047 |
|  |  |  | [-0.428; 0.219] | [-0.376; 0.279] |  | [-0.243; 0.325] | [-0.249; 0.321] |  | [ -0.373; 0.446] | [ -0.361; 0.455] |
|  | Vocational school |  | -0.006 | 0.044 |  | 0.188 | 0.186 |  | **0.379^*^** | **0.382^*^** |
|  |  |  | [-0.308; 0.295] | [-0.254; 0.342] |  | [-0.065; 0.441] | [-0.067; 0.438] |  | **[ 0.034; 0.724]** | **[ 0.037; 0.727]** |
|  | University degree (Ref.) |  |  |  |  |  |  |  |  |  |
| Marital Satus | |  |  |  |  |  |  |  |  |  |
|  | Not Married and no common law spouse |  | 0.031 | -0.028 |  | -0.083 | -0.073 |  | 0.184 | 0.163 |
|  |  |  | [-0.151; 0.213] | [-0.213; 0.157] |  | [-0.358; 0.191] | [-0.342; 0.195] |  | [ -0.027; 0.396] | [ -0.049; 0.374] |
| Friend or family cover expenses: Yes | |  | 0.065 | 0.122 |  | 0.074 | 0.07 |  | 0.12 | 0.13 |
|  |  |  | [-0.162; 0.292] | [-0.106; 0.349] |  | [-0.166; 0.313] | [-0.168; 0.308] |  | [ -0.157; 0.397] | [ -0.145; 0.405] |
| Renting accommodation: Yes | |  | 0.085 | 0.047 |  | -0.025 | -0.007 |  | 0.007 | 0.004 |
|  |  |  | [-0.113; 0.283] | [-0.149; 0.243] |  | [-0.218; 0.167] | [-0.201; 0.188] |  | [ -0.227; 0.240] | [ -0.235; 0.242] |
| Private health insurance: Yes | |  | 0.057 | -0.007 |  | 0.064 | 0.063 |  | -0.143 | -0.159 |
|  |  |  | [-0.097; 0.211] | [-0.160; 0.146] |  | [-0.086; 0.214] | [-0.088; 0.213] |  | [ -0.315; 0.029] | [ -0.331; 0.014] |
| Self-reported health | |  |  | **0.099^*^** |  |  | 0.015 |  |  | **0.125^*^** |
|  |  |  |  | **[ 0.019; 0.179]** |  |  | [-0.054; 0.084] |  |  | **[ 0.035; 0.215]** |
| Comorbidity | |  |  | -0.116 |  |  | -0.066 |  |  | -0.003 |
|  |  |  |  | [-0.312; 0.080] |  |  | [-0.267; 0.136] |  |  | [ -0.233; 0.227] |
| GHQ-case | |  |  | **1.501^*^** |  |  | -0.124 |  |  | **0.413^*^** |
|  |  |  |  | **[ 1.341; 1.661]** |  |  | [-0.262; 0.014] |  |  | **[ 0.222; 0.604]** |
| Outpatient at clinic or hospital | |  |  | 0.059 |  |  | 0.037 |  |  | 0.088 |
|  |  |  |  | [-0.118; 0.235] |  |  | [-0.108; 0.182] |  |  | [ -0.118; 0.293] |
| Night at hospital | |  |  | 0.096 |  |  | 0.009 |  |  | 0.06 |
|  |  |  |  | [-0.216; 0.407] |  |  | [-0.249; 0.267] |  |  | [ -0.294; 0.415] |
| Life satisfaction | |  |  | 0.018 |  |  | -0.142 |  |  | 0.055 |
|  |  |  |  | [-0.159; 0.195] |  |  | [-0.332; 0.049] |  |  | [ -0.178; 0.288] |
|  | nobs | 3372 | 3372 | 3372 | 3372 | 3372 | 3372 | 3372 | 3372 | 3372 |

##### Sample 3 (part b)

|  |  | Waking up at night | | | Early hours waking up | | | Night urination | | |
| --- | --- | --- | --- | --- | --- | --- | --- | --- | --- | --- |
|  |  | (Modified Poisson) | | | (Modified Poisson) | | | (Modified Poisson) | | |
|  |  | **Demo Adj.** | **Socio-eco Adj.** | **Health ad.** | **Demo Adj.** | **Socio-eco Adj.** | **Health ad.** | **Demo Adj.** | **Socio-eco Adj.** | **Health ad.** |
|  | (Intercept) | -3.140^*^ | -3.059^*^ | -3.919^*^ | -3.643^*^ | -3.462^*^ | -3.462^*^ | -3.602^*^ | -3.734^*^ | -3.824^*^ |
|  |  | [-3.953; -2.327] | [ -3.960; -2.159] | [ -4.848; -2.989] | [-4.528; -2.758] | [-4.444; -2.480] | [-4.444; -2.480] | [-4.497; -2.707] | [-4.728; -2.741] | [-4.856; -2.792] |
| Employment | |  |  |  |  |  |  |  |  |  |
|  | Company executive | 0.076 | 0.124 | 0.102 | **0.341^*^** | **0.395^*^** | **0.395^*^** | -0.258 | -0.206 | -0.231 |
|  |  | [-0.246; 0.399] | [ -0.200; 0.448] | [ -0.227; 0.431] | **[ 0.036; 0.645]** | **[ 0.089; 0.700]** | **[ 0.089; 0.700]** | [-0.649; 0.133] | [-0.602; 0.191] | [-0.624; 0.163] |
|  | Employed contract | -0.014 | 0.003 | 0.026 | 0.08 | 0.093 | 0.093 | 0.023 | 0.011 | 0.026 |
|  |  | [-0.276; 0.248] | [ -0.260; 0.266] | [ -0.242; 0.294] | [-0.195; 0.355] | [-0.185; 0.370] | [-0.185; 0.370] | [-0.249; 0.294] | [-0.263; 0.284] | [-0.248; 0.301] |
|  | Employed other | 0.131 | 0.134 | 0.119 | 0.054 | 0.062 | 0.062 | -0.135 | -0.106 | -0.122 |
|  |  | [-0.290; 0.551] | [ -0.286; 0.554] | [ -0.303; 0.542] | [-0.414; 0.522] | [-0.406; 0.531] | [-0.406; 0.531] | [-0.639; 0.370] | [-0.611; 0.400] | [-0.628; 0.384] |
|  | Employed temporary | 0.053 | 0.058 | -0.019 | 0.023 | 0.048 | 0.048 | -0.024 | -0.104 | -0.109 |
|  |  | [-0.698; 0.803] | [ -0.693; 0.810] | [ -0.769; 0.732] | [-0.793; 0.838] | [-0.768; 0.865] | [-0.768; 0.865] | [-0.837; 0.790] | [-0.927; 0.718] | [-0.937; 0.720] |
|  | Help in independent business | 0.118 | 0.142 | 0.129 | 0.083 | 0.096 | 0.096 | -0.083 | 0.058 | 0.076 |
|  |  | [-0.131; 0.367] | [ -0.111; 0.396] | [ -0.125; 0.383] | [-0.200; 0.366] | [-0.192; 0.383] | [-0.192; 0.383] | [-0.377; 0.210] | [-0.236; 0.351] | [-0.220; 0.372] |
|  | Owner of independent business | 0.183 | **0.203^*^** | 0.18 | 0.153 | 0.167 | 0.167 | -0.041 | 0.012 | 0.014 |
|  |  | [-0.008; 0.375] | **[ 0.010; 0.397]** | [ -0.017; 0.377] | [-0.058; 0.364] | [-0.047; 0.381] | [-0.047; 0.381] | [-0.257; 0.175] | [-0.204; 0.229] | [-0.205; 0.232] |
|  | Side job at home | 0.407 | 0.411 | 0.504 | 0.384 | 0.37 | 0.37 | -0.161 | -0.126 | -0.116 |
|  |  | [-0.085; 0.899] | [ -0.091; 0.912] | [ -0.004; 1.011] | [-0.169; 0.938] | [-0.191; 0.930] | [-0.191; 0.930] | [-0.809; 0.488] | [-0.783; 0.532] | [-0.780; 0.547] |
|  | Employed |  |  |  |  |  |  |  |  |  |
| Working time | |  |  |  |  |  |  |  |  |  |
|  | 21 to 30 | 0.182 | 0.169 | 0.117 | 0.097 | 0.084 | 0.084 | 0.046 | 0.034 | 0.022 |
|  |  | [-0.045; 0.408] | [ -0.059; 0.398] | [ -0.112; 0.346] | [-0.145; 0.339] | [-0.159; 0.326] | [-0.159; 0.326] | [-0.204; 0.296] | [-0.218; 0.287] | [-0.231; 0.276] |
|  | 41 to 50 | 0.014 | 0.006 | -0.035 | 0.005 | -0.005 | -0.005 | -0.037 | -0.043 | -0.053 |
|  |  | [-0.198; 0.226] | [ -0.206; 0.218] | [ -0.250; 0.181] | [-0.222; 0.232] | [-0.231; 0.220] | [-0.231; 0.220] | [-0.268; 0.195] | [-0.275; 0.189] | [-0.287; 0.181] |
|  | 51 and over | -0.089 | -0.104 | -0.086 | -0.209 | -0.225 | -0.225 | -0.187 | -0.2 | -0.182 |
|  |  | [-0.352; 0.174] | [ -0.367; 0.160] | [ -0.355; 0.184] | [-0.491; 0.073] | [-0.507; 0.057] | [-0.507; 0.057] | [-0.498; 0.125] | [-0.507; 0.107] | [-0.491; 0.126] |
|  | Less than 20 | 0.098 | 0.096 | 0.072 | 0.011 | 0.013 | 0.013 | 0.025 | 0.073 | 0.064 |
|  |  | [-0.113; 0.308] | [ -0.115; 0.308] | [ -0.142; 0.286] | [-0.221; 0.242] | [-0.221; 0.246] | [-0.221; 0.246] | [-0.231; 0.281] | [-0.189; 0.335] | [-0.194; 0.321] |
|  | 21 to 40 (Ref.) |  |  |  |  |  |  |  |  |  |
| Job satisfaction | |  |  |  |  |  |  |  |  |  |
|  | 2 | 0.061 | 0.041 | -0.05 | 0.196 | 0.171 | 0.171 | 0.045 | 0.025 | -0.007 |
|  |  | [-0.148; 0.271] | [ -0.171; 0.253] | [ -0.263; 0.162] | [-0.026; 0.419] | [-0.052; 0.393] | [-0.052; 0.393] | [-0.170; 0.261] | [-0.191; 0.240] | [-0.224; 0.211] |
|  | 3 | **0.397^*^** | **0.372^*^** | 0.168 | **0.428^*^** | **0.391^*^** | **0.391^*^** | 0.056 | -0.054 | -0.101 |
|  |  | **[ 0.141; 0.654]** | **[ 0.109; 0.634]** | [ -0.107; 0.443] | **[ 0.154; 0.702]** | **[ 0.114; 0.669]** | **[ 0.114; 0.669]** | [-0.241; 0.353] | [-0.354; 0.247] | [-0.410; 0.208] |
|  | 4 | **0.404^*^** | **0.362^*^** | 0.092 | **0.442^*^** | **0.401^*^** | **0.401^*^** | **0.432^*^** | 0.292 | 0.242 |
|  |  | **[ 0.074; 0.735]** | **[ 0.025; 0.699]** | [ -0.248; 0.432] | **[ 0.096; 0.789]** | **[ 0.050; 0.751]** | **[ 0.050; 0.751]** | **[ 0.092; 0.773]** | [-0.051; 0.635] | [-0.107; 0.590] |
|  | 1 (Ref.) |  |  |  |  |  |  |  |  |  |
| Age |  | **0.022^*^** | **0.019^*^** | **0.024^*^** | **0.028^*^** | **0.023^*^** | **0.023^*^** | **0.030^*^** | **0.030^*^** | **0.027^*^** |
|  |  | **[ 0.010; 0.034]** | **[ 0.006; 0.032]** | **[ 0.011; 0.038]** | **[ 0.015; 0.041]** | **[ 0.009; 0.038]** | **[ 0.009; 0.038]** | **[ 0.017; 0.044]** | **[ 0.015; 0.044]** | **[ 0.012; 0.042]** |
| gender: Female | | -0.003 | -0.046 | -0.091 | **-0.210^*^** | **-0.261^*^** | **-0.261^*^** | -0.04 | **-0.299^*^** | **-0.309^*^** |
|  |  | [-0.163; 0.156] | [ -0.215; 0.124] | [ -0.261; 0.079] | **[-0.385; -0.036]** | **[-0.444; -0.078]** | **[-0.444; -0.078]** | [-0.221; 0.142] | **[-0.495; -0.104]** | **[-0.505; -0.113]** |
| Education | |  |  |  |  |  |  |  |  |  |
|  | Elementary to middle school |  | 0.232 | 0.208 |  | **0.294^*^** | **0.294^*^** |  | 0 | 0.014 |
|  |  |  | [ -0.017; 0.480] | [ -0.048; 0.465] |  | **[ 0.022; 0.567]** | **[ 0.022; 0.567]** |  | [-0.273; 0.273] | [-0.263; 0.291] |
|  | High School |  | 0.158 | 0.141 |  | 0.219 | 0.219 |  | -0.027 | -0.013 |
|  |  |  | [ -0.053; 0.370] | [ -0.073; 0.355] |  | [-0.012; 0.449] | [-0.012; 0.449] |  | [-0.254; 0.201] | [-0.244; 0.217] |
|  | Junior college |  | 0.167 | 0.163 |  | 0.123 | 0.123 |  | 0.196 | 0.213 |
|  |  |  | [ -0.181; 0.514] | [ -0.183; 0.508] |  | [-0.327; 0.573] | [-0.327; 0.573] |  | [-0.180; 0.573] | [-0.166; 0.592] |
|  | Vocational school |  | 0.197 | 0.193 |  | 0.317 | 0.317 |  | 0.143 | 0.145 |
|  |  |  | [ -0.103; 0.497] | [ -0.104; 0.491] |  | [-0.005; 0.639] | [-0.005; 0.639] |  | [-0.180; 0.466] | [-0.180; 0.470] |
|  | University degree (Ref.) |  |  |  |  |  |  |  |  |  |
| Marital Satus | |  |  |  |  |  |  |  |  |  |
|  | Not Married and no common law spouse |  | 0.018 | -0.026 |  | 0.047 | 0.047 |  | **0.909^*^** | **0.911^*^** |
|  |  |  | [ -0.178; 0.214] | [ -0.222; 0.170] |  | [-0.174; 0.269] | [-0.174; 0.269] |  | **[ 0.705; 1.112]** | **[ 0.707; 1.114]** |
| Friend or family cover expenses: Yes | |  | -0.019 | 0.012 |  | -0.015 | -0.015 |  | 0.043 | 0.051 |
|  |  |  | [ -0.259; 0.221] | [ -0.230; 0.253] |  | [-0.267; 0.238] | [-0.267; 0.238] |  | [-0.221; 0.307] | [-0.214; 0.316] |
| Renting accommodation: Yes | |  | 0.15 | 0.144 |  | 0.076 | 0.076 |  | 0.019 | 0.029 |
|  |  |  | [ -0.050; 0.351] | [ -0.062; 0.350] |  | [-0.142; 0.293] | [-0.142; 0.293] |  | [-0.218; 0.255] | [-0.208; 0.266] |
| Private health insurance: Yes | |  | -0.011 | -0.029 |  | -0.047 | -0.047 |  | -0.011 | -0.029 |
|  |  |  | [ -0.166; 0.144] | [ -0.185; 0.128] |  | [-0.210; 0.115] | [-0.210; 0.115] |  | [-0.180; 0.158] | [-0.198; 0.141] |
| Self-reported health | |  |  | **0.182^*^** |  |  |  |  |  | 0.043 |
|  |  |  |  | **[ 0.102; 0.262]** |  |  |  |  |  | [-0.044; 0.129] |
| Comorbidity | |  |  | **-0.094** |  |  |  |  |  | 0.143 |
|  |  |  |  | **[ -0.298; 0.110]** |  |  |  |  |  | [-0.071; 0.358] |
| GHQ-case | |  |  | **0.596^*^** |  |  |  |  |  | **0.213^*^** |
|  |  |  |  | **[ 0.443; 0.750]** |  |  |  |  |  | **[ 0.032; 0.393]** |
| Outpatient at clinic or hospital | |  |  | 0.013 |  |  |  |  |  | 0.129 |
|  |  |  |  | [ -0.182; 0.208] |  |  |  |  |  | [-0.066; 0.323] |
| Night at hospital | |  |  | -0.049 |  |  |  |  |  | -0.066 |
|  |  |  |  | [ -0.382; 0.284] |  |  |  |  |  | [-0.349; 0.218] |
| Life satisfaction | |  |  | 0.118 |  |  |  |  |  | -0.134 |
|  |  |  |  | [ -0.081; 0.316] |  |  |  |  |  | [-0.368; 0.099] |
|  | nobs | 3372 | 3372 | 3372 | 3372 | 3372 | 3372 | 3372 | 3372 | 3372 |
